# Systematic common and rare variant association testing in 392,030 whole genomes in *All of Us*

**DOI:** 10.64898/2026.05.08.26350964

**Authors:** Wenhan Lu, Robert J. Carroll, Matthew Solomonson, Jeremy Guez, Megan K. He, Daniel J. Marten, Alejandro Martinez-Carrasco, Ying Wang, Connor S. Dowd, Masahiro Kanai, Bram L. Gorissen, Aymone Jeanne S. Kouame, James Brogan, Bennett J. Waxse, Ryan Samarakoon, Justin A. Cook, Jun Qian, Yu Zhou, Karmel W. Choi, Melissa Basford, Michael Lyons, Jodell E. Linder, Samantha Stewart, Namrata Gupta, Patrick Schultz, Daniel Goldstein, Christopher Llanwarne, Jacqueline I. Goldstein, Edmund G. C. Higham, Daniel C. King, Duncan S. Palmer, Jared S. Elenbaas, Gregory K. Rohlicek, Qin He, Julia K. Goodrich, The All of Us Research Program Genomics Investigators, Jordan W. Smoller, Lee Lichtenstein, Stacey B. Gabriel, Alicia R. Martin, Jason H. Karnes, Scott J. Hebbring, Mark J. Daly, Tara Dutka, Anjene Musick, Joshua C. Denny, Wei Zhou, Dan M. Roden, Benjamin M. Neale, Konrad J. Karczewski

## Abstract

Large-scale genome-wide association studies (GWAS) and rare variant association studies (RVAS) from population biobanks provide valuable resources for gene discovery in complex human traits. We present an analysis of the *All of Us* Research Program v8 release, which includes whole genome sequencing data and harmonized phenotypic information of 392,030 participants after quality control, enabling a unified investigation of rare and common variants across a spectrum of human traits and diseases. We build an extensive phenome- and genome-wide (“All by All”) computational framework to perform GWAS and RVAS on 3,602 phenotypes and identify 49,863 approximately independent, high-quality single-variant and gene-level associations. Meta-analyses of *All of Us* and UK Biobank, with sample sizes as large as 786,871 participants, further enhance statistical power and find 193 pLoF gene-phenotype associations that are not significant in either cohort alone, including 22 associations not highlighted by previous studies. We also present a public interactive browser that integrates association results for common and rare variants to facilitate interpretation and rapid querying of summary statistics, along with supporting documentation, and a Featured Workspace in the *All of Us* Researcher Workbench. Our framework will apply to iterative data releases as *All of Us* grows, empowering researchers worldwide to uncover insights into the functional effects of genetic components on complex traits and diseases.

## Introduction

Large-scale repositories integrating genomic and phenotypic data have enabled systematic discovery of genetic underpinnings of human traits and diseases through phenome-and genome-wide association studies (GWAS) and rare variant association studies (RVAS)^1–9^. Population-based biobanks such as UK Biobank (UKB)^1,6,7,10–12^, Million Veteran Program (MVP)^5,13^, Mass General Brigham Biobank (MGB)^14,15^, FinnGen^4^, Biobank Japan (BBJ)^3^, and other national and regional biobanks^16–18^ have identified thousands of genetic associations, providing insights into disease biology and nominating candidate therapeutic targets^19,20^. These efforts have demonstrated that increasing cohort size and phenotypic breadth substantially improve power to detect genetic associations. Moreover, combining data across multiple biobanks^21^ enables more comprehensive characterization of genetic effects across genetic ancestries and clinical contexts, thus improving discovery power and advancing the translation of human genetics into biological and clinical insights.

The *All of Us* Research Program broadens the landscape of existing biobank efforts as a US-based cohort that has enrolled over 866,000 participants nationwide. Through recruitment across diverse healthcare settings and linkage to clinical and participant-contributed data, *All of Us* enables continuous and heterogeneous phenotypic characterization at scale. With an emphasis on communities historically underrepresented in biomedical research, the program captures a more representative spectrum of human genetic variation across the US population. The program is iteratively releasing data, with the latest release (v8) including whole genome sequencing (WGS) data for 414,830 individuals together with curated extensive biological measures and linked electronic health records^2,22^. These data provide the opportunity to nominate therapeutic targets, enabling association testing across the phenome, including GWAS, which have identified loci such as *TYK2* for autoimmune disease^23^, and RVAS, which have identified loci such as *PCSK9* for heart disease^24^; both of these discoveries have led to approved therapies for their respective diseases^25,26^. This combination of scale, representation, and genomic depth establishes *All of Us* as a critical resource for large-scale genetic association studies and for more comprehensive characterization of genetic effects across diverse genetic backgrounds and phenotypic definitions shaped by both biology and healthcare processes.

Here, we present results from an *All by All* analysis of *All of Us*, encompassing comprehensive single-variant association analyses of common and exonic variants and gene-level burden analyses of rare protein-coding variants across 392,030 participants post quality control (QC) for 3,602 unique phenotypes, leading to 1.337 trillion association tests, which, after results QC, result in 48,831 significant single-variant signals (linkage disequilibrium (LD)-pruned r^2^ < 0.1; *p*_single-variant_ < 5 ⨉ 10^−8^) and 1,032 significant gene-burden associations (*p*_gene-burden_ < 6.7 ⨉ 10^−7^) at maximum minor allele frequency (maxMAF) 0.1%. Of these, we identify 109 gene-level pLoF associations not significant in the UK Biobank exome data^7^. Furthermore, we performed cross-biobank meta-analyses integrating associations from *All by All* and UK Biobank for both single-variant GWAS^6^ and rare variant gene-based burden results^7^, encompassing up to 786,871 samples and 222 phenotypes. By combining summary statistics from two large and complementary biobanks, our meta-analysis increases both cumulative allele counts and allelic diversity, enabling the detection of gene-level associations not observable within individual cohorts. Consistent with this expectation, we identify 22 novel pLoF signals that reach significance exclusively in the cross-biobank meta-analysis and are not detected in either biobank individually or reported in prior studies based on automated literature curation, reinforcing the critical role of ultra-large sample sizes in gene discovery.

To support reproducibility and enable broad use by the global research community, we have released the full *All by All* association results and cross-biobank meta-analysis summary statistics through the Controlled Tier of the *All of Us* Researcher Workbench and through a public interactive data browser. These resources establish one of the largest publicly available association datasets spanning both genome-wide and exome-wide variations, enabling systematic gene discovery, prioritization of therapeutic targets, improved characterization of pleiotropic genetic effects, development of polygenic risk scores, and ultimately advancing our understanding of the genetic architecture and biological basis of human disease.

## Results

### A comprehensive large-scale multi-group All by All resource based on All of Us

The *All of Us* Research Program has curated the genomic and phenotypic data used in this study through established quality control protocols^2^, which we incorporate for large-scale association testing. In this paper, we consider the genetic similarity of each participant to individuals in reference panels as recommended^27^, and generate six genetic similarity groups (**Extended Data Table 1**) for association testing. We recognize that socially defined ancestry and ethnic categories have historically been misused to justify claims about innate biological differences, contributing to harm across populations. Consistent with extensive prior work, genetic ancestry is continuous, with greater variation within than between populations^28^. In this study, we use discrete ancestry groupings solely as an analytic strategy to limit population stratification in large-scale association analyses and to control type I error. These groupings are imperfect proxies for underlying genetic ancestry and population structure and should not be interpreted as biologically discrete or socially meaningful categories. We refer to these hereafter as ‘groups’ and denote them using three-letter codes, following prior work^6^. Here, we combined WGS data of 414,830 participants with harmonized phenotypic data in an “All by All” framework to identify genetic associations with common and rare variants across a broad range of phenotypes, including physical measurements, lab measurements, diseases (PhecodeX^29^), and prescriptions (Anatomical Therapeutic Chemical [ATC] Classification System) derived from electronic health records (EHR), self-reported phenotypes from mental health well-being and personal medical history or personal family health history surveys, and randomly simulated phenotypes for result calibration (**Figure 1a, 1b**).

**Figure 1.**
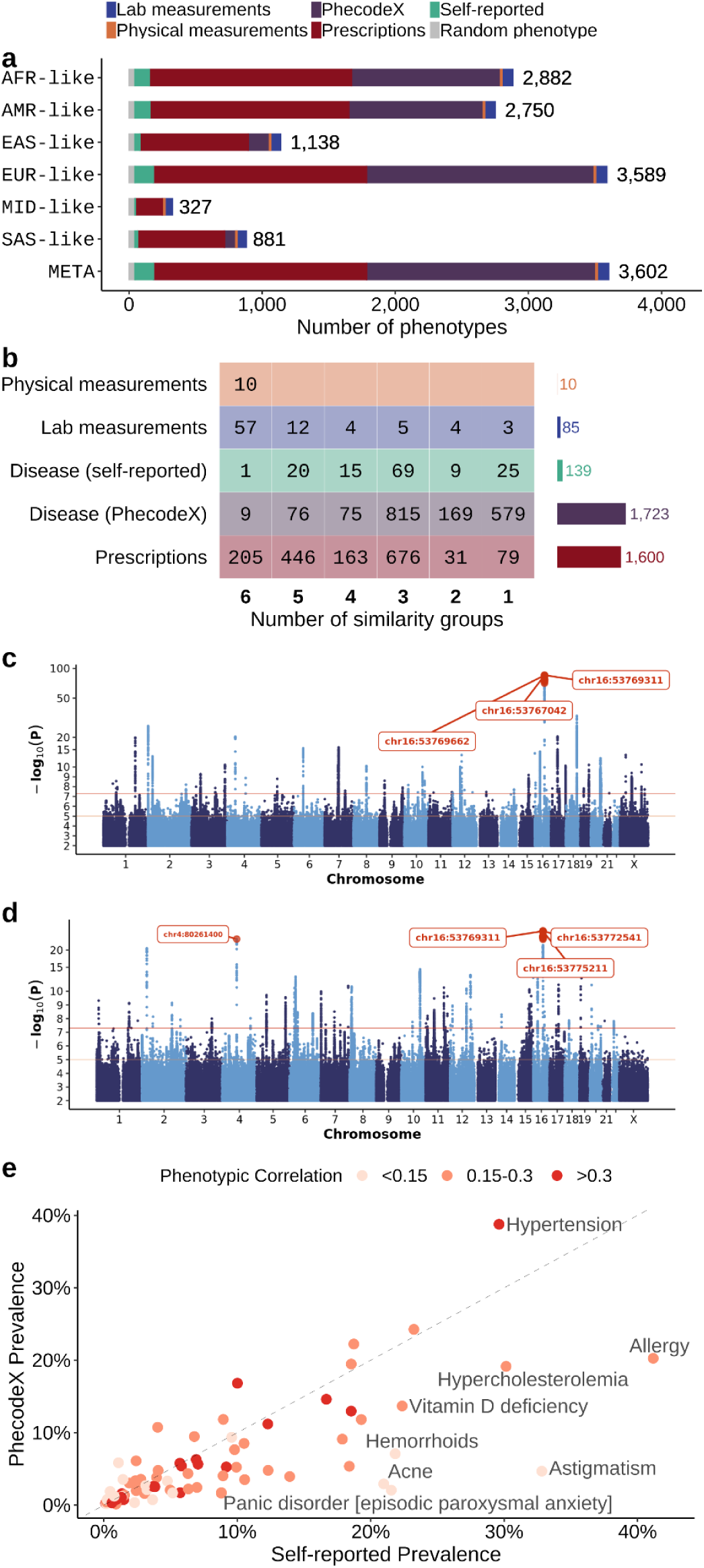
Summary of *All by All* phenotype data. **a**, Number of phenotypes (x-axis) stratified by genetic similarity group (y-axis; see **Supplementary Information**) and phenotype category (color). **b**, Number of phenotypes (cells) within each phenotype category (y-axis) stratified by the number of genetic similarity groups for which association testing was performed (x-axis); bars on the right indicate total phenotypes analyzed per category. **c-d,** Manhattan plots of meta-analyzed common variant associations for **(c)** obesity (EM_236.1; n_cases_= 67,952, n_controls_= 212,095) and **(d)** essential hypertension (CV_401.1; n_cases_= 106,344, n_controls_= 176,132)**. e,** Prevalence comparison for 87 diseases mapped between self-reported diseases from personal family health history surveys (x-axis) and PhecodeX (y-axis). Phenotypes with prevalences differing by more than 20% (i.e., >1.2 or <0.8) and at least one prevalence above 20% are highlighted. Each point represents a matched phenotype pair, with color indicating the corresponding phenotypic correlation. The dashed line denotes y=x.

To ensure data quality and robust association results, we performed systematic sample curation and quality control of both genotype and phenotype data (**Supplementary Figure 2**), resulting in 392,030 participants and 3,602 unique phenotypes (100 quantitative and 3,502 binary traits) with over 200 cases (or samples defined) in at least one of the six similarity groups. Building on these multi-group phenome- and genome-wide resources, we applied SAIGE^30^ and SAIGE-GENE+^31–33^ (**Extended Data Figure 1**) for single-variant and gene-level rare variant association analyses, respectively. For gene-level burden analyses, we aggregated rare protein-coding variants by functional annotation and MAF. In total, we performed 1.333 trillion single-variant tests and 3.726 billion burden tests across similarity groups and meta-analysis (**Supplementary Table 5**). After results QC and restricting to a maximum set of phenotypes with phenotypic r^2^ < 0.5 (**Supplementary Figure 13**), we identify 48,831 significant single-variant signals (LD-pruned r^2^ < 0.1; *p*_single-variant_ < 5 ⨉ 10^−8^) and 1,032 gene-burden significant associations (*p*_gene-burden_ < 6.7 ⨉ 10^−7^) at maxMAF 0.1% across six groups plus meta-analysis (**Supplementary Table 9; Supplementary Figure 16**; **Supplementary Data 1-2**). Unless otherwise stated, all results described hereafter are restricted to associations that passed results QC (**Supplementary Figure 19**).

For instance, we identified strong associations between prescription of GLP-1 analogs (ATC: A10BJ) and common variants near *TCF7L2* and *FTO* (**Extended Data Figure 2**), which are loci with well-documented roles in type 2 diabetes^34^ and obesity^35,36^, respectively. Notably, sex-stratified analyses showed stronger *TCF7L2* (type 2 diabetes locus) associations in males and stronger *FTO* (obesity locus) associations in females with GLP-1 analog usage. These sex-stratified associations likely reflect differences in statistical power for gene detection, owing at least partially to differences in prescribing patterns, as also suggested by the phenotypic correlations between GLP-1 analog prescription and disease diagnoses (r_GLP-1∼T2D; female_= 0.333, r_GLP-1∼T2D; male_= 0.352, OR_T2D∼GLP-1 ⨉ sex_=2.13 [p = 8.59 ⨉ 10-59]; r_GLP-1∼obesity; female_= 0.288, r_GLP-1∼obesity; male_= 0.289, OR_Obesity∼GLP-1 ⨉ sex_=1.03 [*p* = 0.44]; **Extended Data Figure 2d-g**). These results highlight how medication-based phenotypes can capture the interplay between genetic predisposition, disease indication, and prescribing patterns. Additionally, the depth and breadth of the EHR information enabled well-powered investigations of genetic mechanisms across diseases. In particular, we identified 75 and 74 independent common variant associations for the highly polygenic traits, obesity (**Figure 1c**) and essential hypertension (**Figure 1d**), respectively, recapitulating several established GWAS signals. Both traits exhibited strong peaks at loci near gene *FTO*^35–37^, consistent with its well-studied role in obesity-related traits and metabolic risk, including hypertension. Essential hypertension also showed prominent associations at chromosome 4 near gene *FGF5*, a canonical blood-pressure locus^38^.

Leveraging the phenome-wide meta-analysis results, we evaluated the concordance between self-reported and clinically billed (PhecodeX) diseases to assess how phenotype definition influences the performance and interpretation of genetic associations. Across 87 manually mapped diseases, observed prevalences were highly correlated between self-reported diseases and PhecodeX (Pearson’s r = 0.74, *p* = 4.07 ⨉ 10^−16^; **Figure 1e**), although self-reported diseases exhibited slightly higher prevalences for conditions with milder symptoms or routine management, such as acne, allergy, and vitamin D deficiency. These patterns may reflect greater sensitivity to mild or symptom-based conditions in self-reported responses and to more severe, complex diseases captured through clinical ascertainment in electronic health records, illustrating their complementary strengths and the value of integrating both sources for more precise health assessment.

### Consistent signals across genetic similarity groups empower association discovery

Of 140 gene-phenotype associations marginally significant (*p*_gene-burden_ < 1 ⨉ 10^−5^) in at least two similarity groups at maxMAF 0.1%, we observed no associations with burden effect sizes in opposite directions across genetic similarity groups, consistent with previous literature showing few bona fide differences in biological effects across groups^39^. Across the 34,889 LD-pruned (r^2^ < 0.1) meta-analyzed variant-phenotype associations, 1,676 were significant (*p*_single-variant_ < 5 ⨉ 10^−8^) for more than one genetic similarity group, among which only 7 (0.4%) exhibited opposite effect directions, and these associations showed substantially higher heterogeneity among groups than the remaining associations (median Cochran’s Q = 277 vs. 4.65; p_Wilcoxon rank sum test_ = 4.95 ⨉ 10-6).

As expected, the number of significant rare variant burden signals (maxMAF at 0.1%) was positively correlated with group-specific sample sizes (r = 0.995, *p* = 3.302 ⨉ 10^−6^; **Supplementary Table 9**), emphasizing the critical roles of large sample size and statistical power in detecting genetic associations and motivating meta-analysis approaches to enhance statistical power for gene discovery. We therefore performed meta-analyses of both GWAS and RVAS results across similarity groups for phenotypes with over 200 cases (or samples defined) in at least one group, using a fixed-effect inverse-variance weighted model for single-variant tests and Stouffer’s method^40^ for gene-based rare variant tests.

For phenotypes analyzed in at least the three largest genetic similarity groups, we observe 566 unique pLoF gene-phenotype associations, among which 240 (42.4%) were identified as significant only through meta-analysis (**Figure 2a**). We quantify the contribution of each similarity group to the corresponding significant associations in meta-analysis using the group-specific cumulative allele frequencies (CAF), test statistics, and effect directions. Indeed, for most associations significant exclusively through meta-analysis, the average contribution of each similarity group (proportion of the unweighted Z-score relative to the total) was strongly correlated with group-specific sample sizes (Pearson’s r = 0.997, *p* = 1.61 ⨉ 10^−5^; **Figure 2b, Supplementary Figures 24** and **25**, **Supplementary Data 3**), reiterating the need to include as many individuals as possible in these analyses.

**Figure 2.**
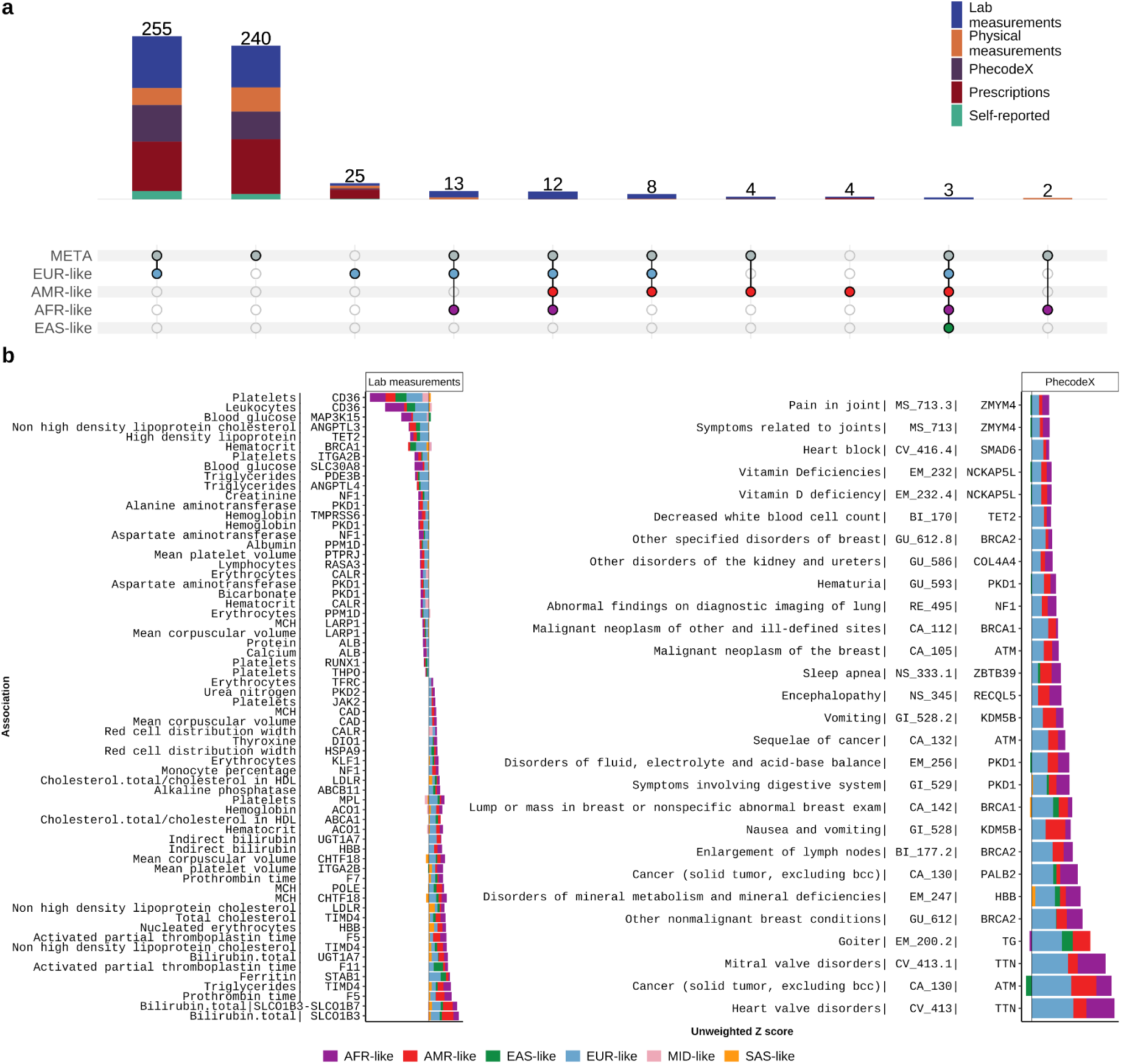
Larger cohorts drive association discoveries. **a,** Number of significant pLoF gene burden associations with maxMAF = 0.1% across combinations of genetic similarity groups (colored dots in the bottom matrix), restricted to phenotypes analyzed in at least the three largest groups (AFR-like, AMR-like, and EUR-like). Bars represent the number of genes with cross-group associations, as defined by the dot combinations in the matrix below, colored and partitioned by phenotype category. **b,** Unweighted Z-scores contributing to meta-analysis associations, defined as 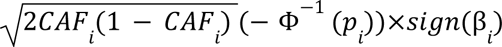, where 𝐶𝐴𝐹_i_, 𝑝_i_ and β_i_ are the combined allele frequency, *p* value, and beta effect size estimates from the pLoF burden test for group 𝑖. Each bar represents a significant QCed meta-analysis association (maxMAF = 0.1%), for which no individual genetic similarity group achieved significance. The magnitude of the unweighted Z-score from each genetic similarity group is indicated by the size of the corresponding colored blocks. Colors in the matrix in **a** and in **b** are consistent.

### Biological insight from All by All meta-analysis

We identified 568 associations for rare pLoF variants after result QC, but as in previous work^6^, quantifying novelty is challenging due to variation in phenotype and locus definitions. We replicated 169 pLoF burden associations in Genebass (*p*_gene-burden_ < 6.7 ⨉ 10^−7^ in both biobanks) at maxMAF of 1% (**Supplementary Figure 18**; **Supplementary Data 4**), such as those between pLoF variants in *GIGYF1* and blood glucose level (*p*_gene-burden_ = 1.19 ⨉ 10^−18^; **Figure 3a**), HbA1c (*p*_gene-burden_ = 1.03 ⨉ 10^−12^), and a set of medications related to blood glucose-lowering drugs or insulins (**Supplementary Data 5**), which exhibits a critical role in regulating insulin signaling and protecting against diabetes as shown previously^7,10,11^. We also highlight a putative novel rare variant association between elevated triglycerides and rare pLoF variants in gene *TIMD4* (*p*_gene-burden_ = 3.14 ⨉ 10^−10^; **Figure 3b**), which is significant only in the cross-group meta-analysis, but not any single similarity group. Common variants near *TIMD4* have previously been found to be associated with serum lipid levels and cardiometabolic disease risk^41–44^, and here we identify rare pLoF variants in *TIMD4* with shared etiology underlying lipid variation and cardiovascular disease, indicating a directionality of the association.

**Figure 3.**
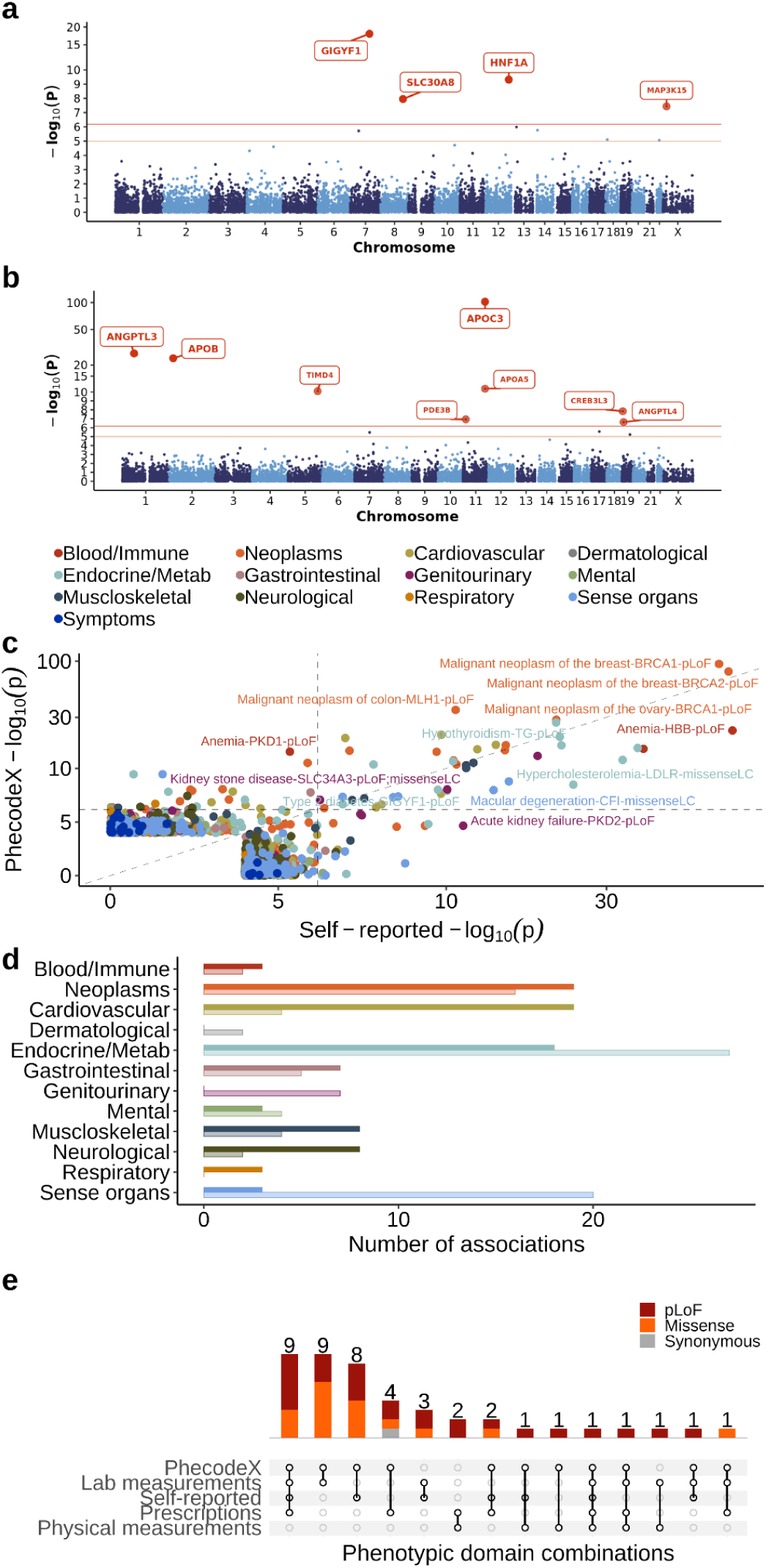
*All by All* meta-analysis of gene-level rare variant burden associations. **a-b**, Manhattan plots of *p* values from the meta-analysis of pLoF rare variant burden tests at maxMAF of 0.001 for **(a)** blood glucose (code: 3004501; N = 215,067) and **(b)** triglycerides (code: 3022192; N = 143,386). **c-d,** Comparison of 87 mapped diseases between personal and family health history (self-reported) and PhecodeX (derived from electronic health record). **c,** Comparison of burden association *p* values between self-reported (x-axis) and PhecodeX (y-axis), restricted to associations with *p*_gene-burden_ < 10^−4^ in either self-reported or PhecodeX. **d,** Number of associations (y-axis) that are significant in only self-reported or PhecodeX (transparency) across disease categories (x-axis). Colors are consistent in **c** and **d,** representing the disease categories initially defined by PhecodeX. **e,** 44 gene-annotation pairs with burden associations spanning more than one phenotypic domain. Each bar represents the number of pairs with associations for the corresponding combination shown in the matrix below; colors indicate functional category. The matrix represents combinations across phenotypic domains.

As expected based on consistency of prevalences (**Figure 1e**), burden association results were also largely consistent between self-reported diseases and PhecodeX (Pearson’s r = 0.29, *p* = 4.27 ⨉ 10^−6^ for those with *p*_gene-burden_ < 0.0001 in both definitions; **Figure 3c**). Among burden associations significant in only one definition, self-reported-only signals were more elevated in sense organ and endocrine diseases, while PhecodeX-only signals showed higher representations in cardiovascular, musculoskeletal, and neurological diseases (**Figure 3d**), further reinforcing the value of incorporating diverse sources of phenotyping in genetic analysis.

The broad range of phenotypes also enables systematic exploration of genes and genetic variants with cross-phenotype associations, defined as statistically significant associations of a gene or variant with more than one phenotypic category, which can result from either horizontal, where genetic effects independently influence multiple phenotypes, or vertical pleiotropy, where effects on one phenotype are mediated through another. Here, we identified 44 gene-annotation pairs with cross-phenotype associations at a maxMAF of 0.1% through meta-analysis (**Figure 3e; Supplementary Data 5**). Identifying multiple phenotypes associated with a gene can often reflect coherent biological patterns, supporting known functional roles and pathways of the gene, thereby providing internal validation of association signals. For example, we observed pLoF variants in *PCSK9* influencing cholesterol-related biomarkers and clinical hyperlipidemia, as well as lipid-lowering medications, consistent with the established role of *PCSK9* in cholesterol metabolism and cardiovascular risk regulation^24,45^; and pLoF variants in *TSHR* associated with multiple phenotypes spanning hypothyroidism, circulating thyroid hormone levels, and thyroid-related treatments, reflecting the central role of *TSHR* in endocrine regulation and energy homeostasis^46^. These exemplify consistent system-wide phenotypic effects of genes and variants with their biological roles in lipid and endocrine regulation.

In addition to these concordant patterns, we identified 26 instances in which cross-phenotype associations exhibited discordant relationships between phenotypic correlation and gene effects (**Supplementary Data 6**). pLoF variants in *TET2*, frequently linked to clonal hematopoiesis of indeterminate potential (CHIP)^47^, show associations with high-density lipoprotein (HDL) levels and red blood cell (RBC) count. Despite a modest negative phenotypic correlation between these traits (r = −0.10), burden effect sizes occurred in the same direction (β_HDL_ = −5.34; β_RBC_ = −6.66). Similarly, pLoF variants in *HBB* exhibit associations with decreased hemoglobin but increased RBC count (β_hemoglobin_ = −20.02; β_RBC_ = 26.12; r = 0.75), consistent with thalassemia physiology and contrasting with iron deficiency anemia, which typically reduces both measures^48,49^.

### Biobanks provide a strong foundation for systematic genetic discovery

We harmonized association data from *All of Us* v8 and UK Biobank to build a resource of multi-phenotype GWAS and RVAS meta-analysis, spanning up to 786,871 samples and 222 harmonized phenotypes, including 42 lab measurements, 10 physical measurements, and 170 diseases. Disease phenotypes were defined using PhecodeX and Phecodes in *All of Us*, and ICD-10 and Phecodes in UK Biobank, with burden association tests performed using PhecodeX-based definitions in *All by All* v8 and ICD-10-based definitions in Genebass. To enable cross-biobank meta-analysis, we match disease phenotypes by linking *All of Us* PhecodeX to UK Biobank ICD-10 codes^7^ through pairwise phenotypic correlations with Phecodes in each biobank, retaining one-to-one matches with the highest r^2^ ⨉ r^2^ among pairs with r^2^ > 0.8, while lab and physical measurements were mapped through manual curation based on measurement definition. We then performed meta-analysis across the two biobanks using Stouffer’s method^40^ for gene-level burden tests between *All of Us* (maxMAF at 0.1%, restricting to ultra-rare variants with minimal LD with common variants while retaining sufficient alleles for association testing) and UK Biobank (only available with maxMAF at 1%) and the fixed-effect inverse-variance weighted model for single-variant association tests. We observed a substantial increase in significance for the cross-biobank associations compared to *p* values in each individual biobank (**Figure 4a**).

**Figure 4.**
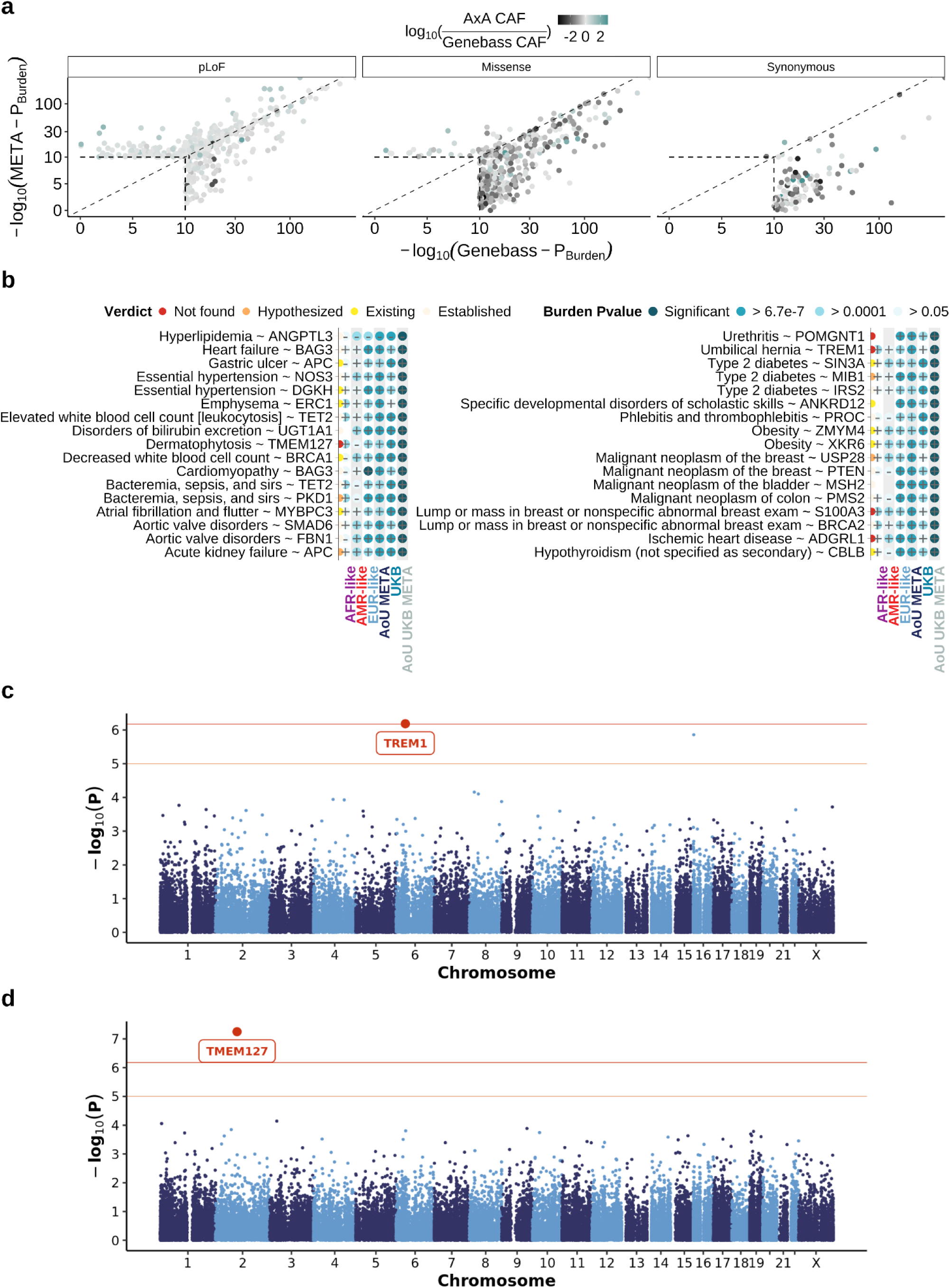
Cross-biobank analysis improves genetic discovery. **a,** Comparison of rare variant burden test significance in Genebass (UK Biobank; EUR-like only) versus meta-analysis across UK Biobank and *All of Us*. Genes with *p* < 10^−10^ in either analysis are shown. Colors indicate the ratio of the cumulative allele frequencies (CAF) of *All of Us* to UK Biobank. The dashed line denotes y=x. The scale is logarithmic for *p* value from 1 to 10^−10^, and log-log for *p* < 10^−10^. **b,** 34 pLoF rare variant burden disease associations (y-axis) that reach significance only through meta-analysis of UK Biobank and *All of Us,* but not within any individual similarity group or biobank. Red sequentially colored dots next to each association indicate AI-agent literature review verdicts (see Supplementary information), summarizing prior evidence for each association. Blue sequentially colored dots, labeled by effect direction, indicate the burden test *p value* for the corresponding cohort shown on the x-axis. **c-d,** Manhattan plots of rare variant burden test meta-analysis results across the two biobanks for **c,** umbilical hernia (GI_520.13; ICD-10: K42; N = 680,366), and **d,** dermatophytosis (DE_660.12; ICD-10: B35; N = 669,659). Each point represents aggregated pLoF variants (maxMAF at 0.001) within a single gene. Gene names for pLoF associations with *p*_gene-burden_ < 6.7 ⨉ 10^−7^ are highlighted in red.

Among 193 pLoF gene-phenotype associations that are not significant in either cohort alone (**Supplementary Data 7; Supplementary Figures 28** and **29**), 34 involve disease outcomes, and among these, we identified 5 with no or limited prior evidence in Open Targets^50^, Pubmed, and GWAS catalog^51^ (**Figure 4b**). For example, we replicated the association between rare pLoF variants in gene *MIB1* and Type 2 diabetes^52^ (*p*_cross-biobank META_ = 2.12 ⨉ 10^−7^), as well as rare pLoF variants in gene *ZMYM4* with obesity^53^ (*p*_cross-biobank META_ = 2.83 ⨉ 10^−7^). Additionally, we identify new disease relationships, such as umbilical hernia associated with gene *TREM1* (*p*_cross-biobank META_ = 6.54 ⨉ 10^−7^, **Figure 4c**), as well as dermatophytosis associated with tumor suppressor gene *TMEM127* (*p*_cross-biobank META_ = 5.65 ⨉ 10^−8^, **Figure 4d**). Our results also connect genes underlying rare diseases to common manifestations; for example, gene *ANKRD12*, previously linked to cognitive traits^7^, is nominally associated with migraine (*p*_cross-biobank META_ = 1.24 ⨉ 10^-6^).

### Data science infrastructure enables flexible large-scale gene discovery

We developed a web browser (https://allbyall.researchallofus.org/) to enable expanded exploration of the *All by All* multi-group GWAS and RVAS association results (**Extended Data Figure 3**). Designed to handle the large data volume of the results (∼100 terabytes), the platform supports efficient querying and interactive visualization across similarity groups, allowing users to compare association patterns and access summary statistics in real time. The *All by All* browser provides an interactive environment for exploring gene-level and single-variant association results through a split-screen interface that links a summary results pane with a detailed view updated by user selection. This browser facilitates efficient comparison of associations across phenotypes, tests, and genomic regions, providing a flexible and open environment that lowers barriers to data access.

In addition to initial data exploration in the browser, researchers can further query the *All by All* data directly from the *All of Us* Researcher Workbench and integrate these results with other types of data in *All of Us* for deeper biological interpretations. Together, these resources empower researchers worldwide to investigate the functional effects of genetic variants on complex diseases with improved resolution and reproducibility.

## Discussion

We present extensive GWAS and RVAS results from the *All by All* project using *All of Us* data to understand the genetic basis of human traits and diseases through both common and rare variants, advancing gene discoveries as well as the development of therapeutic approaches. This serves as a first release of what we envision as an ongoing process of analyzing data with millions of individuals, which will facilitate increasingly powerful association tests for lower-prevalence diseases (e.g., <1%). By including genetically diverse populations, the *All by All* resources improve human genetic research by identifying novel associations attributable to allele frequency differences, evaluating the consistency of effects, and enabling the future identification of putatively causal variants through differences in linkage disequilibrium structure across populations. Given the consistency of genetic effects across populations, it also provides a valuable resource for replication of associations across a broader range of individuals while addressing genomic knowledge gaps arising from the underrepresentation of populations in existing genomic datasets^54,55^.

While *All by All* provides valuable insights, several potential limitations should be acknowledged. First, as a genetic association resource, the associations identified through *All by All* analyses represent statistical correlations between genetic variation and phenotype rather than necessarily identifying causal variants, as observed signals may also reflect linkage disequilibrium, residual population stratification, or other indirect factors. Because the *All by All* analyses are limited to single-gene or single-variant associations, they capture only part of the liability underlying complex traits, which may arise from combined genetic and non-genetic effects. Their principal value, therefore, lies in genetic discovery, whereas translation to therapeutic benefit will require further work to establish mechanisms of genetic effects.

Second, the current sample sizes result in limited power for detecting associations with lower prevalence outcomes, which will improve as the *All of Us* Research Program continues to grow and additional biobanks generate sequencing data. Cross-biobank meta-analysis between the UK Biobank and *All of Us* Research Program in our study has substantially increased statistical power and discovery, but residual heterogeneity introduced by phenotype harmonization across coding systems, healthcare settings, and cohort demographics and structure may still influence inference. Differential participation is an important component of such heterogeneity, for which healthy volunteer bias and survival bias differ remarkably between cohorts, with UK Biobank participants tending to be healthier and older than the underlying UK population, whereas the *All of Us* Research Program intentionally recruits individuals from populations historically underrepresented in biomedical research and includes participants with greater clinical complexity. Moreover, because the UK Biobank is composed predominantly of individuals of European ancestry, the meta-analysis also disproportionately reflects European-specific signals relative to analyses of *All of Us* alone. Thus, associations described as replicated through cross-biobank meta-analysis should be interpreted as replication in a primarily European cohort. A further caveat is that, for practical considerations, different minor allele frequency thresholds were applied for the gene-level rare variant burden test in the two biobanks for meta-analysis: 0.1% for *All of U*s and 1% for UK Biobank. As a result, cross-biobank meta-analyzed burden tests may aggregate partially non-overlapping variant sets across cohorts, and statistics from Stouffer’s test may reflect partly distinct genetic signals. These limitations should be considered when interpreting the cross-biobank findings.

We ensured robustness of our results via a meta-analysis approach in which individuals were assigned to discrete genetic similarity groups, despite the continuous nature of genetic ancestry and the frequent presence of admixture across populations. This framework reduces false positive associations due to population stratification, but modestly reduces statistical power, particularly when admixed individuals are excluded by ancestry group assignment thresholds or when smaller groups are excluded due to limited effective sample sizes, particularly for rarer outcomes. Continued development of scalable methods that incorporate finer-scale population structure, local ancestry, admixture, and heterogeneous effective sample size will therefore be important for improving inference in increasingly large and diverse cohorts.

We further recognize that socially defined ancestry and ethnic categories have historically been misused to justify claims about innate biological differences, contributing to harm across populations. The genetic similarity groupings used in this study aid in controlling type I error, but are imperfect proxies for patterns of genetic variation shaped by population history and should not be interpreted as biologically discrete categories or as equivalent to socially defined race or ethnicity. Group labels such as AFR-like and EUR-like reflect genetic similarity to reference panels, not membership in a population. Readers and downstream users of these summary statistics should interpret group-stratified results with these analytic origins in mind, particularly when considering implications for clinical translation or polygenic score development.

Furthermore, there is a caveat in the arbitrary definition of cutoffs for common variants in the ACAF dataset, which is population-specific allele frequency (AF) > 1% or population-specific allele count (AC) > 100 in any computed similarity groups. This cutoff should not be construed as an implication that all variants above this threshold are not functional and those below are: rather, this cutoff attempts to balance functional consequence with frequency to maximize power. However, such a cutoff can have differential power to detect associations across groups with varying sample sizes or across genes with different distributions of functional consequences.

Finally, our analysis of EHR-based phenotypes was limited to phecodeX or ATC definitions derived from billing and diagnosis codes across multiple healthcare systems. These phenotypes reflect not only underlying biology but also healthcare access, treatment patterns, social and structural inequities, and environmental and behavioral factors that influence diagnosis, treatment, and disease course, as well as differential participation and measurement heterogeneity across healthcare settings. Although we attempted to control for major sources of confounding, some bias may still remain in the association effect size estimates, including from residual stratification of environmental exposures and other non-genetic factors that may intersect with genetic ancestry, gene-environment interactions, or violations of association model assumptions. For most individual associations, we expect these biases to contribute only modestly to estimated effect sizes. In aggregate, however, such as when summing genetic effects to calculate polygenic scores, these biases may compound and limit interpretability and transferability across populations. The present findings should therefore be interpreted as one component of a broader context shaping disease risk and progression across populations. Incorporation of richer phenotypic representations, including longitudinal clinical profiles and life-course exposures, will substantially enhance clinical resolution required to clarify disease trajectories, mechanistic pathways, and etiology.

The analysis of the full GWAS and gene-level RVAS results from our *All by All* study demonstrates the value of genetic association analysis across common and rare variants across a range of diseases, as well as across diverse genetic backgrounds. Additionally, the public release of this resource provides valuable opportunities for researchers around the world to more efficiently identify gene targets and interpret disease pathways. Our framework strongly increases non-European representation, enabling the identification and contextualization of genetic associations in underrepresented populations, while also highlighting the need for continued expansion of diverse cohorts for more balanced and globally representative investigations into the mechanisms of diseases and supporting scientific advances that are broadly applicable for all.

## Extended Data Figures/Tables

**Extended Data Table 1.**
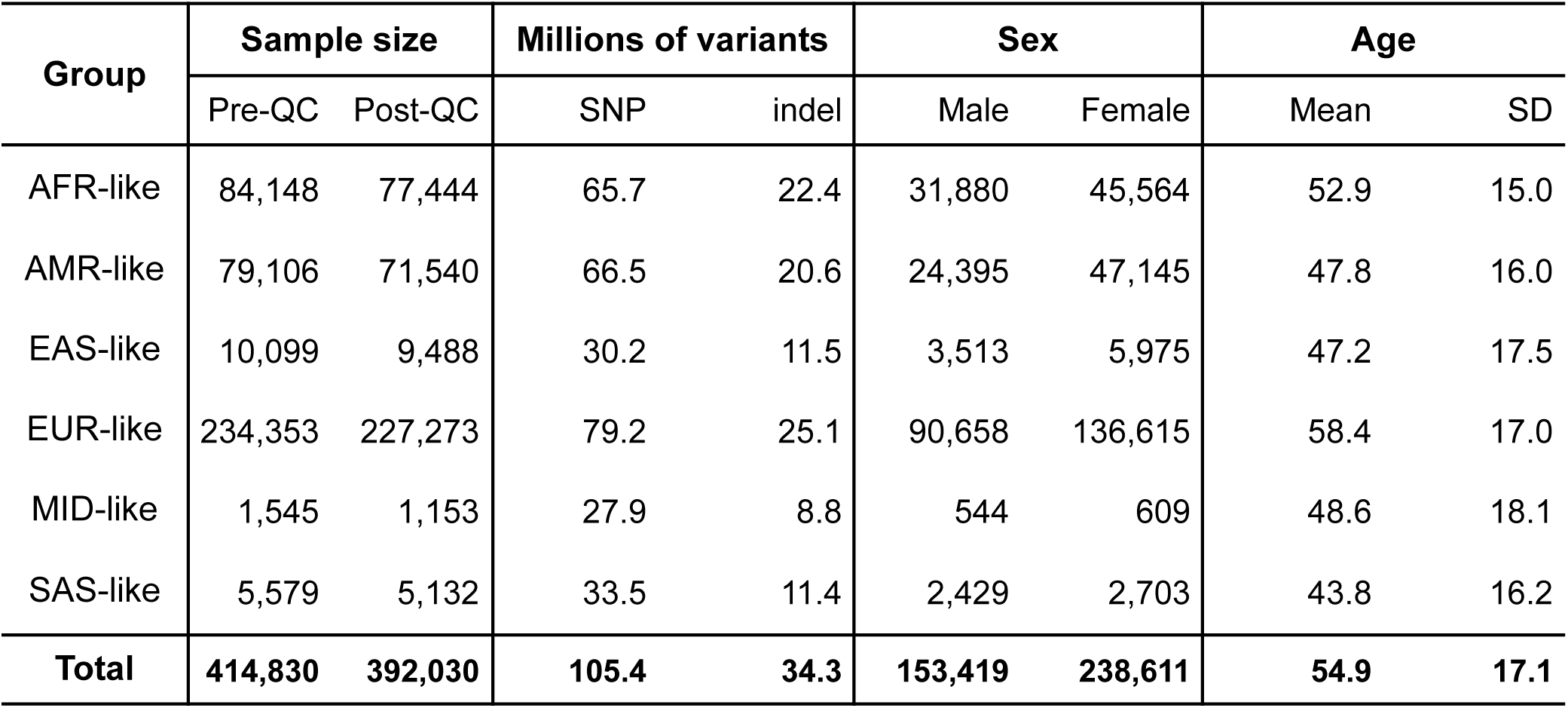
*All by All* sample data summary of study cohort by genetic similarity group. Sample sizes before and after quality control (QC), number of variants (SNPs and indels, in millions), sex distribution, and age distribution (mean ± SD) are shown for each group. Similarity groups were initially assigned based on genetic similarity to reference populations and further refined based on centroid distances from group-specific principal component analysis (PCA): AFR-like, AMR-like, EAS-like, EUR-like, MID-like, and SAS-like (Supplementary Information). All metrics except sample size are summarized using post-QC data.

| Group | Sample size |  | Millions of variants |  | Sex |  | Age |  |
| --- | --- | --- | --- | --- | --- | --- | --- | --- |
|  | Pre-QC | Post-QC | SNP | indel | Male | Female | Mean | SD |
| AFR-like | 84,148 | 77,444 | 65.7 | 22.4 | 31,880 | 45,564 | 52.9 | 15.0 |
| AMR-like | 79,106 | 71,540 | 66.5 | 20.6 | 24,395 | 47,145 | 47.8 | 16.0 |
| EAS-like | 10,099 | 9,488 | 30.2 | 11.5 | 3,513 | 5,975 | 47.2 | 17.5 |
| EUR-like | 234,353 | 227,273 | 79.2 | 25.1 | 90,658 | 136,615 | 58.4 | 17.0 |
| MID-like | 1,545 | 1,153 | 27.9 | 8.8 | 544 | 609 | 48.6 | 18.1 |
| SAS-like | 5,579 | 5,132 | 33.5 | 11.4 | 2,429 | 2,703 | 43.8 | 16.2 |
| **Total** | **414,830** | **392,030** | **105.4** | **34.3** | **153,419** | **238,611** | **54.9** | **17.1** |

**Extended Data Figure 1.**
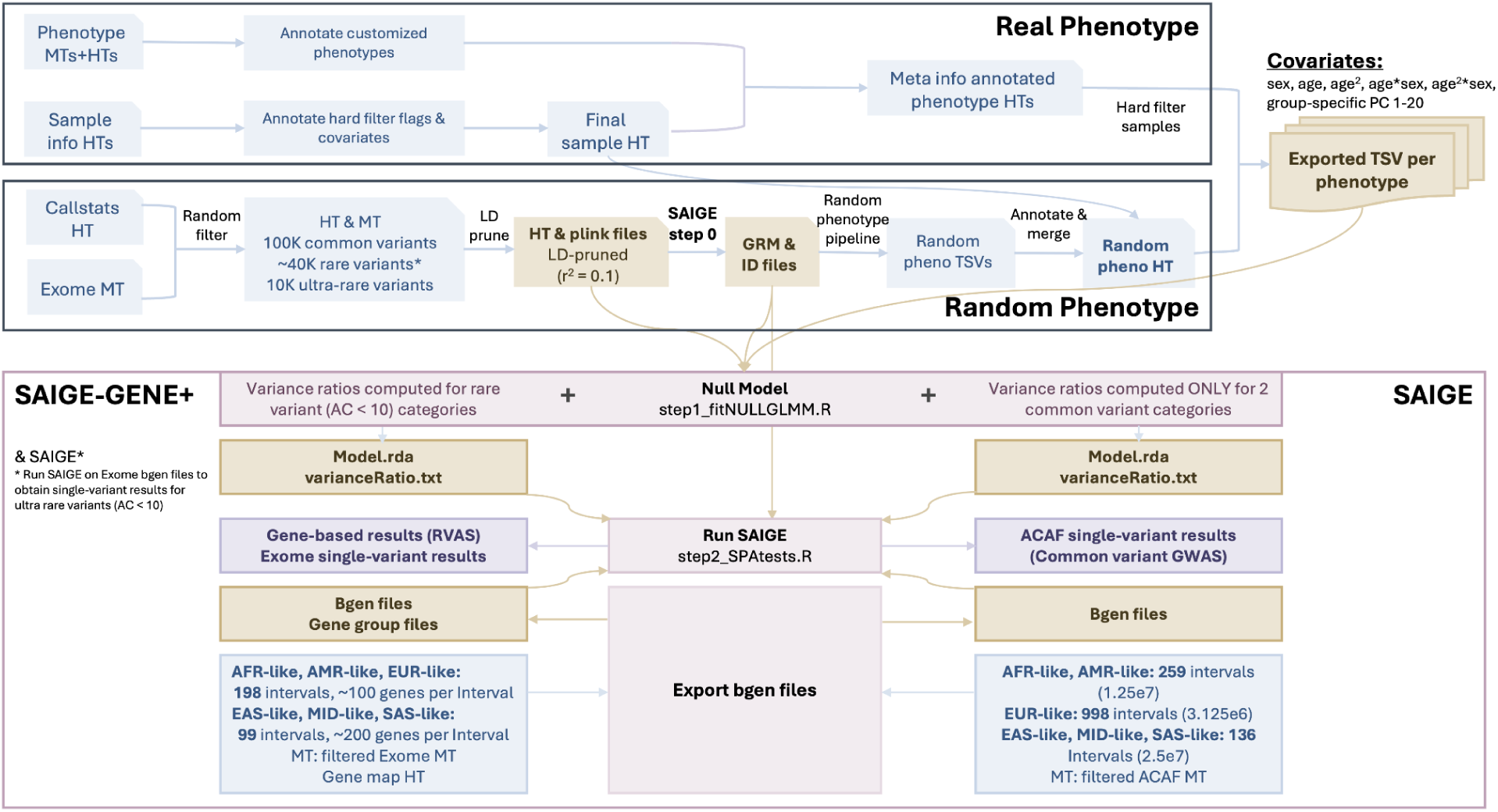
*All by All* computational framework. Quality control (QC) is performed on phenotype and sample data, and phenotype files are prepared for samples post-QC per genetic similarity group, including covariates (age, sex, age^2^, age ⨉ sex, age^2^ ⨉ sex, and group-specific principal components 1–20). SAIGE step 0 computes sparse genetic relationship matrices (GRMs) using LD-pruned common variants from the Exome Hail MatrixTable (MT) in *All of Us*. PLINK files are generated with variants across the allele-frequency spectrum for both GRMs and association testing. Using these GRMs, random phenotypes with varying prevalences are simulated for calibration. SAIGE step 1 fits null generalized linear mixed models for real and simulated phenotypes using phenotype data, covariates, GRMs, and a subset of genotype data from the PLINK files. Separate null models are computed for ACAF (Allele Count/Allele Frequency threshold callset in *All of Us*) and Exome callsets to incorporate variance-ratio estimates for ultra-rare variants, and genotype data are exported from the ACAF and Exome MTs as group-specific BGEN intervals. SAIGE and SAIGE-GENE+ step 2 perform single-variant and set-based association testing (burden, SKAT, and SKAT-O) for all phenotypes passing QC across six genetic similarity groups.

**Extended Data Figure 2.**
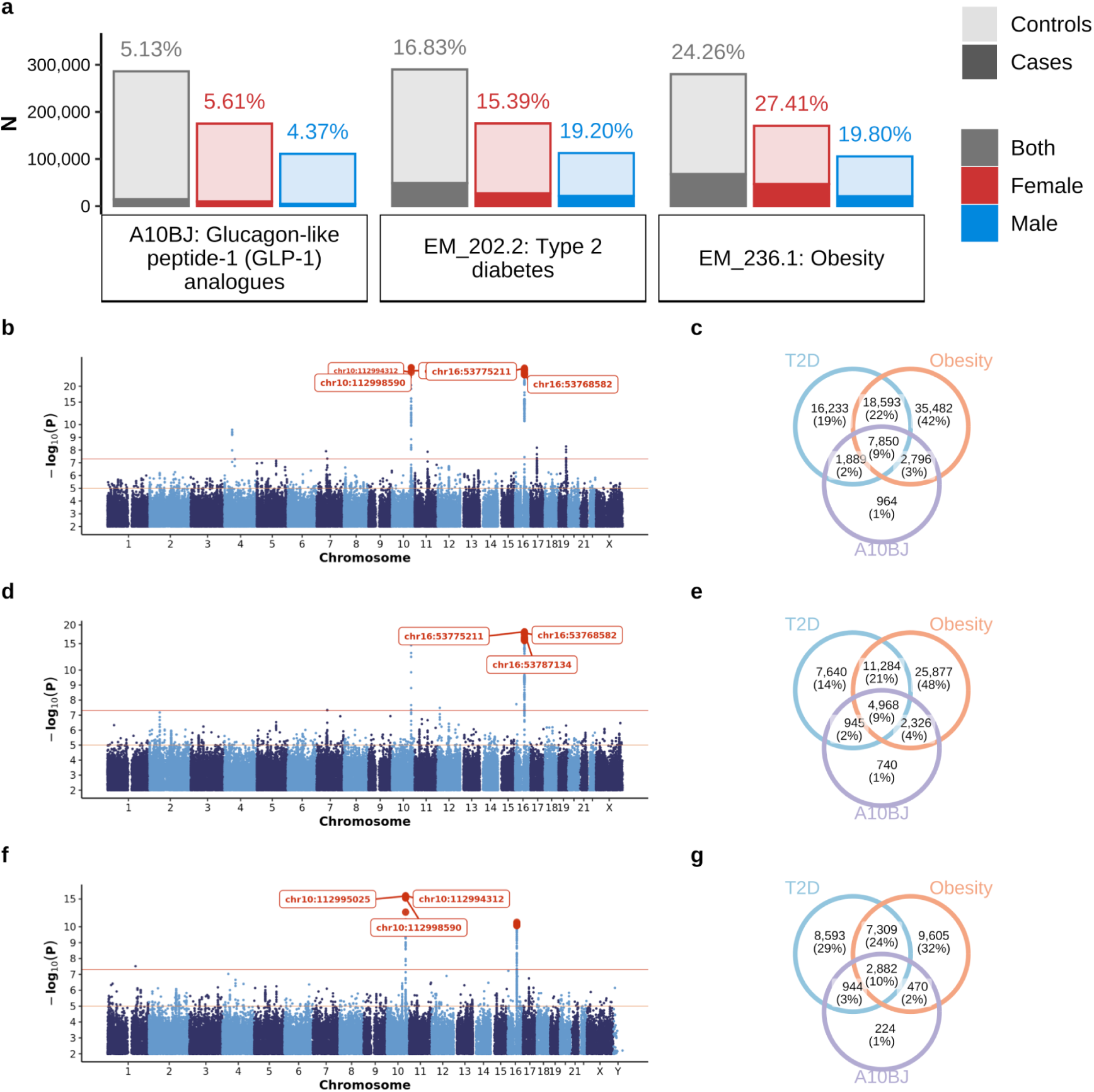
Sex-stratified analysis of GLP-1 analog usage and associated medical conditions. **a**, Comparison of prevalence and sample sizes across sex groups for type 2 diabetes (PhecodeX: EM_202.2), obesity (PhecodeX: EM_236.1), and glucagon-like peptide-1 (GLP-1) analog exposure (ATC: A10BJ). Bar height indicates sample size for the corresponding phenotype (x-axis) and sex group (color). Numbers above bars indicate prevalence (%) for each sex group and phenotype. Transparency distinguishes cases and controls. **b,** Manhattan plots of *p* values from meta-analysis of common variant associations for GLP-1 analog exposure (A10BJ; both sexes: n_cases_= 14,682; n_controls_= 271,438)**. c**, Venn diagram showing overlap among individuals diagnosed with type 2 diabetes, individuals diagnosed with obesity, and individuals taking GLP-1 analog medications. Labels indicate the number of individuals belonging uniquely to one circle or jointly to two or all three circles. **d** and **e** present the same information as **b** and **c**, respectively, for females only (n_cases_= 9,828; n_controls_= 165,308). **f** and **g** present the same information as **b** and **c**, for males only (n_cases_= 4,854; n_controls_= 106,130).

**Extended Data Figure 3.**
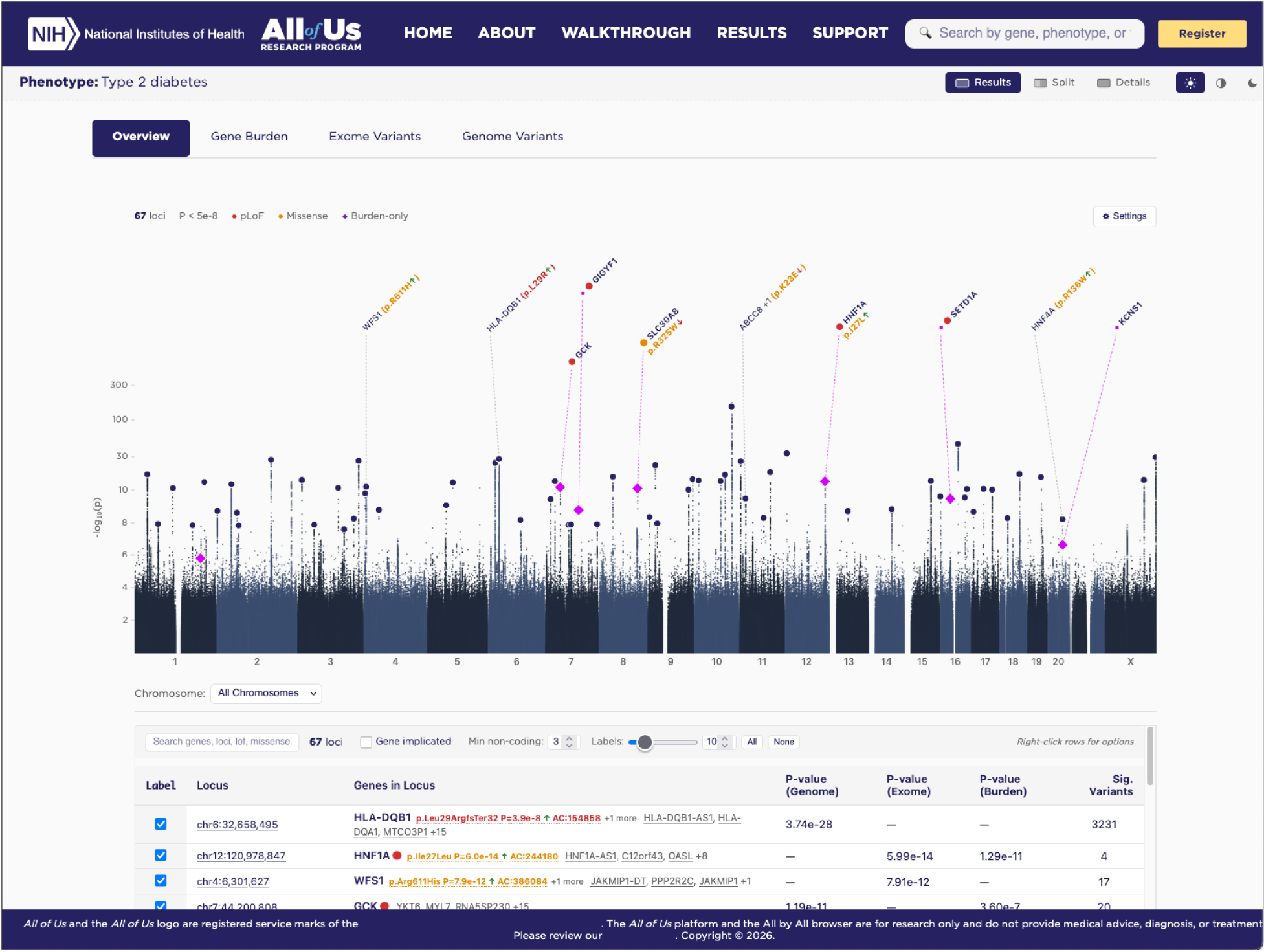
*All by All* browser. Manhattan, QQ, and PheWAS plots serve as primary visual summaries, enabling rapid scanning of genome-wide or phenome-wide patterns and identification of significant or near-threshold signals. Plot points and table rows are clickable, revealing gene-, variant-, or phenotype-specific pages that include genomic coordinate variant tracks, burden test results, and regional views integrating single-variant and gene-level signals within a locus. Filters for mutation class, consequence category, phenotype attributes, allele-frequency ranges, and association test type support targeted exploration, while sortable tables and tooltips aid the interpretation of effect estimates and quality metrics. The interface emphasizes fluid movement between overview and detail: users can resize or hide panels, toggle between plot types, adjust *p* value scaling, or navigate from genome-wide views to PheWAS or locus-level displays without losing context.

## Data availability

All by All Github repository: https://github.com/atgu/aou_gwas

All by All browser: https://allbyall.researchallofus.org/

## Acknowledgements

We gratefully acknowledge *All of Us* participants for their contributions, without whom this research would not have been possible. We also thank the National Institutes of Health’s All of Us Research Program for making available the participant data, samples, and cohorts examined in this study.

## All of Us Research Program - Consortium Contributors

**NIH All of Us Research Program Staff:** Joshua C. Denny^2^, Tara Dutka^2^, Jennifer Adjemian^2^, Naomi Aspaas^2^, Romuladus E. Azuine ^2^, Brian Bataille^2^, Katherine D. Blizinsky^2^, Subhashini Chandrasekharan^2^, Toral Contractor^2^, Alyssa Cotler^2^, Stephanie Devaney^2^, Holly Garriock^2^, Geoffrey S. Ginsburg^2^, Michael J. Hahn^2^, Mary Grace Hébert^2^, Justin Hentges^2^, Katharine Nichole Holm^2^, Sonya Jooma^2^, Minnkyong Lee^2^, Chris Lunt^2^, Ryan Lupi^2^, James McClain^2^, Sydney McMaster^2^, Martin Mendoza^2^, Anjene Musick^2^, Bintu Ojehomon^2^, Olanike T. Oladipo^2^, Brad Ozenberger^2^, Rachele Peterson^2^, Andrea Ramirez^2^, Jessica Reusch^2^, Prachi Patel^2^, Janeth Sanchez^2^, Violeta Seidell^2^, Sheri D. Schully^2^, Katrina I. Theisz^2^, Karriem Watson^2^

**Steering Committee:** Habibul Ahsan^12^, Hoda Anton-Culver^13^, Deborah Banks^3^, Melissa A. Basford^1^, Randee Bloom^3^, Hugo Campos^3^, Mine S. Cicek^15^, Elizabeth Cohn^17^, Stephanie H Cook^18^, Linda Cottler^19^, Kathy Deerinwater^3^, Joshua C. Denny^2^, Stephanie Devaney^2^, Anjali Forber-Pratt^20^, Stacey Gabriel^10^, Holly Garriock^2^, Geoffrey S. Ginsburg^2^, David Glazer^21^, Melissa Haendel^23^, Paul Harris^11^, Scott Joseph Hebbring^24^, Justin Hentges^2^, Beverly Holmes^25^, Gail P Jarvik^9^, Jason Karnes^26^, Winston Koo^3^, Bruce Korf^27^, Monica Kraft^28^, Minnkyong Lee^2^, Kate LeFauve^39^, KiTani Lemieux^30^, Mitchell R. Lunn^31^, April Brinkoetter McCrea^33^, James McClain^2^, Debra Dianne Murray^7^, Saer Oan^3^, Lucila Ohno-Machado^34^, Akinlolu Ojo^35^, Cathryn Peltz-Rauchman^36^, Rachele Peterson^2^, Steven Reis^6^, Edgar M Gil Rico^37^, Grisel Marie Robles-Schrader^38^, Linda Salgin^60^, Janeth Sanchez^2^, Suaih Morales^3^, Sheri D. Schully^2^, Edward Schlicksup^69^, Stephen Smith^39^, Jordan Smoller^40^, Andreas Theodorou^26^, Cheryl Thomas^41^, Eric Topol^42^, Phil Tsao^43^, Jen Uhrig^70^, Julia Moore Vogel^42^, Eboni Winford^44^, Katrina Yamazaki^45^, Stephan Zuchner^46^

**The Broad-LMM-Color Genome Center:** <u>Stacey Gabriel^10^, Niall Lennon^1^</u>^0^, <u>Heidi L. Rehm^10^,</u> <u>Scott Topper</u>^1^, Namrata Gupta^10^, Konrad J. Karczewski^10^, Lee Lichtenstein^10^, Wenhan Lu^10^, Jeremy Guez^10^, Riley Grant^10^, Matt Solomonson^10^, Ben Neale^10^

**Baylor Hopkins Center for Clinical Genomics:** Richard A. Gibbs^7^, Kimberly F. Doheny^5^, Eric Boerwinkle^4^, Ginger A. Metcalf^7^, Donna M. Muzny^7^, Eric Venner^7^

**Northwest Genomics Center:** <u>Gail P. Jarvik^9^, Evan E. Eichler^9^, Chia-Lin We</u>i^9^, Phillip E. Empey^6^, Christian Frazar^9^, Tina Lockwood^9^, Brian Shirts^9^, Joshua D. Smith^9^, Colleen Davis^9^

**The Biobank:** <u>Mine S. Cicek^1^</u>^5^, Tony G. Bilyeu^15^, Ashley L. Blegen^15^, Jolene J. Lane^15^, Jeffrey G. Meyer^15^

**The Data and Research Center:** <u>Paul A. Harris^11^, Melissa A. Basford^11^, Dan M. Roden^1^</u>^1^, Emmanuel Ankumah-Saikoom^1^, Robert J. Carroll^11^, Justin A. Cook^11^, Megan K. He^11^, Aymone Jeanne S. Kouame^11^, Lee Lichtenstein^10^, Jodell E. Linder^11^, Michael Lyons^11^, Brandy M. Mapes^11^, Kayla Marginean^11^, Megan Morris^11^, Jun Qian^11^, Ryan Samarakoon^11^, Samantha Stewart^11^, Chun-I Tang^11^, Yuanyuan Wang^11^, Consuelo H. Wilkins^11^

**The Participant Center (including Scripps Research Translational Institute, CareEvolution and Sage Bionetworks):** Sasri Dedigama^42^, Megan Doerr^67^, Vik Kheterpal^68^, Christine Suver^67^, Ashley Tate^68^, Julia Moore Vogel^42^, John Wilbanks^67^

**The Participant Technology Systems Center:** <u>Mark Begale^49^, Scott Sutherland</u>^49^, Jeff Drommerhausen^49^, John Durham^49^, Catherine E. W. Freeland^49^, Cecilia Gacek^49^, Gail Hamilton^49^, Aditya Naik^49^, Julie Shaw^49^, Daniel Uribe^49^

**Acknowledging former members of the Steering Committee**: Ivye L. Allen^3^, Katie Baca-Motes^42^, Laura Bartlett^47^, Debra Bartley-Rice^48^, Mark Begale^49^, Eric Boerwinkle^4^, Murray H Brilliant^24^, Jessica Burke^14^, Patricia Butts^3^, Michael Castro^2^, John Chaffins^33^, Katherine Chang^3^, Veena Channamsetty^50^, Carmen Chinea^51^, Cheryl Clark^16^,Vivian Colon-Lopez^52^, Toral Contractor^2^, Karl Cooper^20^, Alyssa Cotler^2^, Carolyn Cronin^3^, Martha Daviglus^53^, Liliana Lombardi Desa^3^, Eric Dishman^2^, Charles E. Drum^20^, Mark Edmunds^54^, Dejen M. Eshete^3^, Adolph Falcón^37^, Miguel Flores^3^, Gretchen Funk^55^, Kelly Gebo^2^, Ali G. Gharavi^56^, David B. Goldstein^56^, Joannie Grand^14^, Philip Greenland^22^, Kristin Hall^57^, Kimiyo Harris-Williams^3^, Edina Harsay^3^, Richard Hochfelder^3^, Nazia Hussain^3^, Praduman Jain^49^, Gwynne Jenkins^2^, Ayanna Jenkins^48^, Christine C. Johnson^36^, Anna Johnston^3^, Parinda Khatri^44^, Linda Perez Laras^29^, Patricia Watkins Lattimore^41^, Brendan Lee^7^, Dessie Levy^3^, Megan Lewis^3^, Chris Lunt^2^, Emily Makahi^32^, Patrick McGovern^33^, Michelle McNeely^2^, Martin Mendoza^58^, Wanda Montalvo^59^, Francisco Moreno^26^, Fatima Munoz^60^, Narayana Murali^61^, Christopher O’Donnell^43^, May Okihiro^62^, Greg Orr^57^, Andrea Ramirez^11^, Fornessa Randal^63^, Dara Richardson-Heron^2^, Dan Roden^11^, Joni Rutter^2^, Eric Schlueter^25^, Yashoda Sharma^28^, Hannah Sinemus^6^, Vicki Smith^42^, Teshia Solomon^26^, Louisa Stark^64^, Steven Steinhubl^42^, Christine Suver^65^, Gregory Talavera^60^, Amy Taylor^50^, Ronnie Tepp^66^, Steve Thibodeau^15^, Scott Topper^1^, Vanessa Uzoh^3^, Karriem Watson^2^, David Wellis^54^, Consuelo Wilkins^11^, Amanda Wilson^47^, Joyce Winkler^3^, Alicia Y. Zhou^1^

**Acknowledging in memoriam:** Stephen Mikita^3^, Deborah A Nickerson^9^, Lauren Ryan^1^, Karl Surkan^3^

**Affiliations:** ^1^Color Health; ^2^All of Us Research Program / National Institutes of Health; ^3^Participant Steering Committee Member; ^4^The University of Texas Health Science Center at Houston; ^5^Johns Hopkins University School of Medicine; ^6^University of Pittsburgh; ^7^Baylor College of Medicine; ^8^Laboratory for Molecular Medicine; ^9^University of Washington; ^10^Broad Institute; ^11^Vanderbilt University Medical Center; ^12^University of Chicago; ^13^University of California, Irvine; ^14^The MITRE Corporation; ^15^Mayo Clinic; ^16^Brigham and Women’s Hospital; ^17^The Feinstein Institutes for Medical Research; ^18^New York University; ^19^University of Florida; ^20^American Association on Health and Disability; ^21^Verily Life Sciences; ^22^Northwestern University; ^23^University of North Carolina School of Medicine; ^24^Marshfield Clinic Research Institute; ^25^Cooperative Health; ^26^University of Arizona; ^27^University of Alabama at Birmingham; ^28^Icahn School of Medicine at Mount Sinai; ^29^San Juan Bautista School of Medicine; ^30^Xavier University; ^31^Stanford University School of Medicine; ^32^Waianae Coast Comprehensive Health Center; ^33^HungryHeart Media, Inc. (Wondros); ^34^Yale School of Medicine; ^35^University of Kansas Medical Center; ^36^Henry Ford Health; ^37^National Alliance for Hispanic Health; ^38^Pyxis Partners; ^39^NORC; ^40^Partners Healthcare System; ^41^Delta Research and Educational Foundation; ^42^Scripps Research Translational Institute; ^43^U.S. Department of Veterans Affairs; ^44^Cherokee Health Systems; ^45^California State University Los Angeles; ^46^University of Miami School of Medicine; ^47^National Library of Medicine; ^48^Jackson-Hinds Comprehensive Health Center; ^49^Vibrent Health; ^50^Community Health Center; ^51^HRHCare; ^52^University of Puerto Rico Comprehensive Cancer Center; ^53^University of Illinois at Chicago; ^54^San Diego Blood Bank; ^55^FiftyForward; ^56^Columbia University Irving Medical Center; ^57^Walgreens; ^58^National Institutes of Health; ^59^University of Pennsylvania; ^60^San Ysidro Health; ^61^Geisinger Health System; ^62^University of Hawai’i; ^63^Asian Health Coalition; ^64^University of Utah School of Medicine; ^65^Emory University; ^66^HCM Strategists LLC; ^67^Sage Bionetworks; ^68^CareEvolution; ^69^IBM; ^70^RTI

**Note:** Underlining indicates the grant’s Principal Investigators

