## supplementary_information for "Systematic common and rare variant association testing in 392,030 whole genomes in *All of Us*"

### AxA Supplementary Information

AxA Supplementary Information 1
All of Us data description 7
Genotype Data 7
Phenotype Data 8
Supplementary Figure 1 | Cross-biobank consistency of phenotype prevalence. Comparison of Phecode prevalence between UK Biobank (x-axis) and All of Us v8 (y-axis) across genetic similarity groups (panels and colors). Each point represents a Phecode. The dashed lines represent y=x. 10
Supplementary Table 1 | Cross-biobank concordance of Phecode sample size and prevalence. Pearson's correlation coefficient (r) and corresponding p values for the number of cases, the number of controls, and prevalence for Phecodes in UK Biobank and All of Us v8 across similarity groups. 10
Pre-GWAS QC 11
Genotype QC 11
Sample QC 11
Pruning similarity group outliers and group-specific PCA 12
Supplementary Figure 2 | Sample QC and phenotype QC pipeline. The left column describes sample information obtained from the All of Us Researcher Workbench and filtering metrics used for sample QC. The right column describes phenotype categories and the selection criteria for phenotypes included in association testing. The middle column bar charts show the number of samples and phenotypes across similarity groups and phenotypic categories after applying sample and phenotype QC filters and then restricting to phenotypes with a group-specific number of cases ≥ 200. The bottom matrix shows the number of phenotypes (cells) within each phenotype category (y-axis) stratified by the number of genetic similarity groups sharing each phenotype (x-axis). Bars to the right of the matrix indicate total phenotypes analyzed per category. 14
Supplementary Figure 3 | PC-based centroid outlier pruning pipeline. Starting from the genotype and sample-QCed ACAF callset, samples were separated by relatedness. PCA was performed on unrelated samples and then computed for related samples by projecting the related genotypes on the unrelated PC space. Using these PC scores, centroid outliers were removed, and PCA was recomputed on the pruned set using the same procedure as described earlier for the first round. 14
Supplementary Figure 4 | PCA in All of Us v8 participants assigned to AFR-like and corresponding centroid distance across 3 PCs. Centroid distance distributions and PC biplots for the first 6 PCs are shown before (top) and after (bottom) pruning outliers. The vertical line in the two left centroid distance histograms shows the threshold chosen to remove outliers. 15
Supplementary Figure 5 | PCA in All of Us v8 participants assigned to AMR-like and corresponding centroid distance across 3 PCs. Centroid distance distributions and PC biplots for the first 6 PCs are shown before (top) and after (bottom) pruning outliers. The vertical line in the two left centroid distance histograms shows the threshold chosen to remove outliers. 15
Supplementary Figure 6 | PCA in All of Us v8 participants assigned to EAS-like

and corresponding centroid distance across 3 PCs. Centroid distance distributions and PC biplots for the first 6 PCs are shown before (top) and after (bottom) pruning outliers. The vertical line in the two left centroid distance histograms shows the threshold chosen to remove outliers. 16
Supplementary Figure 7 | PCA in All of Us v8 participants assigned to EUR-like and corresponding centroid distance across 5 PCs. Centroid distance distributions and PC biplots for the first 6 PCs are shown before (top) and after (bottom) pruning outliers. The vertical line in the two left centroid distance histograms shows the threshold chosen to remove outliers. 16
Supplementary Figure 8 | PCA in All of Us v8 participants assigned to MID-like and corresponding centroid distance across 5 PCs. Centroid distance distributions and PC biplots for the first 6 PCs are shown before (top) and after (bottom) pruning outliers. The vertical line in the two left centroid distance histograms shows the threshold chosen to remove outliers. 17
Supplementary Figure 9 | PCA in All of Us v8 participants assigned to SAS-like and corresponding centroid distance across 3 PCs. Centroid distance distributions and PC biplots for the first 6 PCs are shown before (top) and after (bottom) pruning outliers. The vertical line in the two left centroid distance histograms shows the threshold chosen to remove outliers. 17
Supplementary Table 2 | Sample and variant kept across QC stages by genetic similarity groups. Number of samples shown after hard filtering (post initial QC) and centroid pruning (pruned), along with variant counts before downsampling and after LD pruning. 18
Phenotype curation 18
Supplementary Table 3 | Phenotypes kept after sample QC and phenotype selection by phenotype category and genetic similarity group. Values subtracted from the total column on the right represent rare random phenotypes lacking defined cases in the three smallest similarity groups and therefore excluded due to null model non-convergence in SAIGE step1. (Note: random phenotypes span prespecified prevalence ranges for result validation, and are not subject to the case count filters applied to other phenotypes). 19
Association testing 19
SAIGE 20
SAIGE-GENE+ 21
Meta-analysis 22
Meta-analysis across the UK Biobank and All of Us 24
Supplementary Table 4 | Cross-biobank phenotype mapping overview. Summary of phenotypes mapped between All of Us and UK Biobank, with counts stratified by broad phenotypic classes and disease categories. 25
Meta-analysis vs. Mega-analysis 25
Supplementary Figure 10 | Meta- v.s. mega-analysis concordance across phenotypes. Manhattan and quantile-quantile (QQ) plot comparison between meta-analysis and mega-analysis across selected phenotypes. Each point represents a common variant from the All of Us ACAF callset. In the Manhattan plots, the x-axis shows genomic position, and the y-axis shows -log10(p). Chromosome and position for variants with $p < 5 \times 10$-8 are highlighted in red. In the QQ-plots, expected versus observed -log10(p) are shown, with expectation

based on the 2 distribution. The red line indicates y=x and GC is displayed in the upper left. A-B, Simulated quantitative phenotype with index 1: mega-analysis (N = 407,490) and meta-analysis (N = 392,030). C-D, Type II Diabetes (PhecodeX: EM_202.2): mega-analysis (N = 302,693; Ncases=51,058, Ncontrols=251,635) and meta-analysis (N = 290,052; Ncases= 48,828, Ncontrols=241,224). E-F, Triglycerides (3022192): mega-analysis (N = 148,030) and meta-analysis (N = 143,386). 26

Computational framework 27

Supplementary Table 5 | Association testing workload summary. Total number of association tests performed and jobs executed, stratified by result type across genetic similarity groups and meta-analysis. 28

Supplementary Figure 11 | Computational cost and runtime of SAIGE-based association pipelines. Distribution of per-phenotype computational cost (USD; top row) and CPU time (minutes; bottom row) for SAIGE and SAIGE-GENE+ Step 1 and Step 2 across genetic similarity groups. Columns denote association analyses (GWAS, ExWAS, and gene-level burden tests). Similarity groups are shown on the x-axis and by color. Total USDs and CPU hours across similarity groups are labeled on the top left and bottom left of the corresponding panels, respectively. 28

Result Format 28

Supplementary Figure 12 | Final result outputs and meta-analysis MatrixTable computation workflow. The upper panel shows the number of phenotypes analyzed for each genetic similarity group and in the meta-analysis. The lower panel illustrates the workflow for aggregating group-specific Hail MatrixTables and generating meta-analysis results. 30

Supplementary Table 6 | Hail data sizes and association summary. The upper table reports sizes of Hail Matrix Tables (MTs) and Hail Tables (HTs), and the lower table reports numbers of association statistics, both stratified by genetic similarity groups and association types, along with counts for meta-analysis and overall total. 30

QC of summary statistics 30

Independent phenotypes 30

Supplementary Figure 13 | Phenotype independence assessment. A, Histogram of phenotype pair counts (y-axis) by squared correlation (r2; x-axis). B, Number of phenotypes removed (y-axis, and labeled on top of each bar) using the maximum independent set method across r2 threshold (x-axis). 31

Simulated random phenotypes 31

Supplementary Figure 14 | Pipeline for generating group-specific PLINK files and sparse genetic GRMs for SAIGE step 1, with downstream random phenotype simulation. Genotype- and sample-QCed data from the Exome MT are used to select group-stratified autosomal variants across allele frequency bins, followed by LD pruning (r2 = 0.1). GRMs are then computed and used to simulate random phenotypes with varying prevalence via the R package sparseMVN. 32

Supplementary Figure 15 | Quantile–quantile (QQ) plots of gene-based burden association results for simulated phenotypes (maxMAF = 0.001). Each panel corresponds to a prespecified phenotype prevalence used in the simulations. Points represent gene–annotation pairs, and are colored by genetic similarity group. The x-axis shows expected $-\log_{10}$(p values) under the $\chi^2$ distribution, and

the y-axis shows observed $-\log_{10}$(p values) from burden tests. The dashed line denotes the null expectation (y = x). Colored numbers in the upper left of each panel indicate the genomic inflation control factor (GC) for the corresponding prevalence with text color matching the similarity group. 33
Empirical p value threshold 34
Supplementary Figure 16 | Distribution of empirical p value significance thresholds. Each point represents a specific combination of association test type (GWAS, ExWAS, burden, SKAT, or SKATO; x-axis) and genetic similarity group (color). The y-axis shows negative log10 of the empirical p value thresholds estimated using random phenotypes from All of Us. The horizontal dashed line indicate the threshold computed from Genebass random phenotypes, with test types labeled at the upper right of each line. 34
Consistency of variant effect sizes on height across cohorts 35
Supplementary Table 7 | GWAS height associations identified in All of Us v8 and GIANT across similarity groups. Variants were restricted to those present in both cohorts with allele frequency > 0.001. Bolded values in the top and bottom rows denote the total number of significant associations reported by each dataset, including those not observed in the other cohort. 35
Supplementary Figure 17 | Concordance of height effect sizes between All of Us v8 and GIANT across genetic similarity groups. Comparison of variant-level height effect sizes for A, EUR-like, and B, AFR-like. EAS-like and SAS-like groups are not shown due to limited overlap. Each point represents a variant, with All of Us v8 effect sizes on the x-axis and GIANT effect sizes on the y-axis. Effect sizes were oriented to the positive axis by aligning directions. Error bars denote 95% confidence intervals, with arrows indicating values extending beyond the plotting range. Dashed lines indicate concordance under y = x and y = −x. Point colors denote variant significance categories, defined by combining summary statistics from GIANT, All of Us v8, and pan-UK Biobank. 36
Consistency of burden effect sizes between All by All and Genebass 36
Supplementary Figure 18 | Concordance of rare variant burden associations between All by All v8 and Genebass. A, Comparison of burden effect sizes between Genebass and EUR-like in All by All at maxMAF = 0.001, with effect sizes oriented to the positive axis. Error bars denote 95% confidence intervals, with arrows indicating values extending beyond the plotting range. B, Comparison of burden test p values between Genebass and All by All meta-analysis at maxMAF = 0.001. In both panels, each point represents a gene-level variant group, where color indicates functional annotation, and transparency indicates significance status across the two biobanks. 37
Expected p value ranking and lambda GC 37
Result QC metrics 38
Supplementary Table 8 | Phenotypes excluded during result QC. Phenotypes were excluded if the genomic control inflation factors (λGC) exceeded 2 in any analysis or if meta-analysis ACAF λGC was < 0.8 within variant allele frequency bins with AF > 0.1. 40
Supplementary Figure 19 | Overview of QC pipeline applied to gene-level and single-variant association results. Genomic control inflation factors (λGC) were first computed per phenotype. Phenotypes were excluded if λGC > 2 in any similarity group or if meta-analysis ACAF λGC < 0.8 within the AF > 10% bin. For

gene-level analyses, gene-specific λGC values were summarized across well-calibrated phenotypes and evaluated against sequencing coverage, number of contributing variants, and cumulative allele frequency (CAF), motivating minimum thresholds for coverage, variant count, and CAF. Gene–phenotype associations were further filtered using an expected allele count criterion (CAF × number of cases ≥ 5). For single-variant analyses, λGC was summarized across allele frequency bins, rare variants with unstable λGC estimates were excluded, and variant–phenotype associations were filtered using an expected allele count threshold (AF × number of cases ≥ 5). 41

Supplementary Figure 20 | λGC before and after QC. λGC (y-axis) for gene-level burden association results before and after QC filtering (x-axis), stratified by similarity group (panels) and phenotype category (colored box plots). The horizontal dashed line denotes the null expectation (λGC = 1). 42

Supplementary Figure 21 | λGC before and after AC. λGC (y-axis) for ACAF single-variant association results before and after QC filtering (x-axis), stratified by similarity group (panels) and phenotype category (colored box plots). The horizontal dashed line denotes the null expectation (λGC = 1). 43

Supplementary Figure 22 | λGC before and after QC. λGC (y-axis) for Exome single-variant association results before and after QC filtering (x-axis), stratified by similarity group (panels) and phenotype category (colored box plots). The horizontal dashed line denotes the null expectation (λGC = 1). 44

Analysis of summary statistics 45

Summary of significant associations 45

Supplementary Figure 23 | Cumulative distribution of significant associations across phenotypes by similarity group. The y-axis shows the cumulative phenotype percentile of significant association counts (x-axis) across phenotypes within each genetic similarity group (color). A, Single-variant associations combining both ACAF and Exome results, after QC and LD pruning using PLINK clumping (r2 < 0.1). B, Gene-level burden associations, after QC with maxMAF = 0.001. 46

Supplementary Table 9 | Summary of significant associations after result QC. Shown are numbers of QCed significant association across similarity groups, including the meta-analysis. Results are reported for LD-pruned (r2 < 0.1) single-variant associations (ACAF and Exome), and for gene-level burden tests, stratified by the four functional annotation categories. The bottom row (“Total”) reports the naive sum of association counts across similarity groups after result QC, whereas the penultimate row (“Unique”) reports the number of unique associations after accounting for overlap across similarity groups and restricting to phenotypes with squared correlation r2 < 0.5. 46

Contribution of similarity groups to associations identified from meta-analysis 47

Supplementary Figure 24 | Similarity contributions to meta-analysis gene-level burden associations. Proportion of the unweighted Z-score contributing to meta-analysis associations, defined as 2CAFi(1-CAFi) (-Φ-1 (pi))×sign(βi), where CAFi, pi and βi are the combined allele frequency (CAF), p value and beta effect size from the pLoF burden test for similarity group i (color). Each bar represents a significant association from the meta-analysis (pgene-burden < 6.7 × 10-7), for which none of the component similarity groups presented significance. Colored segments indicate the proportional contribution of each similarity group to the total unweighted Z score chunks. 48

Supplementary Figure 25 | Contribution of similarity groups to association signals relative to sample size. Relationship between the average unweighted Z-score contribution of each similarity group (x-axis), defined as the proportion of the total unweighted Z-score, and the corresponding average sample size across phenotypes (y-axis). Each point represents a similarity group. The dashed line denotes the identity line (y = x). 49
Novelty assessment pipeline for gene-phenotype associations 50
Overview 50
Supplementary Figure 26 | Schematic representation of the novelty assessment agentic pipeline. 51
Literature Analysis 51
Synonym Generation 51
PubMed Search Strategy 51
Novelty Classification 53
Open Targets Analysis 53
Association Score Retrieval and Filtering 53
Phenotype Matching 53
Score-Based Classification 53
Verdict Integration 54
Supplementary Figure 27 | Confusion matrix between the Literature agent scores and Open Targets agent scores across 272 associations significant in only cross-biobank meta-analysis, not in either biobank alone. 54
Supplementary Figure 28 | Rare pLoF burden associations with laboratory measurements identified only through cross-biobank meta-analysis. 89 associations (y-axis) are significant in the meta-analysis of UK Biobank and All of Us, but not in any individual genetic similarity groups or biobanks (x-axis), with 12 classified as potentially novel (red points). Red sequentially colored dots adjacent to each association indicate AI-assisted evidence grading, reflecting the level of proper support in scientific literatures. Blue sequentially colored dots with sign labels represent cohort-specific burden test p value magnitude (color intensity), and effect directions (sign). 56
Supplementary Figure 29 | Rare pLoF burden associations with physical measurements identified only through cross-biobank meta-analysis. 70 associations (y-axis) are significant in the meta-analysis of UK Biobank and All of Us, but not in any individual genetic similarity groups or biobanks (x-axis), with 5 classified as potentially novel (red points). Red sequentially colored dots adjacent to each association indicate AI-assisted evidence grading, reflecting the level of proper support in scientific literatures. Blue sequentially colored dots with sign labels represent cohort-specific burden test p value magnitude (color intensity), and effect directions (sign). 56
References 57

#### *All of Us data description*

##### Genotype Data

The pipeline for generating whole genome sequence (WGS) data from participant biospecimens has been previously described for the Curated Data Repository (CDR) v7 release[1]. The generation of the CDR v8 genomic data used similar methods, with the full Genomic Quality Report providing a detailed description of the curation of the CDR v8 dataset (https://support.researchallofus.org/hc/en-us/articles/29390274413716-All-of-Us-Genomic-Quality-Report). Briefly, the biospecimens undergo a harmonized process of DNA extraction and sequencing with rigorous quality control (QC) processes to generate clinical-grade genomic sequences. In addition to the full callset, short read WGS data is available in smaller callsets (https://support.researchallofus.org/hc/en-us/articles/14929793660948-Smaller-Callsets-for-Analyzing-Short-Read-WGS-SNP-Indel-Data-with-Hail-MT-VCF-and-PLINK), which cover genomics regions that are of particular interest to investigators.

Genetic similarity to reference groups is calculated for each sample based on labels used in gnomAD[2,3], the Human Genome Diversity Project[4], and 1000 Genomes[5]. Individuals are assigned into a genetic similarity group, which have labels as derived from the reference panel, including AFR, AMR, EAS, EUR, MID, and SAS. Throughout, we refer to individuals in one of these groups based on their similarity (e.g. AFR-like), as recommended[6]. Sample relatedness was calculated using the Hail pc_relate function (https://hail.is/docs/0.2/methods/relatedness.html#hail.methods.pc_relate), and any pairs with a kinship score above 0.1 were reported. More information about the samples, including genetic similarity calculations and sample relatedness, is available in the Genomic Quality Report.

#### Phenotype Data

Participant phenotype data is gathered by healthcare organizations involved in the *All of Us* Research Program, who are responsible for participant recruitment as well as data collection and transfer. Data is collected locally from multiple data sources, including surveys, in personal clinical visits, and the electronic health record (EHR), and undergoes a process of data transformation and standardization to the Observational Medical Outcomes Partnership (OMOP) Common Data Model prior to being made available to researchers. More information about the data collection and transformation for survey data, physical measurements, and EHR data can be found in the relevant User Support articles:

- Survey data: https://support.researchallofus.org/hc/en-us/articles/6085114880148-Introduction-to-All-of-Us-Survey-Collection-and-Data-Transformation-Methods
- Physical measurements: https://support.researchallofus.org/hc/en-us/articles/29888188023060-Introduction-to-All-of-Us-Physical-Measurement-Data-Collection-and-Transformation-Methods
- EHR data: https://support.researchallofus.org/hc/en-us/articles/30125602539284-Introduction-to-All-of-Us-Electronic-Health-Record-EHR-Collection-and-Data-Transformation-Methods.

Using health data and biological measurements from the *All of Us* (AoU) Research Program, we curated 10,031 phenotypes of seven different categories, including 10 physical measurements (7 original, 3 derived from the original ones), 86 lab measurements, 1,843 Phecodes (https://phewascatalog.org/phecodes), 3,426 PhecodeX (https://github.com/PheWAS/PhecodeXVocabulary), 145 from self-reported responses, including those from the Mental Health Well-being survey, and the Personal Medical History or Personal and Family Health History surveys, and 4,521 drug/medications (https://athena.ohdsi.org/search-terms/start).

The pipeline for curating phenotypes is described in several Featured Workspaces on the Researcher Workbench. Row-level phenotype data were curated for downstream use in the *All by All* analysis pipeline. Each phenotype is available as an individual notebook within the corresponding Featured Workspace. Each Featured Workspace also includes the row-level phenotype data used in *All by All* analysis and a ReadMe file including an overview of the pipeline, code used for phenotype generation, and an index of all phenotypes.

For categorical phenotypes (Phecode, PhecodeX, Prescriptions, and self-reported responses from Personal and Family Health History and Mental Health and Well Being surveys), a table of True/False values including all phenotypes in the category for each participant was generated for downstream analysis. A minimum case count of two was used for Phecode and PhecodeX phenotypes. After review, only PhecodeX data was included in downstream analysis due to large overlap of phenotypes and more robust coverage of phenotypes using the updated PhecodeX vocabulary[7].

For continuous data types (Physical Measurements, Lab Measurements), participant-level summaries for each phenotype were generated for downstream analysis. For phenotypes in the physical measurement category, mean values for each participant were compiled into a table. For lab measurements, a novel pipeline for harmonizing lab measurements data was developed and used to generate statistics (mean, median, latest, minimum, maximum, and total number of counts) for each lab measurement phenotype for each participant[8].

Overall, prevalence estimates were broadly consistent for disease phenotypes shared across UK Biobank[9] and the *All of Us* Research Program and across genetic similarity groups (**Supplementary Figure 1**; **Supplementary Table 1**). Observed discrepancies are likely attributable to differences in phenotyping methods and sampling frameworks. In particular, the

UK Biobank comprises predominantly healthy volunteers over the age of 40 with relatively high socioeconomic status, whereas *All of Us* includes a more heterogeneous cohort recruited from both clinical settings and community populations across the United States.

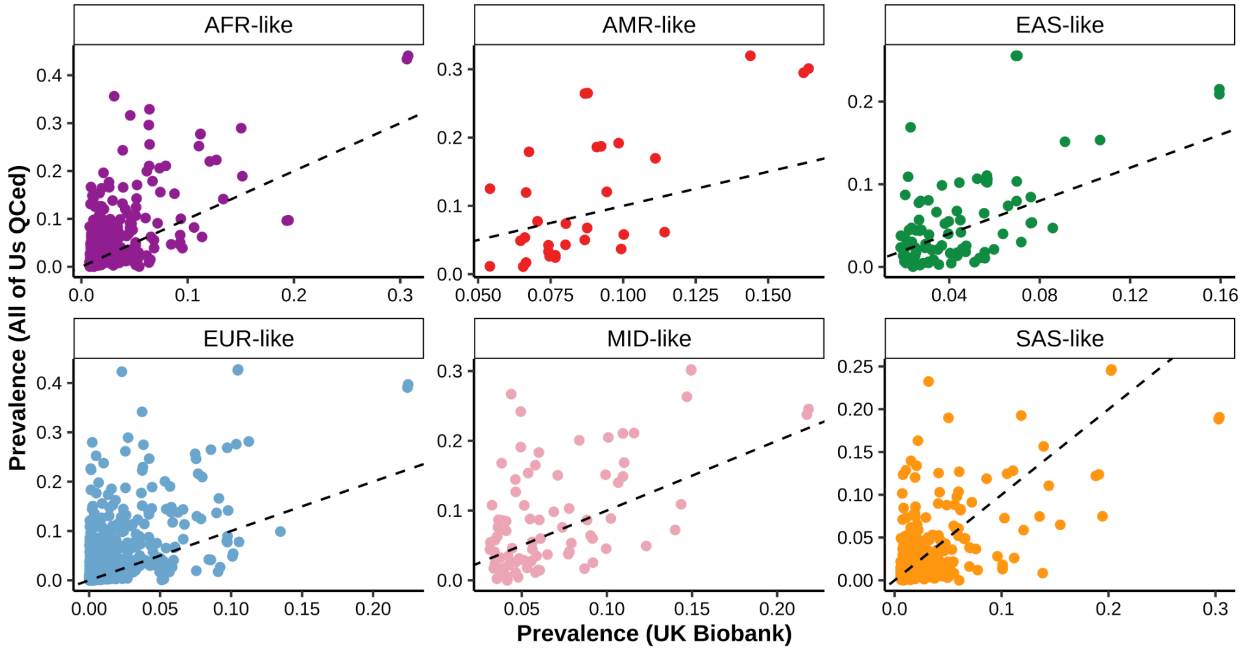


**Supplementary Figure 1** | Cross-biobank consistency of phenotype prevalence. Comparison of Phecode prevalence between UK Biobank (x-axis) and *All of Us* v8 (y-axis) across genetic similarity groups (panels and colors). Each point represents a Phecode. The dashed lines represent y=x.

| | | **Case count** | | **Control count** | | **Prevalence** | |
|---|---|---|---|---|---|---|---|
| **Similarity Group** | **N Phenos Shared** | **r** | ***p*** | **r** | ***p*** | **r** | ***p*** |
| AFR-like | 335 | 0.660 | $2.98 \times 10^{-43}$ | 0.829 | $5.99 \times 10^{-86}$ | 0.623 | $2.49 \times 10^{-37}$ |
| AMR-like | 31 | 0.707 | $8.86 \times 10^{-6}$ | 0.657 | $5.86 \times 10^{-5}$ | 0.687 | $1.97 \times 10^{-5}$ |
| EAS-like | 87 | 0.653 | $7.02 \times 10^{-12}$ | 0.896 | $1.23 \times 10^{-31}$ | 0.624 | $1.08 \times 10^{-10}$ |
| EUR-like | 1,326 | 0.622 | $1.19 \times 10^{-142}$ | 0.868 | 0 | 0.608 | $6.97 \times 10^{-135}$ |
| MID-like | 81 | 0.579 | $1.53 \times 10^{-8}$ | 0.715 | $6.60 \times 10^{-14}$ | 0.570 | $2.73 \times 10^{-8}$ |
| SAS-like | 410 | 0.624 | $1.45 \times 10^{-45}$ | 0.906 | $1.22 \times 10^{-154}$ | 0.609 | $5.71 \times 10^{-43}$ |
| Overall | 2,270 | 0.621 | $1.15 \times 10^{-242}$ | 0.968 | 0 | 0.614 | $5.85 \times 10^{-235}$ |

**Supplementary Table 1** | Cross-biobank concordance of Phecode sample size and prevalence. Pearson's correlation coefficient (r) and corresponding *p* values for the number of cases, the number of controls, and prevalence for Phecodes in UK Biobank and *All of Us* v8 across similarity groups.

#### *Pre-GWAS QC*

##### Genotype QC

We performed genotype-level QC on both the Allele Count/Allele Frequency (ACAF) threshold callset and the Exome callset obtained from the v8 release in *All of Us* Researcher Workbench(https://support.researchallofus.org/hc/en-us/articles/29475228181908-How-the-All-of-Us-Genomic-data-are-organized). Variants were restricted to those that passed Variant Quality Score Recalibration (VQSR), as indicated by the 'FT' field in the callset, and exhibited a global allele count (AC) > 0. Genotype filtering was based on a modified version of the previously defined "adj" criteria in gnomAD[2,3]. Specifically, we retained genotypes with genotype quality (GQ) ≥ 30, and a minor allele balance > 0.2 across all alternate alleles for heterozygous calls. We also removed genotypes on chromosome Y for samples with self-reported sex as female.

##### Sample QC

At the sample level, we applied a series of hard QC filters. First, we excluded individuals with reported age > 100 years, those with sex (self-reported sex at birth) not annotated as female or male, those flagged by the *All of Us* QC report, and those without a similarity group assignment based on principal component analysis (PCA) derived from the AoU dataset. PCA was conducted using Hail's hwe_normalized_pca method (https://hail.is/docs/0.2/methods/genetics.html#hail.methods.hwe_normalized_pca). We also removed 1,971 duplicate samples identified using kinship coefficients from the AoU relatedness table, which was computed using Hail's pc_relate method. Pairs of samples with a kinship coefficient > 0.375 were considered duplicates, and we retained only one sample from each related pair to produce a maximal set of unique individuals (**Supplementary Figure 2**).

Pruning similarity group outliers and group-specific PCA

To further reduce genetic similarity group stratification, we removed outlier samples from each pre-assigned genetic similarity group. Specifically, we reran PCA within each similarity group using the subset of samples that passed the initial hard filters (**Supplementary Figure 3**). Group outliers were then identified and removed based on their multidimensional distances from the group-specific centroid in PC space.

Starting from the ACAF callset, we applied the sample QC (except for the final PC pruning metric determined here) and genotype QC steps as described above to the ACAF callset. For each group, we first downsampled to one million autosomal SNPs from the ACAF callset, randomly selecting variants with allele frequency (AF) ≥ 0.01 and group-specific call rate ≥ 0.9, and excluding those located within the HLA region or the chromosome 8 inversion. We then apply linkage disequilibrium (LD) pruning on these variants at $r^2$ = 0.1 (**Supplementary Table 2**).

Using this LD-pruned variant set, we recomputed PCA for individuals within each assigned genetic similarity group, restricting the PCA training set to samples identified as unrelated in the *All of Us* relatedness table, and subsequently projecting related individuals into the unrelated PC space. We quantified each individual's distance to similarity group centroids across the first 10 PCs. We computed distances relative to 3-5 centroids per group, and removed samples exceeding the group-specific outlier threshold, depending on the degree of within-group heterogeneity defined as below:

$$d = \sum_{i=1}^{n} \frac{(X_i - \overline{X}_i)^2}{\sigma_{X,i}^2}$$

where $d$ is the total centroid distance summed over $n$ total dimensions of an ellipse (i.e., PCs), $X_i$ is a vector of PCs, $\overline{X}_i$ is the mean PC score, and $\sigma_{X,i}^2$ is the variance of the PC scores for the $i^{th}$ PC.

We identified similarity group outliers by examining the distribution of centroid distances within each group. Specifically, we plotted histograms of these distances and excluded individuals falling in the extreme upper tail[10] (**Supplementary Figure 4-9**). After outlier removal, approximately 96% of samples (392,030/407,490) were retained. We then repeated PCA using only the unrelated individuals from the refined sample set and subsequently projected the related individuals into the resulting PC space. This procedure yielded a complete set of group-specific PCA coordinates for all pruned samples, later to be used in association testing (**Supplementary Table 2**).

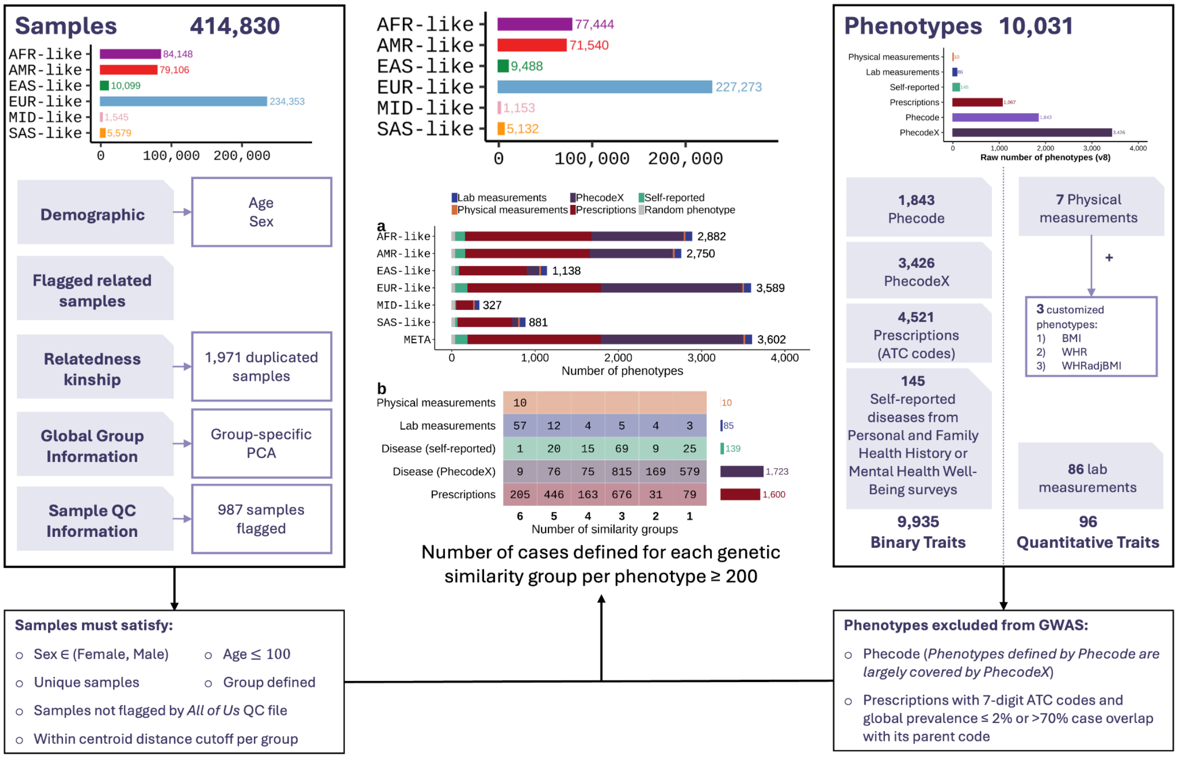


**Supplementary Figure 2** | Sample QC and phenotype QC pipeline. The left column describes sample information obtained from the *All of Us* Researcher Workbench and filtering metrics used for sample QC. The right column describes phenotype categories and the selection criteria for phenotypes included in association testing. The middle column bar charts show the number of samples and phenotypes across similarity groups and phenotypic categories after applying sample and phenotype QC filters and then restricting to phenotypes with a group-specific number of cases ≥ 200. The bottom matrix shows the number of phenotypes (cells) within each phenotype category (y-axis) stratified by the number of genetic similarity groups sharing each phenotype (x-axis). Bars to the right of the matrix indicate total phenotypes analyzed per category.

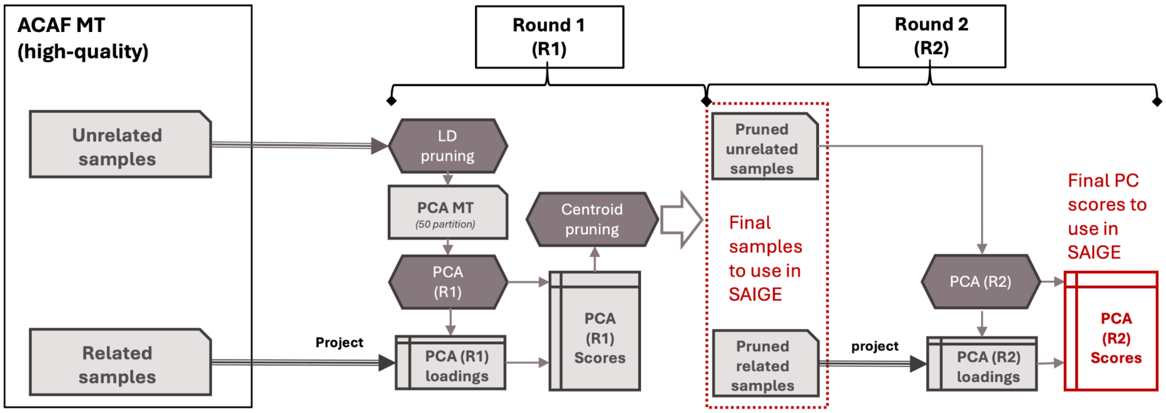


**Supplementary Figure 3** | PC-based centroid outlier pruning pipeline. Starting from the genotype and sample-QCed ACAF callset, samples were separated by relatedness. PCA was performed on unrelated samples and then computed for related samples by projecting the related genotypes on the unrelated PC space. Using these PC scores, centroid outliers were removed, and PCA was recomputed on the pruned set using the same procedure as described earlier for the first round.

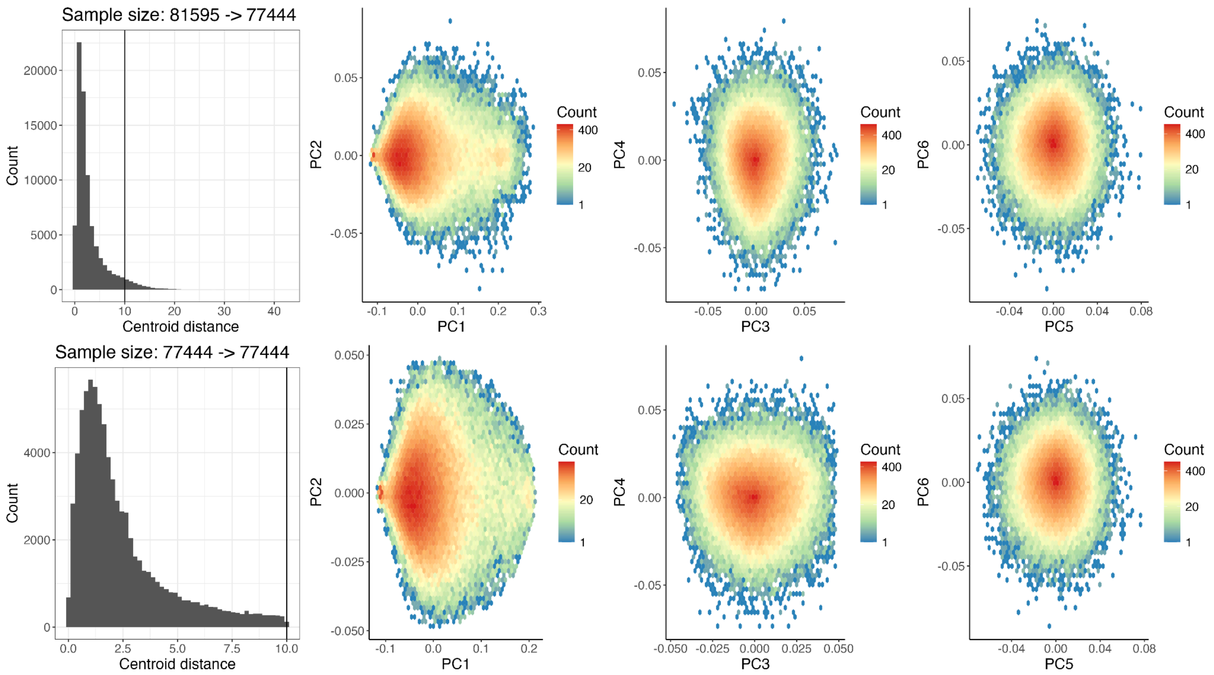


**Supplementary Figure 4** | PCA in *All of Us* v8 participants assigned to AFR-like and corresponding centroid distance across 3 PCs. Centroid distance distributions and PC biplots for the first 6 PCs are shown before (top) and after (bottom) pruning outliers. The vertical line in the two left centroid distance histograms shows the threshold chosen to remove outliers.

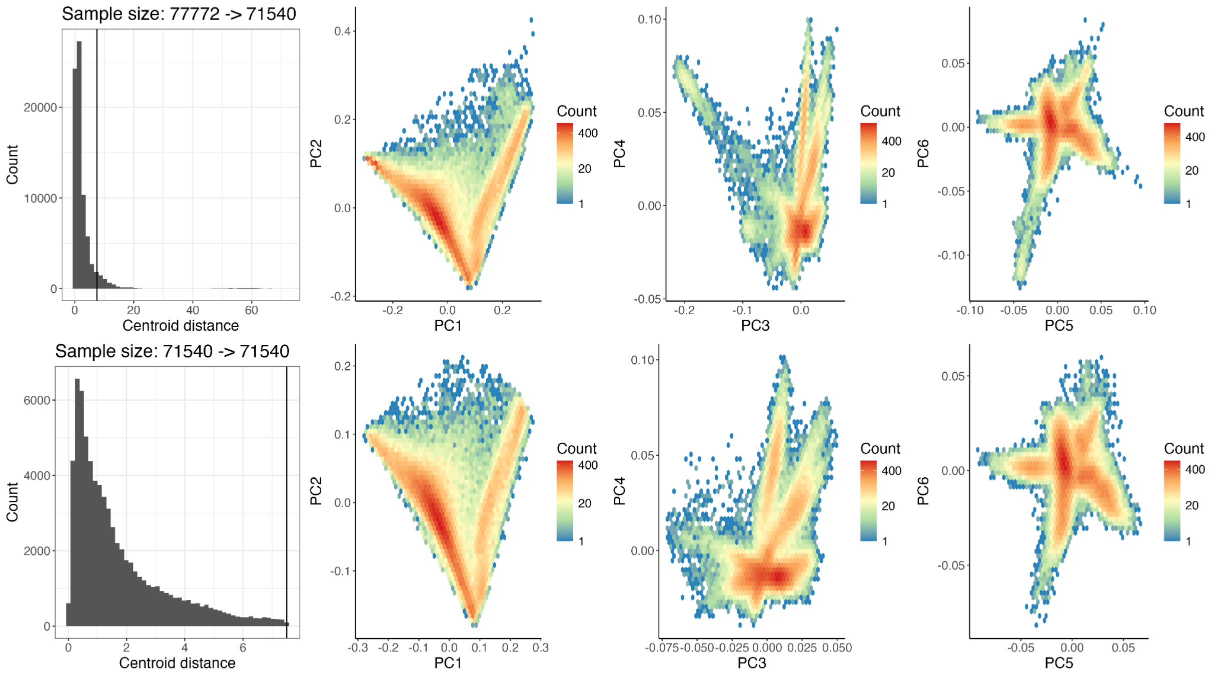


**Supplementary Figure 5** | PCA in All of Us v8 participants assigned to AMR-like and corresponding centroid distance across 3 PCs. Centroid distance distributions and PC biplots for the first 6 PCs are shown before (top) and after (bottom) pruning outliers. The vertical line in the two left centroid distance histograms shows the threshold chosen to remove outliers.

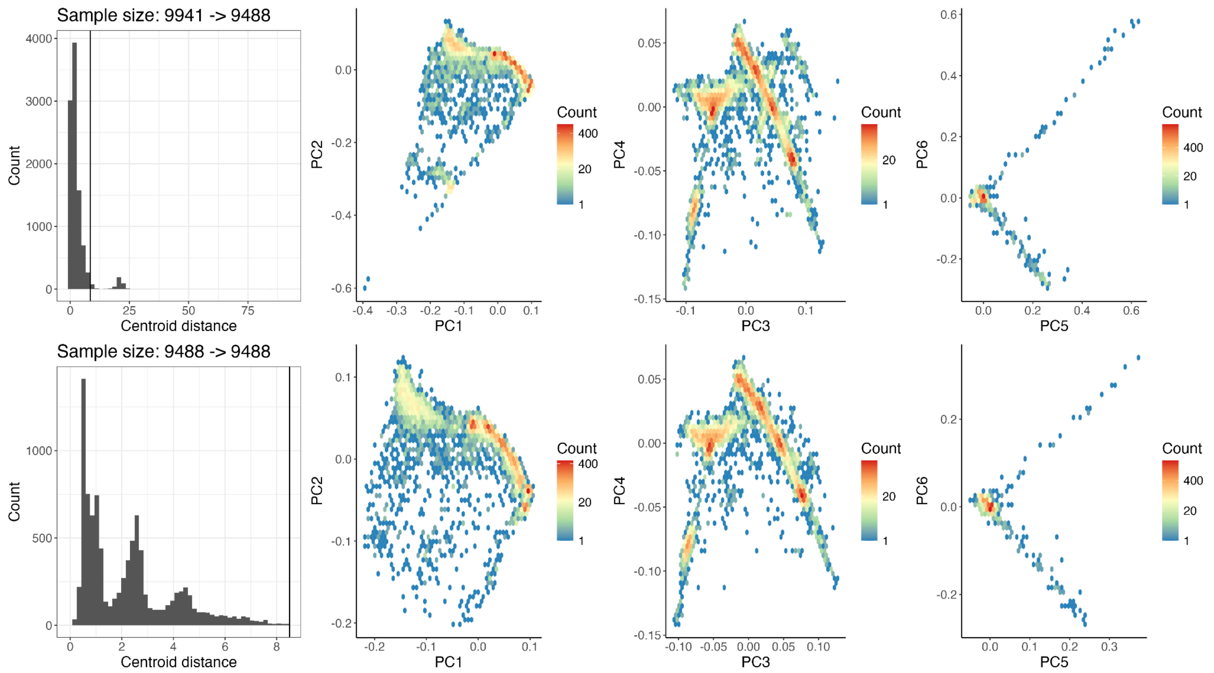


**Supplementary Figure 6** | PCA in *All of Us* v8 participants assigned to EAS-like and corresponding centroid distance across 3 PCs. Centroid distance distributions and PC biplots for the first 6 PCs are shown before (top) and after (bottom) pruning outliers. The vertical line in the two left centroid distance histograms shows the threshold chosen to remove outliers.

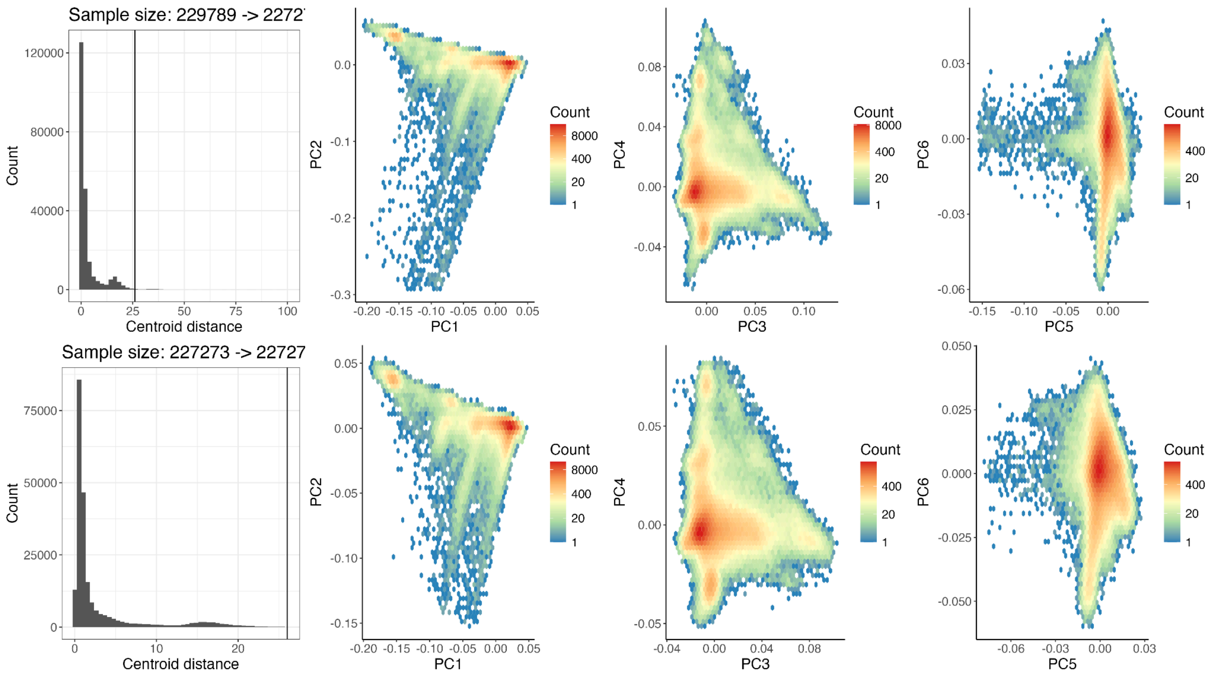


**Supplementary Figure 7** | PCA in *All of Us* v8 participants assigned to EUR-like and corresponding centroid distance across 5 PCs. Centroid distance distributions and PC biplots for the first 6 PCs are shown before (top) and after (bottom) pruning outliers. The vertical line in the two left centroid distance histograms shows the threshold chosen to remove outliers.

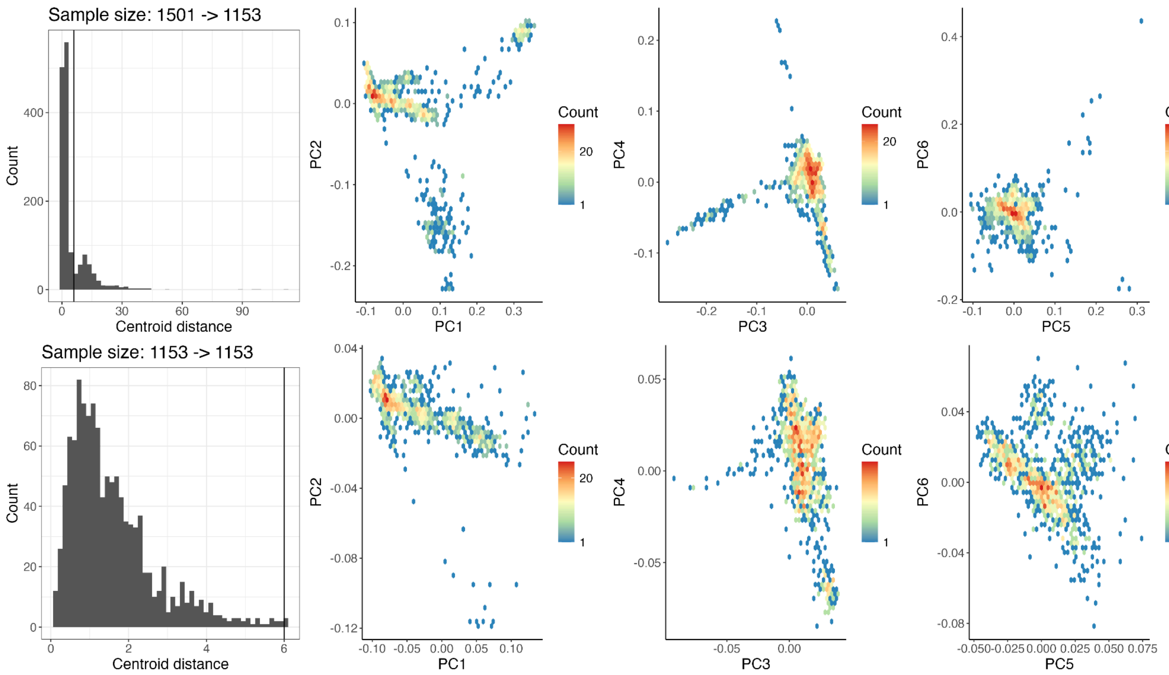


**Supplementary Figure 8** | PCA in *All of Us* v8 participants assigned to MID-like and corresponding centroid distance across 5 PCs. Centroid distance distributions and PC biplots for the first 6 PCs are shown before (top) and after (bottom) pruning outliers. The vertical line in the two left centroid distance histograms shows the threshold chosen to remove outliers.

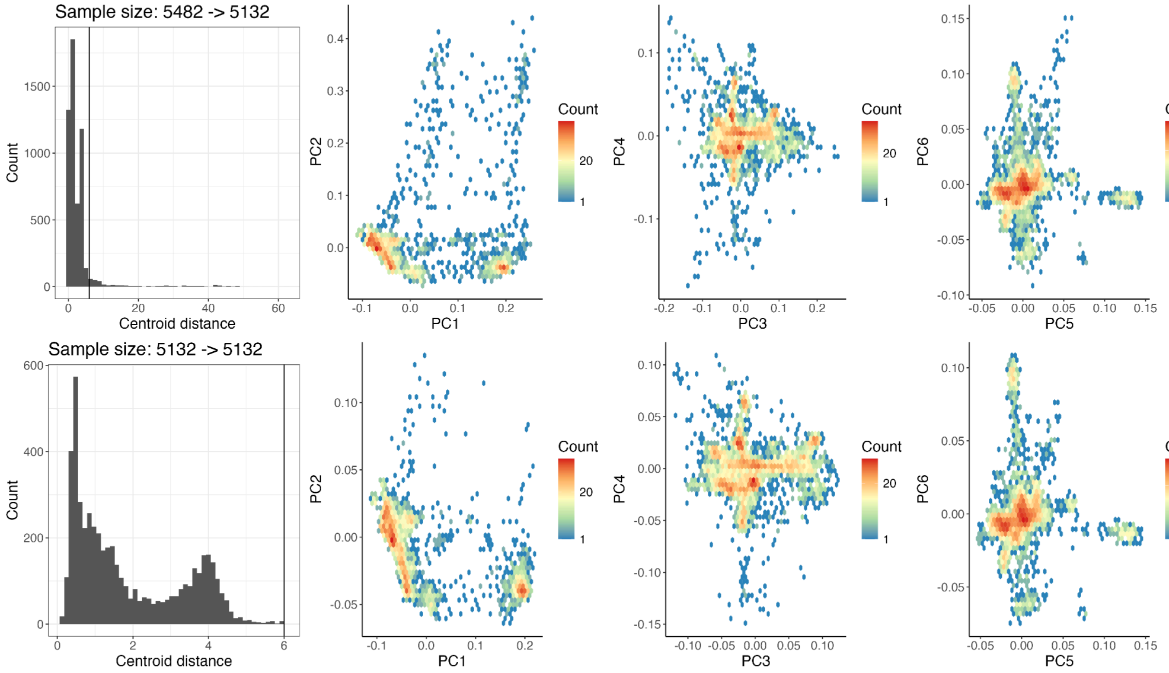


**Supplementary Figure 9** | PCA in *All of Us* v8 participants assigned to SAS-like and corresponding centroid distance across 3 PCs. Centroid distance distributions and PC biplots for the first 6 PCs are shown before (top) and after (bottom) pruning outliers. The vertical line in the two left centroid distance histograms shows the threshold chosen to remove outliers.

| | Raw sample size | N samples (post initial QC) | | | N samples (pruned) | | | N variants (pre-down sampling) | N PCA variants (post-LD-pruning) |
|---|---|---|---|---|---|---|---|---|---|
| | | Unrelated | Related | Total | Unrelated | Related | Total | | |
| **AFR-like** | 84,148 | 72,628 | 8,967 | 81,595 | 68,810 | 8,634 | 77,444 (94.91%) | 14,500,300 | 302,378 |
| **AMR-like** | 79,106 | 69,587 | 8,185 | 77,772 | 64,114 | 7,426 | 71,540 (91.99%) | 9,522,180 | 210,078 |
| **EAS-like** | 10,099 | 9,576 | 365 | 9,941 | 9,150 | 338 | 9,488 (95.44%) | 7,818,375 | 190,779 |
| **EUR-like** | 234,353 | 219,969 | 11,230 | 231,199 | 216,236 | 11,037 | 227,273 (98.30%) | 8,591,656 | 193,886 |
| **MID-like** | 1,545 | 1,428 | 73 | 1,501 | 1,090 | 63 | 1,153 (76.82%) | 9,639,664 | 225,426 |
| **SAS-like** | 5,579 | 5,281 | 201 | 5,482 | 4,944 | 188 | 5,132 (93.62%) | 8,920,134 | 211,161 |

**Supplementary Table 2** | Sample and variant kept across QC stages by genetic similarity groups. Number of samples shown after hard filtering (post initial QC) and centroid pruning (pruned), along with variant counts before downsampling and after LD pruning.

##### Phenotype curation

We removed prescription phenotypes corresponding to 7-digit subcategory ATC codes when their global prevalence was ≤ 2% or when >70% of their cases overlapped with those of their parent ATC code, thereby reducing redundancy and minimizing potential misclassification of rare drug codes. To ensure adequate statistical power for downstream association analyses, we further restricted the phenotype set to those with at least 200 cases passing QC within each similarity group, yielding 11,556 group-phenotype pairs (**Supplementary Figure 2** and **Supplementary Table 3**). Following this filtering, we designated phenotypes with > 90% of cases originating from a single sex (self-reported sex at birth as female or male) as sex-specific and retained only individuals of the majority sex group. As a result, some phenotypes may appear with slightly fewer than 200 cases after sex-specific filtering

| | Lab measurement | Physical measurement | Random phenotype | PhecodeX | Prescription | Self-reported | Total |
|---|---|---|---|---|---|---|---|
| AFR-like | 81 | 10 | 45 | 1,120 | 1,518 | 108 | 2,882 |
| AMR-like | 79 | 10 | 45 | 1,011 | 1,493 | 112 | 2,750 |
| EAS-like | 73 | 10 | 45 | 160 | 814 | 36 | 1,138 - 3 |
| EUR-like | 85 | 10 | 45 | 1,711 | 1,600 | 138 | 3,589 |
| MID-like | 57 | 10 | 45 | 9 | 205 | 1 | 327 - 5 |
| SAS-like | 69 | 10 | 45 | 85 | 651 | 21 | 881 - 3 |
| **Total** | **444** | **60** | **270** | **4,096** | **6,281** | **416** | **11,556** |
| META (unique) | 85 | 10 | 45 | 1,723 | 1,600 | 139 | 3,602 |

**Supplementary Table 3** | Phenotypes kept after sample QC and phenotype selection by phenotype category and genetic similarity group. Values subtracted from the total column on the right represent rare random phenotypes lacking defined cases in the three smallest similarity groups and therefore excluded due to null model non-convergence in SAIGE step1. (Note: random phenotypes span prespecified prevalence ranges for result validation, and are not subject to the case count filters applied to other phenotypes).

#### *Association testing*

For each genetic similarity group, we used the genotype data from the Exome call set that passed the genotype- and sample-level QC procedures described above. We constructed a group-stratified subset of autosomal variants with group-specific minor allele count (MAC) > 0 and call rate ≥ 90%. From this set, we sampled approximately 2,000 variants from each allele count (AC) bin of 1–5, 6–10, and 11–20; approximately 10,000 variants from AC 20 to allele frequency (AF) 0.1% (not available for EAS-like, MID-like, or SAS-like) and from AF 0.1–1% (corresponding to AC 2–23 in MID-like and overlapping with earlier bins); and approximately 100,000 variants with AF > 1%. After linkage disequilibrium pruning at $r^2 < 0.1$, variants were exported to PLINK files. Sparse genetic relationship matrices (GRM) were then generated for each genetic similarity group using SAIGE step 0 with default parameters, including 2,000 markers for kinship estimation and a relatedness cutoff of 0.125, using variants with AF > 1% from the subset PLINK files (**Supplementary Figure 14**). The group-specific PLINK files and GRMs were both used as input files in step 1 of SAIGE and SAIGE-GENE+ for constructing the null models (**Extended Data Figure 1**).

For each phenotype-group pair, we generated a covariate file including phenotype values and model covariates: age, sex, age$^2$, 20 group-specific PCs recomputed from centroid-pruned samples, and interaction terms (age × sex, and age$^2$ × sex). Quantitative phenotypes were inverse-rank normal transformed prior to analysis. Genotypes were extracted, subset, and converted to BGEN format with Hail (https://hail.is/). Using the post-QC phenotype and genotype datasets, we performed 11,556 group-specific single-variant tests for common variants and exonic variants using SAIGE[11], and 11,556 series of gene-level rare variant burden tests using SAIGE-GENE+[12,13], which simultaneously conduct burden test, SKAT[14], and SKAT-O[14,15] for aggregated predicted loss-of-function (pLoF), missense, synonymous, pLoF + missense variants and combinations thereof within each gene at a maximum minor allele frequency (MAF) of 1%, 0.1%, and 0.01%. Thus, for each phenotype and similarity group combination, we produced three sets of results: (1) single-variant association testing results of common variants from the ACAF callset, (2) single-variant association testing results of exonic variants from the exome callset, and (3) gene-level RVAS results from the exome callset, for 3,602 unique phenotypes across all similarity groups available.

#### SAIGE

We conducted genome-wide association studies (GWAS) using the QCed *All of Us* ACAF genotype dataset and the SAIGE software (v1.4.8). In Step 1, we fitted phenotype-specific null models (excluding genotypes) using the curated phenotype data, a sparse genetic relationship matrix (GRM), PLINK-formatted genotype files, default SAIGE parameters, and the covariates described above. For Step 2, genotypes from the ACAF callset were partitioned into BGEN interval files stratified by genetic similarity group: 136 intervals of 25 Mb for EAS-like, MID-like, and SAS-like groups; 259 intervals of 12.5 Mb for AFR-like and AMR-like; and 998 intervals of 3.125 Mb for EUR-like (**Extended Data Figure 1**). Association testing was then performed using these group-specific BGEN files together with the phenotype-specific null models generated in step 1, with default SAIGE parameters and leave-one-chromosome-out (LOCO) disabled (LOCO = FALSE).

For the *All of Us* Exome callset, we similarly executed Step 1 of SAIGE to fit null models using the same data inputs as described above, but additionally specified the arguments cateVarRatioMinMACVecExclude = 0.5, 1.5, 2.5, 3.5, 4.5, 5.5, 10.5, 15.5, 20.5 and cateVarRatioMaxMACVecInclude = 1.5, 2.5, 3.5, 4.5, 5.5, 10.5, 15.5, 20.5 to enable computation of variance ratio estimates for ultra-rare variants (allele count < 10). Exome genotypes were partitioned into BGEN interval files by similarity group, with 99 intervals containing approximately 200 nearby genes each for EAS-like, MID-like, and SAS-like, and 198 intervals containing approximately 100 nearby genes each for AFR-like, AMR-like, and EUR-like. In Step 2, association testing was performed using these BGEN files, together with the phenotype-specific null models, using default SAIGE parameters, LOCO = FALSE, and is_single_in_groupTest = TRUE. The same cateVarRatioMinMACVecExclude and cateVarRatioMaxMACVecInclude settings described above were also applied during Step 2 to ensure appropriate modeling of ultra-rare variants.

##### SAIGE-GENE+

We performed gene-level rare variant association studies (RVAS) using the QCed genotype data from the *All of Us* exome callset and SAIGE-GENE+ v1.4.8. To enable gene-level tests of rare variants sharing the same functional annotation, we generated “gene map” files corresponding to each genomic interval of ~100 or ~200 adjacent protein-coding genes, consistent with the BGEN partitions used for the SAIGE exome analyses described in the previous section (**Extended Data Figure 1**). Variant annotation followed the scheme used by Genebass.org[16], using VEP v105 as implemented in Hail with default parameters for GRCh38, including LOFTEE[2]. We defined four functional variant groups: (1) predicted loss-of-function (pLoF) variants, limited to high-confidence LOFTEE annotations; (2) missense-like variants, including missense variants and variants annotated by LOFTEE as low-confidence loss-of-function; (3) synonymous variants; and (4) a combined group containing both pLoF and missense-like variants.

We then performed Step 2 of SAIGE-GENE+, conducting gene-level burden tests using the same BGEN input files as in the exome GWAS but additionally incorporating the gene-group files that defined variant sets by functional category and genomic interval, plus the same null models generated during Step 1 of SAIGE for the exome analyses. Step 2: Association testing was conducted with default parameters, supplemented with the following settings:

- annotation_in_groupTest = pLoF, missenseLC, synonymous, pLoF:missenseLC
- maxMAF_in_groupTest = 0.01, 0.001, 0.0001
- LOCO = FALSE
- is_single_in_groupTest = TRUE
- cateVarRatioMinMACVecExclude = 0.5, 1.5, 2.5, 3.5, 4.5, 5.5, 10.5, 15.5, 20.5
- cateVarRatioMaxMACVecInclude = 1.5, 2.5, 3.5, 4.5, 5.5, 10.5, 15.5, 20.5

These parameters enable set-based association tests (Burden, SKAT, and SKAT-O) for all QCed phenotypes across six similarity groups, including up to four functional variant groups (pLoF, missense-like, synonymous, and pLoF + missense-like) and three maximum minor allele frequency cutoffs for grouping rare variants (0.01, 0.001, 0.0001). Additionally, as a key feature of SAIGE-GENE+[17], we adopted the default parameter MACCutoff_to_CollapseUltraRare=10, which collapses ultra rare variants with MAC <= 10 into a single pseudo-variant that is tested jointly with other rare variants in the gene-based tests. This approach improves statistical calibration and helps control type 1 error rates in rare variant set-based tests.

##### Meta-analysis

For single-variant association results derived from both the ACAF and Exome call sets, we performed a fixed-effect inverse-variance weighted meta-analysis across all similarity groups with at least 200 cases available for each phenotype. For gene-based association tests, including burden test, SKAT, and SKATO, we integrated group-specific test statistics using Stouffer's method[18], which computes a weighted Z-score for similarity group $i$ and gene-annotation group $g$ as:

$$Z_{weighted,i,g} = \sqrt{2 * CAF_{i,g} * (1 - CAF_{i,g}) * N_i} \times (-\Phi^{-1}(p_i)) \times sign(\beta_i),$$

where, for the weight, $CAF_{i,g}$ denotes the combined allele frequency of variants in the corresponding annotation group within the protein coding gene specified for similarity group $i$, and $N_i$ denotes the effective sample size for similarity group $i$. The variable $p_i$ corresponds to the group-specific *p* value and is defined as $p_i = p/2$ for the two-tailed Burden test and $p_i = p$ for SKAT and SKATO. The sign term $\beta_i$ is taken as the estimated Burden effect size for two-tailed Burden tests and set to 1 for genes where SKAT and SKATO results are defined.

The meta-analytic Z-score across all similarity groups for gene-annotation group $g$ is then calculated as:

$$Z_{meta,g} = \frac{\sum_i Z_{weighted,i,g}}{\sqrt{\sum_i 2*CAF_{i,g}*(1-CAF_{i,g})N_i}}.$$

The *p* value for the two-tailed Burden meta-analysis is subsequently computed as:

$$P_{Burden,\, meta,g} = 2 \times [1 - \Phi(|Z_{meta,g}|)],$$

And the one-tailed SKAT/SKATO meta-analysis *p* value is computed as:

$$P_{one-tail,\, meta,g} = 1 - \Phi(|Z_{meta,g}|)$$

This approach enables harmonized gene-based association inference across multiple similarity groups by appropriately weighting group-specific statistical evidence according to sample size and allele frequencies of variants from the same functional category.

**Meta-analysis across the UK Biobank and *All of Us***

We adopted two complementary approaches to harmonize phenotypes between UK Biobank and *All of Us*. For quantitative phenotypes, including laboratory- and physical-measurement traits, we performed manual curation by fuzzy string matching of phenotype descriptions between two biobanks, followed by detailed review to confirm semantic equivalence. For disease phenotypes, we computed pairwise correlations between ICD-10 codes and Phecodes using phenotype data from Pan UK Biobank[10]. We retained only positively correlated pairs with squared correlation $r^2 \geq 0.8$, and for each Phecode selected the ICD-10 code with the highest $r^2$. An analogous procedure was applied within *All of Us* to map Phecode and PhecodeX. Finally, we linked *All of Us* PhecodeX to ICD-10 codes by merging two maps through the shared Phecode to produce a PhecodeX-Phecode-ICD10 chain. Each PhecodeX was then linked to UKB Genebass phenotype identifier via its mapped ICD10 code. We resolved the conflict by retaining the match with the highest combined score ($r^2_{\text{PhecodeX-Phecode}} \times r^2_{\text{Phecode-ICD10}}$) for cases where multiple PhecodeX phenotypes were mapped to the same UKB phenotype. The resulting disease phenotype map was then combined with the manually curated quantitative trait map yielding a total of 222 mapped phenotypes, including 42 laboratory measurements, 10 physical measurements, and 170 disease phenotypes, spanning 15 disease domains (**Supplementary Table 4**). Entry-level QC filters (expected CAC, etc) are not applied to cross-biobank meta-analysis summary statistics given the large sample size and well-calibrated statistic distributions.

We note that methodological choices in the meta-analysis may influence inference. Single-variant associations were combined using inverse-variance weighted meta-analysis, which is appropriate when effect estimates and standard errors are directly comparable across strata. Gene-level rare variant burden associations were combined using Stouffer’s method because burden and SKAT-based tests summarize variant aggregations and may not yield a single effect size with a consistent direction or interpretation across genes, variant masks, and cohorts. This approach provides a practical framework for combining gene-level signals, but its results may still be influenced by differences in effective sample size and phenotype prevalence across cohorts.

| Laboratory Measurement | PhecodeX | Physical Measurement |
|---|---|---|
| 42 | 170 | 10 |

⇓

| | | | | |
|---|---|---|---|---|
| Blood/Immune | Cardiovascular | Congenital | Dermatological | Endocrine/metabolic |
| 8 | 21 | 6 | 19 | 12 |
| Gastrointestinal | Genetic | Genitourinary | Infections | Mental |
| 19 | 2 | 7 | 7 | 8 |
| Muscloskeletal | Neoplasms | Neurological | Respiratory | Sense organs |
| 4 | 15 | 12 | 16 | 14 |

**Supplementary Table 4** | Cross-biobank phenotype mapping overview. Summary of phenotypes mapped between *All of Us* and UK Biobank, with counts stratified by broad phenotypic classes and disease categories.

##### Meta-analysis vs. Mega-analysis

Besides the meta-analyses results incorporated from multi-group associations, we explored an all-in-one mega-analysis pipeline to increase sample size and maximize power. We compute a PC-adjusted genetic relatedness matrix[19] for 407,490 high-quality samples (pre-PC centroid pruning) and apply the same association framework to the full cohort without similarity group stratification. We evaluate the performance of the mega-analysis pipeline on simulated phenotypes and compare its real phenotype association results to those from the meta-analyses pipeline (**Supplementary Figure 10A**-**B**). Overall, meta-analysis and mega-analysis performed similarly well on the simulated quantitative phenotype. However, for real-world phenotypes with slightly smaller sample sizes, meta-analysis results tend to have a lower genomic inflation control factor ($\lambda_{GC}$), indicating reduced power, better controlled population stratification, or a combination thereof (**Supplementary Figure 10C-F**). Because these were largely similar, we opted for the meta-analysis approach to maintain tight type I error control.

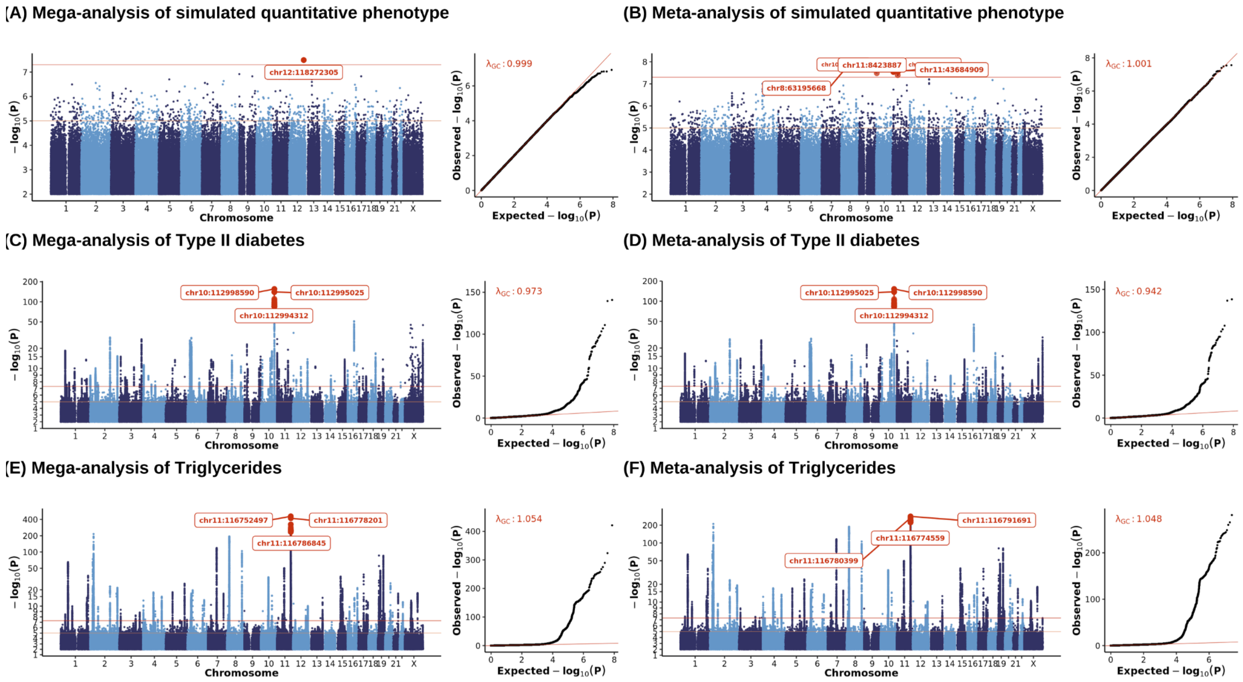


**Supplementary Figure 10** | Meta- v.s. mega-analysis concordance across phenotypes. Manhattan and quantile-quantile (QQ) plot comparison between meta-analysis and mega-analysis across selected phenotypes. Each point represents a common variant from the *All of Us* ACAF callset. In the Manhattan plots, the x-axis shows genomic position, and the y-axis shows $-\log_{10}(p)$. Chromosome and position for variants with $p < 5 \times 10^{-8}$ are highlighted in red. In the QQ-plots, expected versus observed $-\log_{10}(p)$ are shown, with expectation based on the $\chi^2$ distribution. The red line indicates y=x and $\lambda_{GC}$ is displayed in the upper left. **A-B**, Simulated quantitative phenotype with index 1: mega-analysis (N = 407,490) and meta-analysis (N = 392,030). **C-D,** Type II Diabetes (PhecodeX: EM_202.2): mega-analysis (N = 302,693; $N_{cases}$=51,058, $N_{controls}$=251,635) and meta-analysis (N = 290,052; $N_{cases}$= 48,828, $N_{controls}$=241,224). **E-F**, Triglycerides (3022192): mega-analysis (N = 148,030) and meta-analysis (N = 143,386).

#### Computational framework

We implemented this pipeline using Hail Batch (https://hail.is/docs/batch/index.html), which enables efficient, scalable, and highly parallel computation. Hail Batch is a Python-based framework for defining computational tasks and their dependencies as a directed acyclic graph. The pipeline was executed on the Hail Batch Service, a managed multi-tenant compute cluster on Google Cloud, which provides elastic scaling on spot instances and robust resource management. The full association testing pipeline ran over 9 million jobs (**Supplementary Table 5**), which completed in approximately two weeks of wall-clock time, consuming over 2 million CPU hours at a total cost of approximately 70,000 USD across all three analyses (GWAS, ExWAS, and gene-level RVAS). This corresponds to an amortized cost of roughly 10 USD per phenotype when considering all three association types plus additional costs from pre-GWAS data processing (**Supplementary Figure 11**). In contrast, the unamortized cost of analyzing a single phenotype is substantially higher—on the order of hundreds of USD per similarity group—largely due to the additional computational expense of generating genetic relationship matrices (GRMs) and BGEN files for varying sample sizes, which are not included in the cost estimate reported here.

| | AFR-like | AMR-like | EAS-like | EUR-like | MID-like | SAS-like | META | Total |
|---|---|---|---|---|---|---|---|---|
| **N phenotypes** | **2,882** | **2,750** | **1,135** | **3,589** | **322** | **878** | **3,602** | **GWAS: 15,158** |
| N ACAF variants | 66,579,788 | 67,084,982 | 33,200,692 | 71,029,353 | 32,529,713 | 37,447,844 | **87,421,121** | |
| N exome variants | 12,748,718 | 12,696,740 | 4,584,172 | 23,831,780 | 1,609,982 | 3,733,048 | **38,868,098** | **Single-variant Associations: 1,333,410,488,573** |
| **N variants** | 79,328,506 | 79,781,722 | 37,784,864 | 94,861,133 | 34,139,695 | 41,180,892 | **126,289,219** | |
| **N associations** | 228,624,754,292 | 219,399,735,500 | 42,885,820,640 | 340,456,606,337 | 10,992,981,790 | 36,156,823,176 | **454,893,766,838** | |
| N gene groups | 248,352 | 247,906 | 235,604 | 250,956 | 146,093 | 229,743 | **253,116** | **Burden Associations: 3,726,063,720** |
| **N associations** | 715,750,464 | 681,741,500 | 267,410,540 | 900,681,084 | 47,041,946 | 201,714,354 | **911,723,832** | |
| N chunks | 257 | 257 | 138 | 960 | 138 | 138 | **NA** | |
| **N ACAF jobs** | **737,792** | **704,000** | **156,630** | **3,445,440** | **44,436** | **121,164** | **7,204** | **Jobs: 9,356,476** |
| N chunks | 397 | 397 | 199 | 397 | 199 | 199 | **NA** | |
| **N gene jobs** | **1,144,154** | **1,091,750** | **225,865** | **1,424,833** | **64,078** | **174,722** | **14,408** | |

**Supplementary Table 5** | Association testing workload summary. Total number of association tests performed and jobs executed, stratified by result type across genetic similarity groups and meta-analysis.

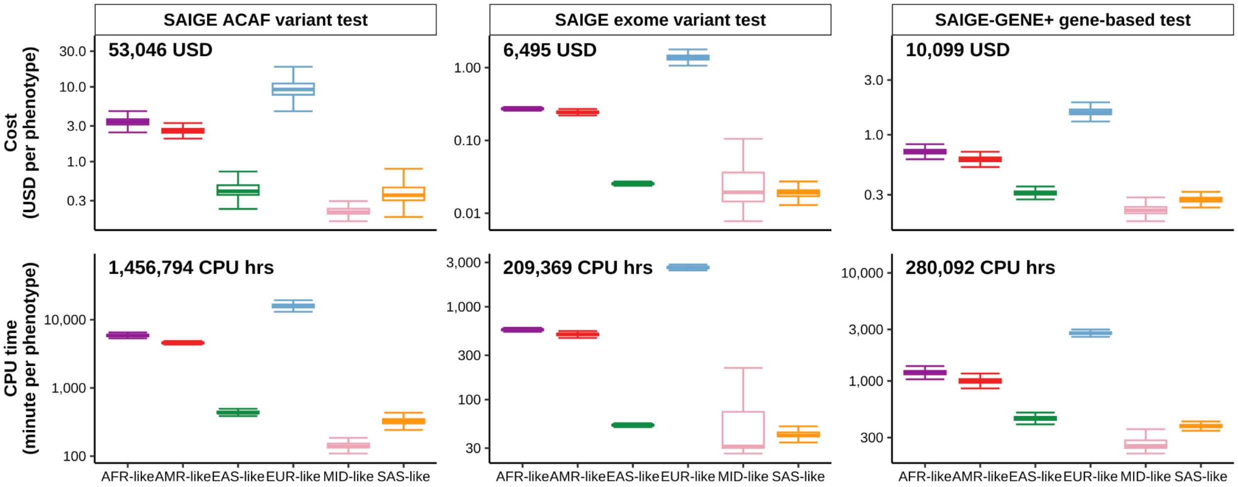


**Supplementary Figure 11** | Computational cost and runtime of SAIGE-based association pipelines. Distribution of per-phenotype computational cost (USD; top row) and CPU time (minutes; bottom row) for SAIGE and SAIGE-GENE+ Step 1 and Step 2 across genetic similarity groups. Columns denote association analyses (GWAS, ExWAS, and gene-level burden tests). Similarity groups are shown on the x-axis and by color. Total USDs and CPU hours across similarity groups are labeled on the top left and bottom left of the corresponding panels, respectively.

##### Result Format

We stored all association results in two Hail data formats: Hail Table (HT) and Hail MatrixTable (MT)[20]. In total, we generated 34,668 group-specific phenotype HTs, corresponding to three result types across 11,556 group-specific association tests. These phenotype-level HTs were subsequently merged hierarchically into similarity group-specific MTs, yielding 18 MTs in total (three result types across six similarity groups).

For each result type, we extracted summary statistics from the similarity group-specific MTs and combined them into a single MT with phenotypes represented as columns and association statistics stored as arrays in the entry fields. Using these three group-aggregated MTs, which encompassed 3,602 unique phenotypes represented across the 11,556 group-specific GWAS and RVAS analyses, we performed meta-analyses across up to six groups per phenotype, producing three meta-analysis MTs (**Supplementary Figure 12**). In parallel, we also generated meta-analysis results in HT format by directly meta-analyzing the individual similarity group-specific HTs.

In total, we provide 45,474 individual phenotype HTs, corresponding to all group-specific and meta-analyzed results across the three association types that take a total of about 95 TiB of storage, along with 21 MTs. The complete set of MTs occupies approximately 80 TiB of storage, including 69 TiB for ACAF-based GWAS results, 11 TiB for exome-wide association results, and 0.46 TiB for gene-level burden test results (**Supplementary Table 6**).

Gene-level Hail Tables (HTs) are keyed by phenoname, gene_id, gene_symbol, annotation, and max_MAF. The corresponding gene-level Hail MatrixTables (MTs) use the same key structure: columns contain phenotype-level metadata keyed by phenoname; rows represent gene groups keyed by gene_id, gene_symbol, annotation, and max_MAF; and entries store summary statistics for each phenotype × gene × annotation × max_MAF combination (e.g., *p* value, effect size β). Single-variant association HTs from ACAF and exome analyses are keyed by phenoname, locus, and alleles (GRCh38). The corresponding single-variant MTs follow the same schema, with columns containing phenotype metadata keyed by phenoname, rows representing variants keyed by locus and alleles (GRCh38), and entries storing summary statistics for each phenotype–variant pair (e.g., *p* value, effect size β). All datasets are publicly available through a browser-based framework and in Hail-native formats on the *All of Us* Researcher Workbench, with standardized filtering criteria, metadata, and functional annotations provided at the gene, variant, phenotype, and summary-statistics levels.

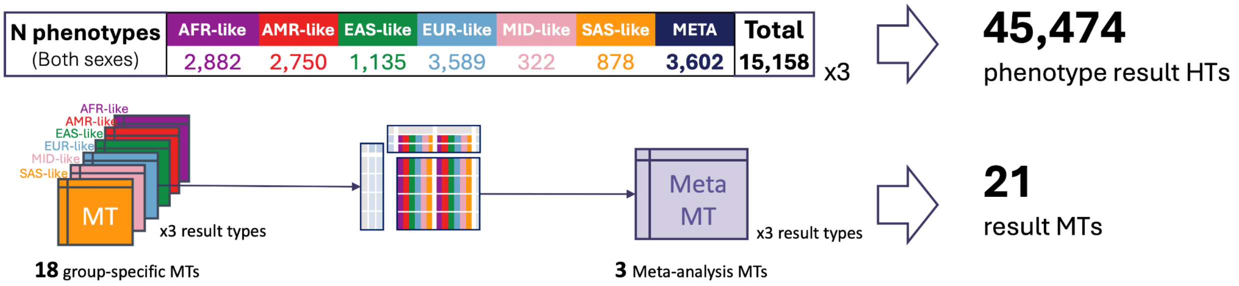


**Supplementary Figure 12** | Final result outputs and meta-analysis MatrixTable computation workflow. The upper panel shows the number of phenotypes analyzed for each genetic similarity group and in the meta-analysis. The lower panel illustrates the workflow for aggregating group-specific Hail MatrixTables and generating meta-analysis results.

| **Sizes of Hail Matrix Tables (MTs) and Hail Tables (HTs)** | | | | | | | | |
|---|---|---|---|---|---|---|---|---|
| | AFR-like | AMR-like | EAS-like | EUR-like | MID-like | SAS-like | META | **Total** |
| **ACAF variants** | 8.2 TiB | 7.69 TiB | 1.17 TiB | 10.54 TiB | 332.85 GiB | 1.1 TiB | 39.96 TiB | **68.99 TiB** |
| **Exome variants** | 1.21 TiB | 1.05 TiB | 117.47 GiB | 2.78 TiB | 14.06 GiB | 88.44 GiB | 6.03 TiB | **11.28 TiB** |
| **Gene** | 35.65 GiB | 31.38 GiB | 8.86 GiB | 46.33 GiB | 1.1 GiB | 4.84 GiB | 332.96 GiB | **461.12 GiB** |
| **Total** | 9.44 TiB | 8.77 TiB | 1.29 TiB | 13.37 TiB | 0.35 TiB | 1.19 TiB | 46.32 TiB | **80.72 TiB** |
| **Hail Table Size** | 16.33 TiB | 14.85 TiB | 2.31 TiB | 21.85 TiB | 586.53 GiB | 2.12 TiB | 36.8 TiB | **94.83 TiB** |
| **Number of Statistics** | | | | | | | | |
| | AFR-like | AMR-like | EAS-like | EUR-like | MID-like | SAS-like | META | **Total** |
| **Variants (ACAF + Exome)** | 228,624,754,292 | 219,399,735,500 | 42,885,820,640 | 340,456,606,337 | 10,992,981,790 | 36,156,823,176 | **454,893,766,838** | **1,333,410,488,573** |
| **Gene** | 715,750,464 | 681,741,500 | 267,410,540 | 900,681,084 | 47,041,946 | 201,714,354 | 911,723,832 | **3,726,063,720** |

**Supplementary Table 6** | Hail data sizes and association summary. The upper table reports sizes of Hail Matrix Tables (MTs) and Hail Tables (HTs), and the lower table reports numbers of association statistics, both stratified by genetic similarity groups and association types, along with counts for meta-analysis and overall total.

#### *QC of summary statistics*

##### Independent phenotypes

We further pruned to a set of relatively uncorrelated phenotypes. Using phenotypic information of samples post-QC, we generated a pairwise correlation table for 8,188 phenotypes using a matrix multiplication of the Hail MatrixTable and its transpose (**Supplementary Figure**

**13A**), and filtered the table to phenotype pairs with correlations ($r^2$) over 0.5. We then applied the *maximal_independent_set* function in Hail (https://hail.is/) to the remaining phenotype pairs with a tie-breaker function preferring phenotypes with more cases, resulting in a set of 2,502 related phenotypes to remove for the maximally independent phenotype set at $r^2$ <0.5 (**Supplementary Figure 13B**). Of these, 1,366 were included in our association analysis and are thus removed only when summarizing the count of approximately independent associations.

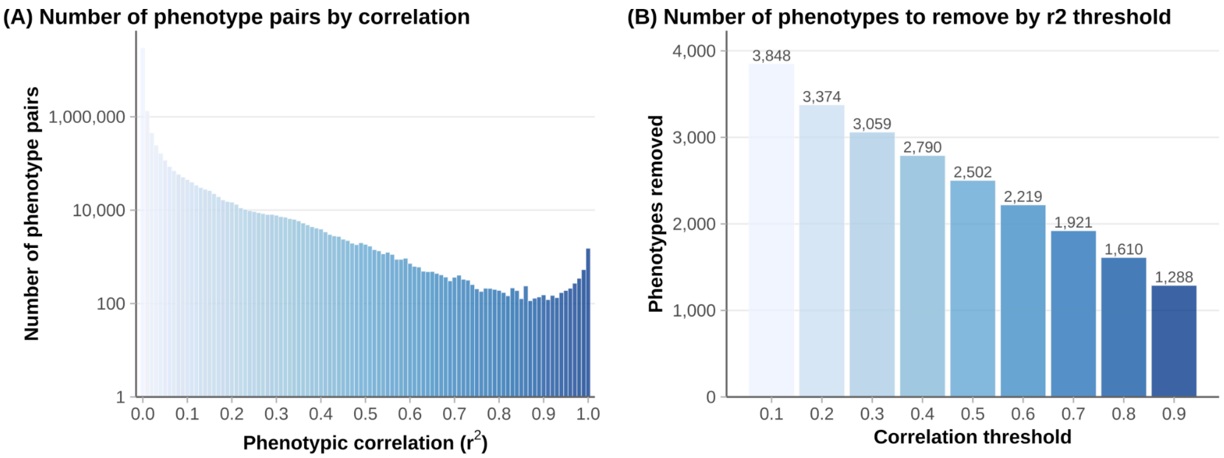


**Supplementary Figure 13** | Phenotype independence assessment. **A**, Histogram of phenotype pair counts (y-axis) by squared correlation ($r^2$; x-axis). **B**, Number of phenotypes removed (y-axis, and labeled on top of each bar) using the maximum independent set method across $r^2$ threshold (x-axis).

##### Simulated random phenotypes

To evaluate the asymptotic properties of the association tests, we adopted the simulation framework from Genebass. Using the genetic relationship matrix (GRM) described above, we simulated phenotypes from multivariate normal distributions using the sparseMVN package in R. Empirical heritability was fixed at 50%, and a total of 45 phenotypes were simulated, including both continuous traits and binary traits with varying prevalences. Binary phenotypes were generated with prevalences of 0.01%, 0.1%, 1%, 2%, 5%, 10%, 20%, and 50%, with five simulated phenotypes per prevalence level (**Supplementary Figure 14**).

Association testing showed good calibration for random phenotypes with prevalences > 1%, with burden test *p* value distributions closely following the null expectation and genomic control ($\lambda_{GC}$) values near 1. These results support our phenotype selection threshold requiring a group-specific case count of at least 200. In contrast, for rarer outcomes, we observed substantial deviations from the null, including inflated or deflated GC values, particularly in similarity groups with smaller sample sizes. These patterns likely reflect a combination of limited power, extreme case–control imbalance, sparse genotype–phenotype overlap, and breakdown of asymptotic assumptions and variance component estimation in gene-based mixed models under very small case counts (**Supplementary Figure 15**).

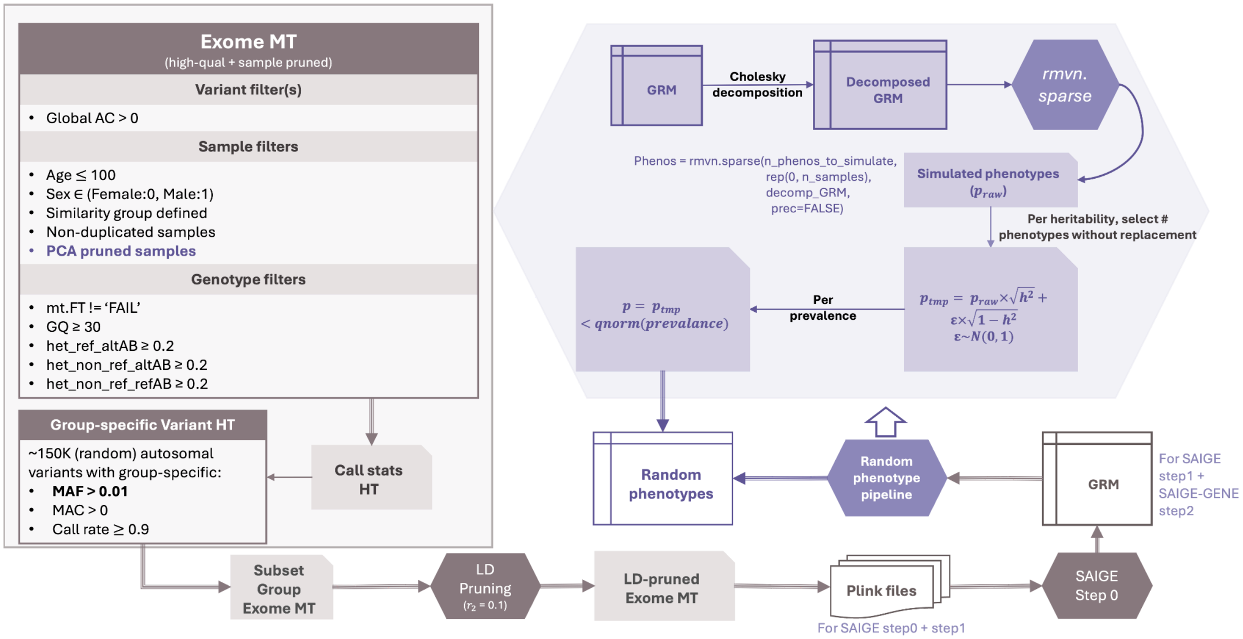


**Supplementary Figure 14** | Pipeline for generating group-specific PLINK files and sparse genetic GRMs for SAIGE step 1, with downstream random phenotype simulation. Genotype- and sample-QCed data from the Exome MT are used to select group-stratified autosomal variants across allele frequency bins, followed by LD pruning ($r^2 = 0.1$). GRMs are then computed and used to simulate random phenotypes with varying prevalence via the R package sparseMVN.

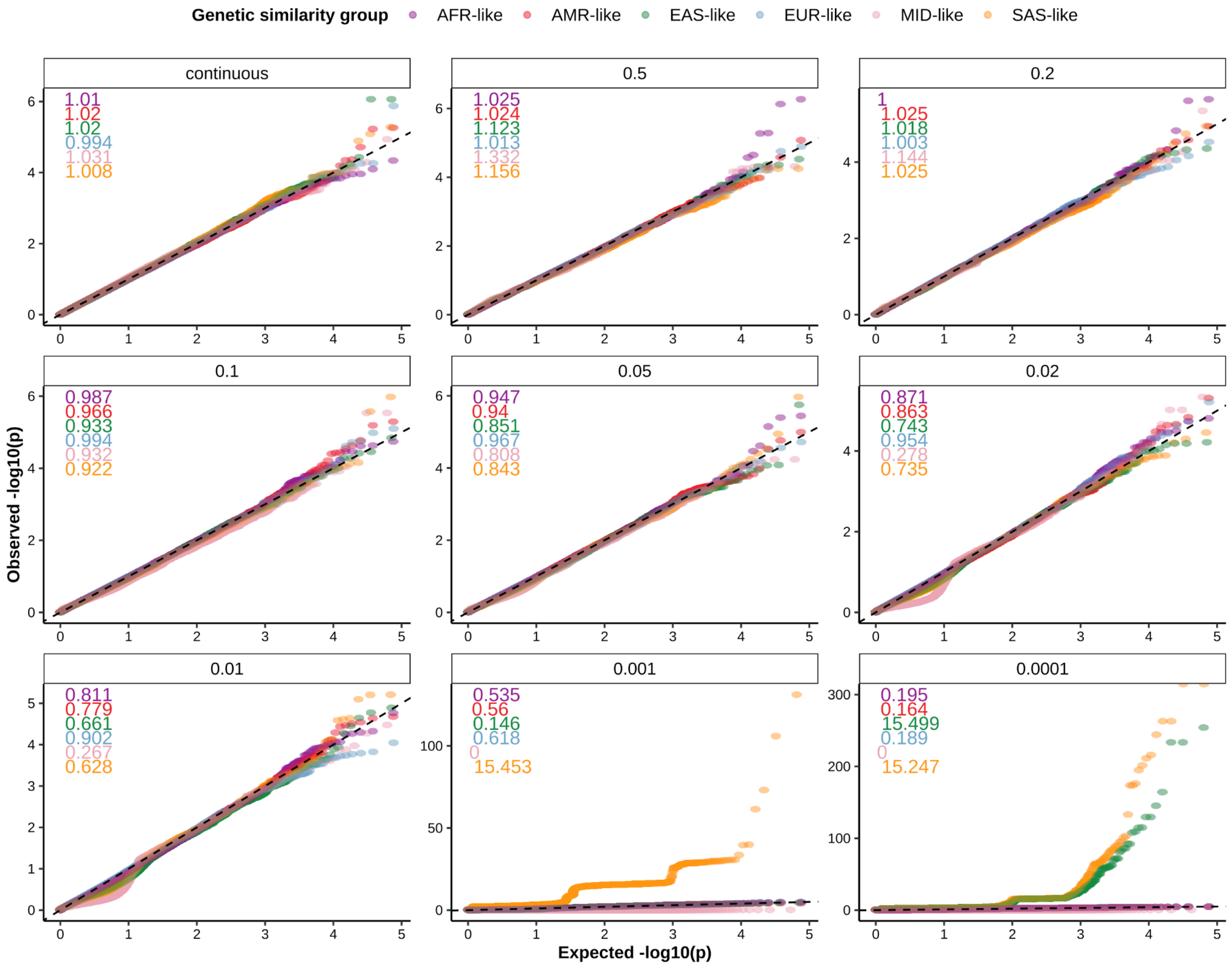


**Supplementary Figure 15** | Quantile–quantile (QQ) plots of gene-based burden association results for simulated phenotypes (maxMAF = 0.001). Each panel corresponds to a prespecified phenotype prevalence used in the simulations. Points represent gene–annotation pairs, and are colored by genetic similarity group. The x-axis shows expected $-\log_{10}$($p$ values) under the $\chi^2$ distribution, and the y-axis shows observed $-\log_{10}$($p$ values) from burden tests. The dashed line denotes the null expectation (y = x). Colored numbers in the upper left of each panel indicate the genomic inflation control factor ($\lambda_{GC}$) for the corresponding prevalence with text color matching the similarity group.

##### Empirical *p* value threshold

Using association results from simulated quantitative random phenotypes, we estimated empirical *p* value significance thresholds as 0.05 times the median of the minimum *p* values observed across five quantitative traits, evaluated separately for ACAF single-variant tests, exome single-variant tests, and gene-level burden, SKAT, and SKATO tests, across six similarity groups and their meta-analysis (**Supplementary Figure 16**). The empirical thresholds obtained for the meta-analysis ($p_{exome} = 1.2 \times 10^{-9}$, $p_{burden} = 2.3 \times 10^{-7}$, and $p_{SKATO} = 2.6 \times 10^{-7}$) were consistent with those previously reported by Genebass ($p_{exome} = 8 \times 10^{-9}$, $p_{burden} = 6.7 \times 10^{-7}$, and $p_{SKATO} = 2.5 \times 10^{-7}$). Given this close agreement, we adopted the Genebass-recommended gene-level significance threshold $6.7 \times 10^{-7}$ and the conventional genome-wide significance threshold of $5 \times 10^{-8}$ for single-variant analyses in this study, ensuring comparability with prior work.

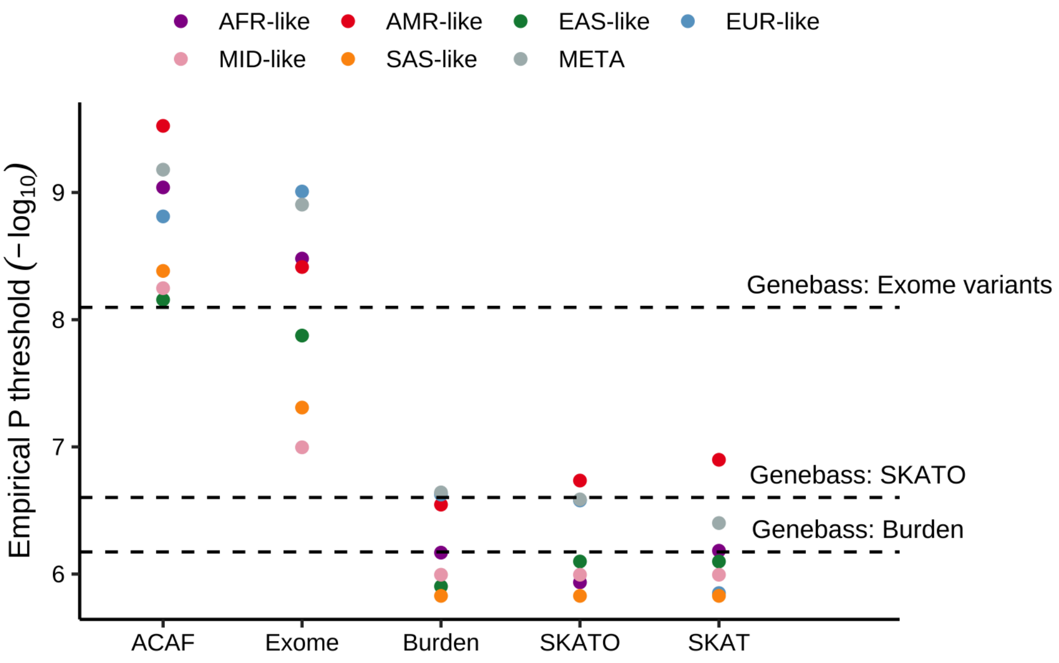


**Supplementary Figure 16** | Distribution of empirical *p* value significance thresholds. Each point represents a specific combination of association test type (GWAS, ExWAS, burden, SKAT, or SKATO; x-axis) and genetic similarity group (color). The y-axis shows negative $\log_{10}$ of the empirical *p* value thresholds estimated using random phenotypes from *All of Us.* The horizontal dashed line indicate the threshold computed from Genebass random phenotypes, with test types labeled at the upper right of each line.

#### Consistency of variant effect sizes on height across cohorts

We compared the ACAF GWAS summary statistics for height from *All of Us* v8 with results from gold-standard GWAS results from the GIANT consortium[21], across four similarity groups where data were available. The number of shared genome-wide significant associations ($p_{single\text{-}variant} < 5 \times 10^{-8}$) scaled with group-specific sample size, with the largest number of replicated associations observed in EUR-like (8,588; **Supplementary Table 7**). For both EUR-like and AFR-like, effect sizes for variants with significant height association in at least one cohort were highly concordant between *All of Us* v8 and GIANT, consistent with the scatter plot comparisons shown in **Supplementary Figure 17**. EAS-like and SAS-like were not included in effect size comparisons due to the limited number of overlapping variants passing frequency and significance filters.

| *AF > 0.001 in both* | AFR-like | EAS-like | EUR-like | SAS-like |
|---|---|---|---|---|
| **N variants in AoU** | **64,723,707** | **31,695,444** | **66,087,558** | **35,423,709** |
| Significant in AoU but not GIANT | 295 | 0 | 192 | 0 |
| Significant in both | 648 | 6 | 8,588 | 0 |
| Significant in GIANT but not AoU | 1,253 | 6,501 | 40,458 | 54 |
| **N variants in GIANT** | **1,373,215** | **1,299,805** | **1,372,528** | **1,280,311** |

**Supplementary Table 7** | GWAS height associations identified in *All of Us* v8 and GIANT across similarity groups. Variants were restricted to those present in both cohorts with allele frequency > 0.001. Bolded values in the top and bottom rows denote the total number of significant associations reported by each dataset, including those not observed in the other cohort.

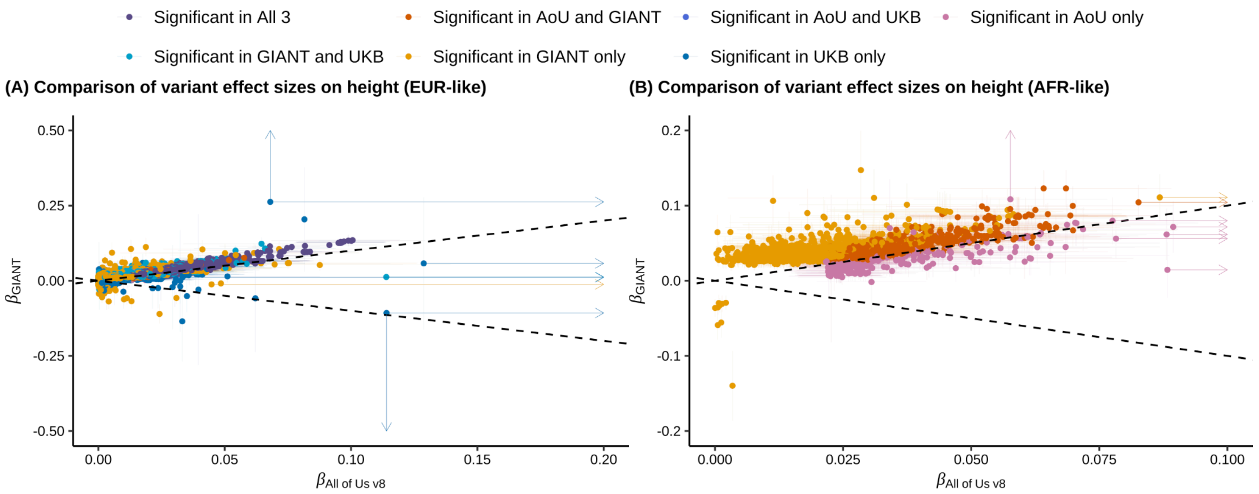


**Supplementary Figure 17** | Concordance of height effect sizes between *All of Us* v8 and GIANT across genetic similarity groups. Comparison of variant-level height effect sizes for **A**, EUR-like, and **B**, AFR-like. EAS-like and SAS-like groups are not shown due to limited overlap. Each point represents a variant, with *All of Us* v8 effect sizes on the x-axis and GIANT effect sizes on the y-axis. Effect sizes were oriented to the positive axis by aligning directions. Error bars denote 95% confidence intervals, with arrows indicating values extending beyond the plotting range. Dashed lines indicate concordance under y = x and y = −x. Point colors denote variant significance categories, defined by combining summary statistics from GIANT, *All of Us* v8, and pan-UK Biobank.

#### Consistency of burden effect sizes between *All by All* and Genebass

At the gene level, burden association results were consistent with previously reported rare variant burden associations from UK Biobank[16]. Burden effect sizes were concordant between Genebass and *All of Us* v8 EUR-like across phenotypes that could be mapped between the two cohorts (**Supplementary Figure 18A**), supporting the robustness and calibration of our association analyses. We also observed modestly stronger burden *p* value significance in Genebass compared to *All of Us* meta-analysis (**Supplementary Figure 18B**), indicating differences in statistical power attributable to sample size.

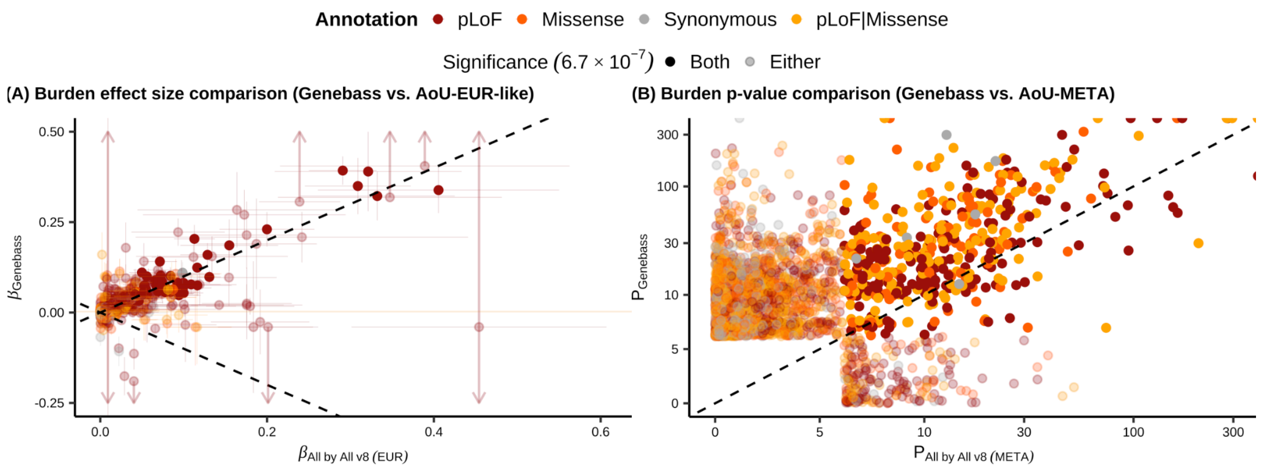


**Supplementary Figure 18** | Concordance of rare variant burden associations between *All by All* v8 and Genebass. **A**, Comparison of burden effect sizes between Genebass and EUR-like in *All by All* at maxMAF = 0.001, with effect sizes oriented to the positive axis. Error bars denote 95% confidence intervals, with arrows indicating values extending beyond the plotting range. **B,** Comparison of burden test *p* values between Genebass and *All by All* meta-analysis at maxMAF = 0.001. In both panels**,** each point represents a gene-level variant group, where color indicates functional annotation, and transparency indicates significance status across the two biobanks.

#### Expected *p* value ranking and lambda GC

Quantile-quantile (QQ) plots and the genomic control inflation factors ($\lambda_{GC}$) are essential for examining the calibration of GWAS results. QQ plots compare the distribution of observed *p* values against the expected distribution under the null hypothesis, while $\lambda_{GC}$ quantifies the degree of systematic inflation or deflation by comparing the median observed test statistic to its expected value. Deviations from the diagonal in QQ plots, particularly in the bulk of the distribution, or $\lambda_{GC}$ values substantially different from 1, can indicate issues related to population stratification, cryptic relatedness, technical artifact, and so on. Deviation in the tail of the QQ plot reflects true association signals.

To generate QQ plots for association results, expected *p* values based on the $\chi^2$ distribution are required for each variant based on its rank in the observed *p* value distribution. To avoid the computational burden of exact global sorting for datasets containing tens of millions of variants, we employed an approximate ranking strategy using the hl.agg.approx_cdf aggregator in Hail, which construct a streaming approximate cumulative distribution function of *p* values in O(N) time. Ranks were then interpolated for each variant based on its position within the CDF breakpoints, and expected *p* values were calculated as rank/(N+1), where N is the total number of variants. To ensure precision in the scientifically relevant tail of the distribution, variants in the top 1% by rank or with $p < 1 \times 10^{-3}$ were re-sorted exactly and assigned precise ranks before computing their expected *p* values. $\lambda_{GC}$ was computed by first estimating the median *p* value directly from the approximate CDF via interpolation at the 0.5 quantile, then applying the standard formula $\lambda_{GC} = \chi^2(p_{median}, \text{df} = 1)/\chi^2(0.5, \text{df} = 1)$, where $\chi^2$ denotes the chi-square distribution.

This approach avoids explicitly identifying the median variant through sorting or selection algorithms, instead leveraging the pre-computed CDF sketch, reducing computational complexity from O(N logN) to effectively O(N) for both QQ plot generation and $\lambda_{GC}$ calculation, which enables efficient quality control of GWAS summary statistics at scale with negligible loss of precision.

##### Result QC metrics

Using the approach described above, we compute $\lambda_{GC}$ at both the gene and single-variant levels, stratified by sequencing coverage bins and allele frequency (AF) or cumulative allele frequency (CAF) bins. As an initial QC step, we defined a phenotype-level flag (hq_phenotype) to exclude phenotypes with $\lambda_{GC} > 2$ in any analysis or with meta-analysis ACAF $\lambda_{GC} < 0.8$ for variant bin with AF > 0.1, which should present well-calibrated $\lambda_{GC}$ based on both sample size and variant frequency (**Supplementary Table 8**).

For gene-level analyses, we adopted the calibration framework from Genebass, computing gene-specific $\lambda_{GC}$ values across phenotypes with $\lambda_{GC} < 2$. Gene-level $\lambda_{GC}$ was strongly associated with mean sequencing coverage (approximated as the mean of variant_qc.gq_stats.mean / 3 across variants within each gene–annotation group), the number of contributing variants, and CAF. Based on these relationships, we defined hard filters to exclude genes with low coverage (hq_coverage_gene, coverage < 10) or few variants (hq_n_var_gene, < 5 variants). When computing final $\lambda_{GC}$ values for phenotypes included in the release, we further excluded gene-level tests with fewer than 10 variants and CAF < $1 \times 10^{-5}$ (or < $1 \times 10^{-4}$ for MID-like and SAS-like). We additionally applied an entry-level expected allele count filter (hq_exp_CAC), excluding gene–phenotype associations with expected allele count (CAF × number of cases, or CAF × number of non-missing individuals for quantitative traits) < 5. Genes with at least one phenotype passing this filter were flagged as hq_exp_CAC_gene. The gene-level QC flags (hq_coverage_gene, hq_n_var_gene, and hq_exp_CAC_gene) were combined into a single indicator (hq_gene), which, together with the entry-level hq_exp_CAC filter, defined the default criteria for retaining high-quality gene-level association results (**Supplementary Figure 19**, and **Supplementary Figure 20**). All gene-level QC metrics were computed using results with a maximum minor allele frequency (MAF) threshold of 0.001.

For single-variant analyses of both ACAF and exome variants, we similarly computed $\lambda_{GC}$ for each phenotype and summarized $\lambda_{GC}$ across variants within AF bins of [0%, 0.01%], (0.01%, 0.1%], (0.1%, 1%], (1%, 10%], and >10%. Given the increased instability of $\lambda_{GC}$ estimates for very rare variants, particularly for rare outcomes, we defined a variant-level filter (hq_AF_variant) to exclude variants with AF < $1 \times 10^{-4}$ when computing final $\lambda_{GC}$ values. We also applied an entry-level expected allele count filter (hq_exp_AC), excluding variant–phenotype associations with expected allele count (AF × number of cases, or AF × number of non-missing individuals for quantitative traits) < 5, and defined a variant-level flag (hq_exp_AC_variant) to retain variants with at least one phenotype passing this criterion. The variant-level QC flags (hq_AF_variant and hq_exp_AC_variant) were combined into a single indicator (hq_variant), which, together with the entry-level hq_exp_AC filter, defined the default criteria for retaining high-quality single-variant association results (**Supplementary Figure 19**, **Supplementary Figure 21**, and **Supplementary Figure 22**)

| Phenotypes with any $\lambda_{GC} > 2$ | | | | | | |
|---|---|---|---|---|---|---|
| Phenotype | Similarity Group | $N_{cases}$ | $N_{controls}$ | $\lambda_{GC_{ACAF}}$ | $\lambda_{GC_{Exome}}$ | $\lambda_{GC_{Gene}}$ |
| Uterine size date discrepancy (PP_928.1) | AFR-like | 285 | 59,706 | 0.55 | 0.04 | 0.71 |
| | AMR-like | 332 | 51,808 | 0.47 | 0.03 | 0.74 |
| | EUR-like | 272 | 175,890 | 34.41 | 0.99 | 47.37 |
| | META | 889 | 287,404 | 0.79 | 0.41 | 12.12 |
| Abnormality in fetal heart rate and rhythm complicating labor and delivery (PP_932.11) | AFR-like | 308 | 59,524 | 0.53 | 0.05 | 0.74 |
| | AMR-like | 293 | 51,566 | 0.43 | 0.02 | 0.73 |
| | EUR-like | 215 | 175,681 | 31.07 | 1.00 | 44.99 |
| | META | 816 | 286,771 | 0.77 | 0.22 | 9.76 |

| Phenotypes with meta-analysis ACAF $\lambda_{GC} < 0.8$ among variants with AF > 10% | | | |
|---|---|---|---|
| Phenotype | Description | N | $\lambda_{GC_{ACAF: AF > 10\%}}$ |
| 3011099 | Sex hormone binding globulin | 249 | 0.125 |
| 3046664 | Lipoprotein A [Moles/volume] in Serum or Plasma | 636 | 0.338 |
| 3013861 | LPA mass | 1,028 | 0.441 |
| 3052648 | Fibrin D-dimer FEU [Mass/volume] in Platelet poor plasma by Immunoassay | 1,646 | 0.736 |
| 3045792_3016921 | Smith antibody | 2,570 | 0.775 |

**Supplementary Table 8** | Phenotypes excluded during result QC. Phenotypes were excluded if the genomic control inflation factors ($\lambda_{GC}$) exceeded 2 in any analysis or if meta-analysis ACAF $\lambda_{GC}$ was < 0.8 within variant allele frequency bins with AF > 0.1.

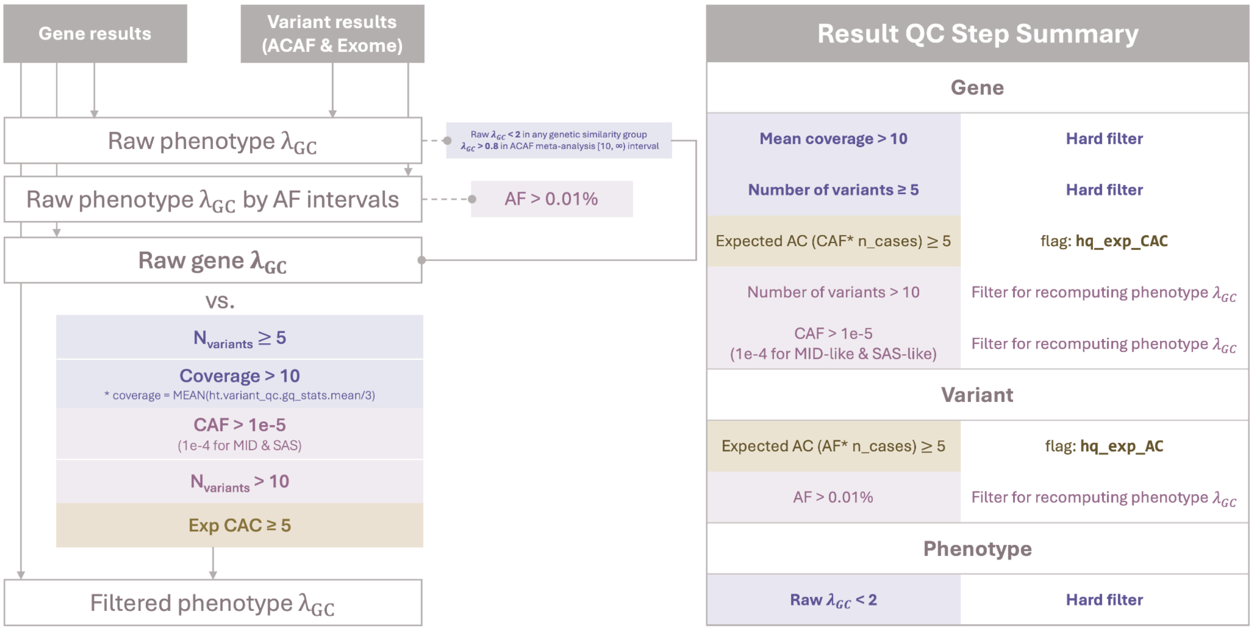


**Supplementary Figure 19** | Overview of QC pipeline applied to gene-level and single-variant association results. Genomic control inflation factors ($\lambda_{GC}$) were first computed per phenotype. Phenotypes were excluded if $\lambda_{GC}$ > 2 in any similarity group or if meta-analysis ACAF $\lambda_{GC}$ < 0.8 within the AF > 10% bin. For gene-level analyses, gene-specific $\lambda_{GC}$ values were summarized across well-calibrated phenotypes and evaluated against sequencing coverage, number of contributing variants, and cumulative allele frequency (CAF), motivating minimum thresholds for coverage, variant count, and CAF. Gene–phenotype associations were further filtered using an expected allele count criterion (CAF × number of cases ≥ 5). For single-variant analyses, $\lambda_{GC}$ was summarized across allele frequency bins, rare variants with unstable $\lambda_{GC}$ estimates were excluded, and variant–phenotype associations were filtered using an expected allele count threshold (AF × number of cases ≥ 5).

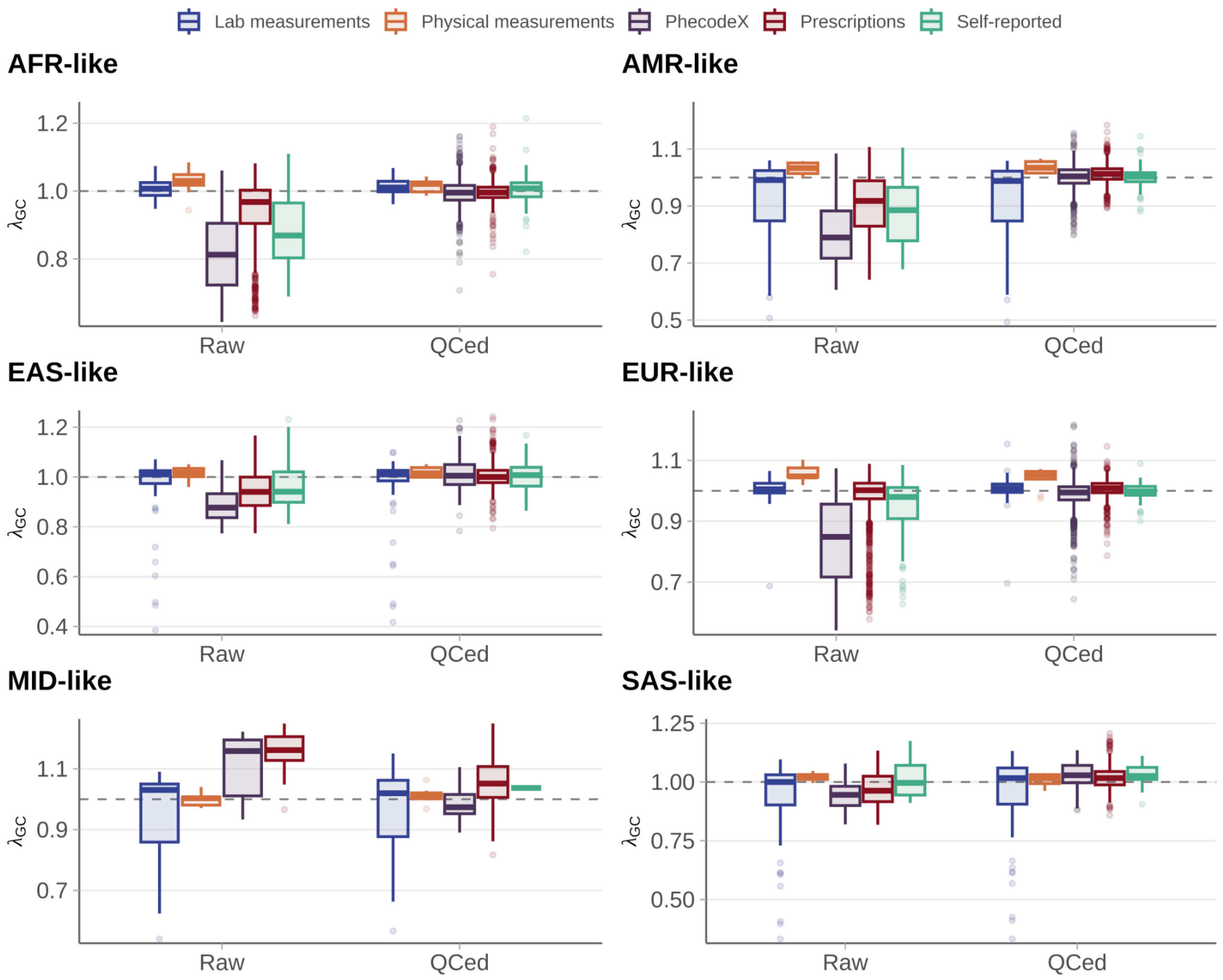


**Supplementary Figure 20** | $\lambda_{GC}$ before and after QC. $\lambda_{GC}$ (y-axis) for gene-level burden association results before and after QC filtering (x-axis), stratified by similarity group (panels) and phenotype category (colored box plots). The horizontal dashed line denotes the null expectation ($\lambda_{GC}$ = 1).

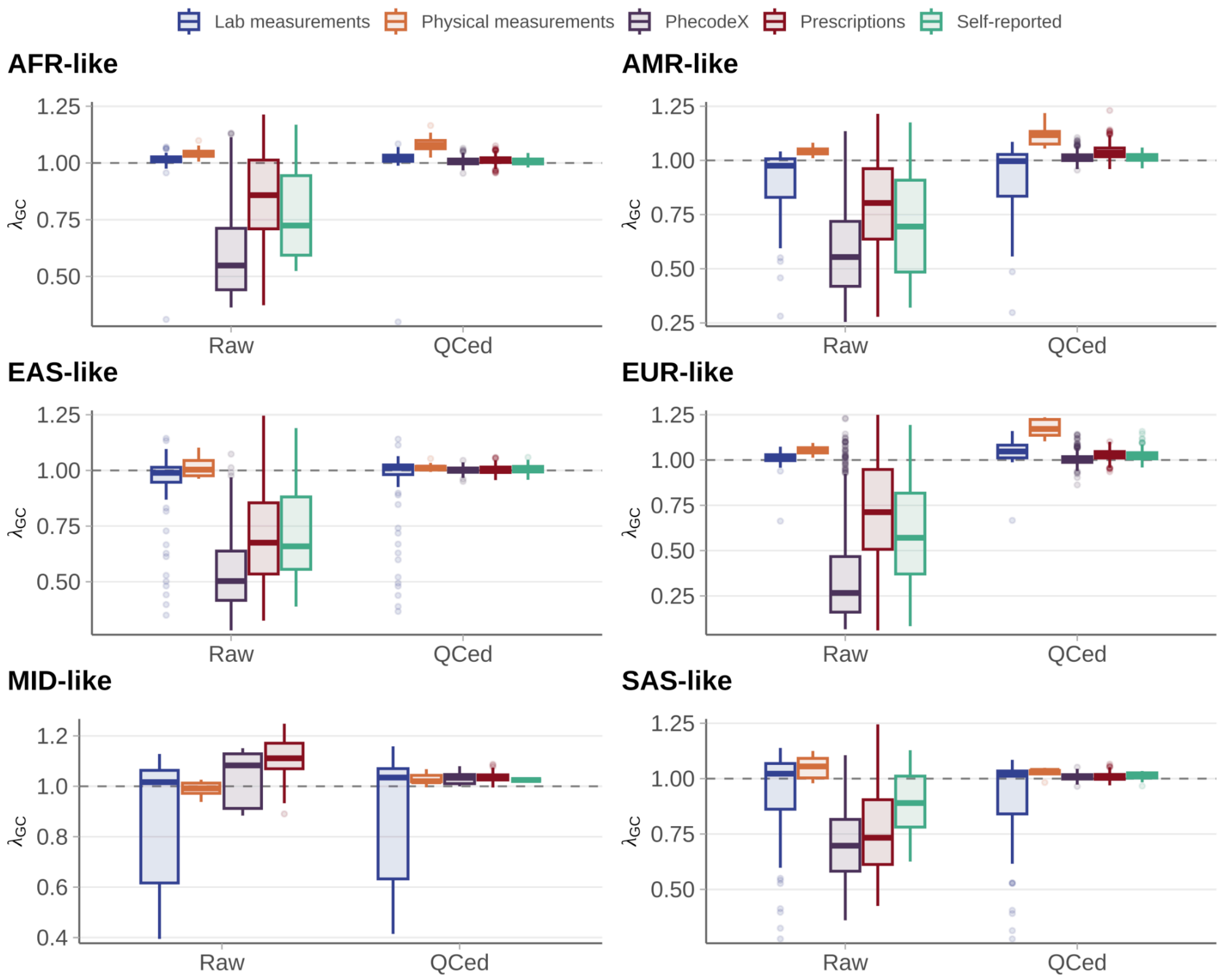


**Supplementary Figure 21** | $\lambda_{GC}$ before and after AC. $\lambda_{GC}$ (y-axis) for ACAF single-variant association results before and after QC filtering (x-axis), stratified by similarity group (panels) and phenotype category (colored box plots). The horizontal dashed line denotes the null expectation ($\lambda_{GC}$ = 1).

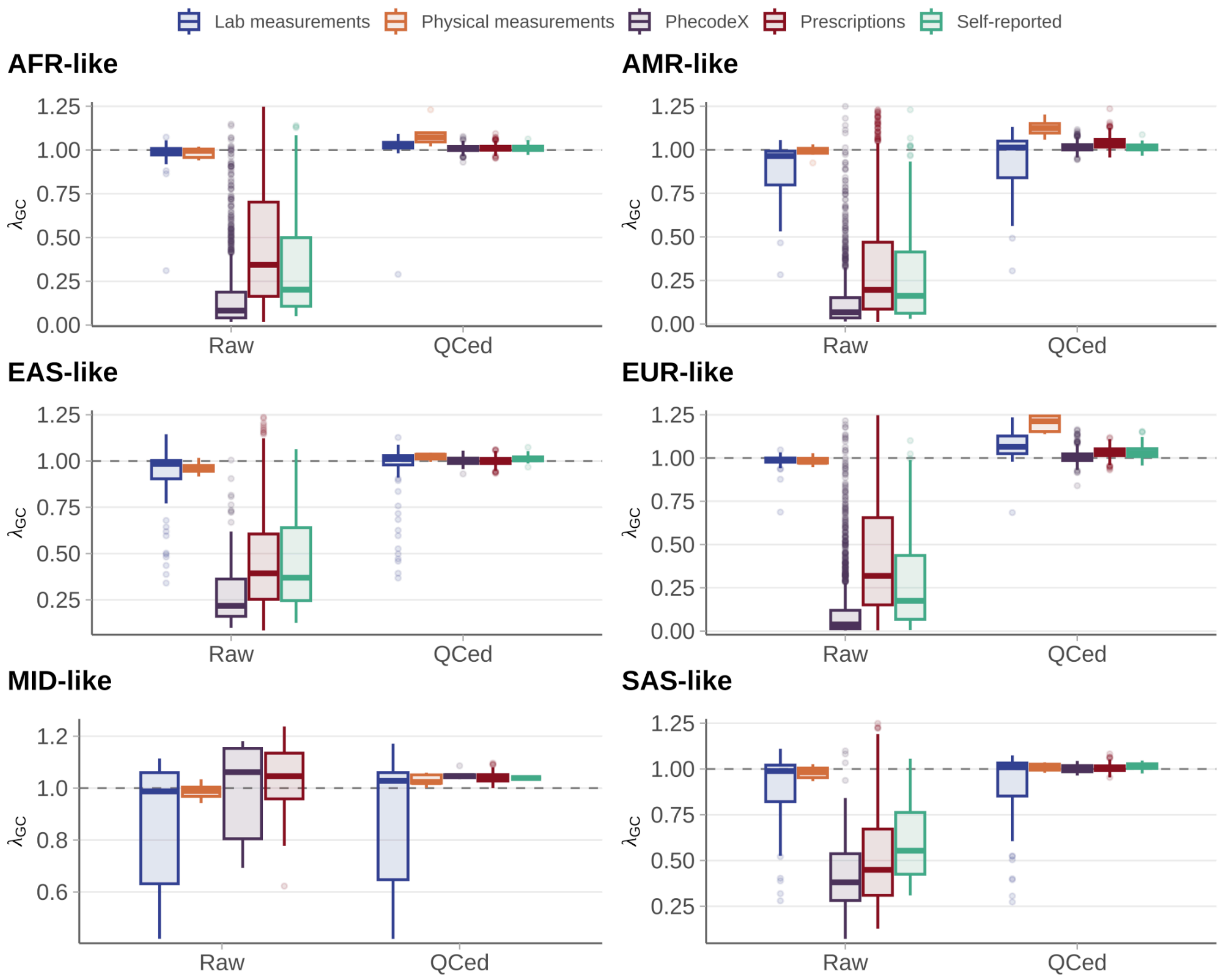


**Supplementary Figure 22** | $\lambda_{GC}$ before and after QC. $\lambda_{GC}$ (y-axis) for Exome single-variant association results before and after QC filtering (x-axis), stratified by similarity group (panels) and phenotype category (colored box plots). The horizontal dashed line denotes the null expectation ($\lambda_{GC}$ = 1).

#### *Analysis of summary statistics*

##### Summary of significant associations

We performed 1,333,410,488,573 single-variant association tests across variants from the Allele Count/Allele Frequency (ACAF) and exome call sets in the *All of Us* Research Program, together with 3,726,063,720 gene-level association tests (**Supplementary Table 6**). After results QC and restricting to a maximally independent phenotype set (phenotypic $r^2 < 0.5$), we identified 48,831 relatively independent genome-wide significant single-variant associations (LD-pruned at $r^2 < 0.1$; $p_{\text{single-variant}} < 5 \times 10^{-8}$) and 1,480, 1,032, and 789 significant gene-level burden associations ($p_{\text{gene-burden}} < 6.7 \times 10^{-7}$) at maximum MAF thresholds of 1%, 0.1%, and 0.01%, respectively (**Supplementary Table 9**, and **Supplementary Data 1**).

The distributions of association counts across phenotypes showed similar patterns across genetic similarity groups, with differences in the rate of saturation across phenotypes reflecting the relative magnitude of sample sizes. Owing to the substantially increased statistical power from aggregating samples across genetic similarity groups, the meta-analysis identified more associations per phenotype at both single-variant level and gene-level. Approximately 9% of the phenotypes had ≥ 10 single-variant associations (**Supplementary Figure 23A**), and around 3% of the phenotypes having ≥ 3 gene-level burden associations (maxMAF = 0.1%) (**Supplementary Figure 23B**).

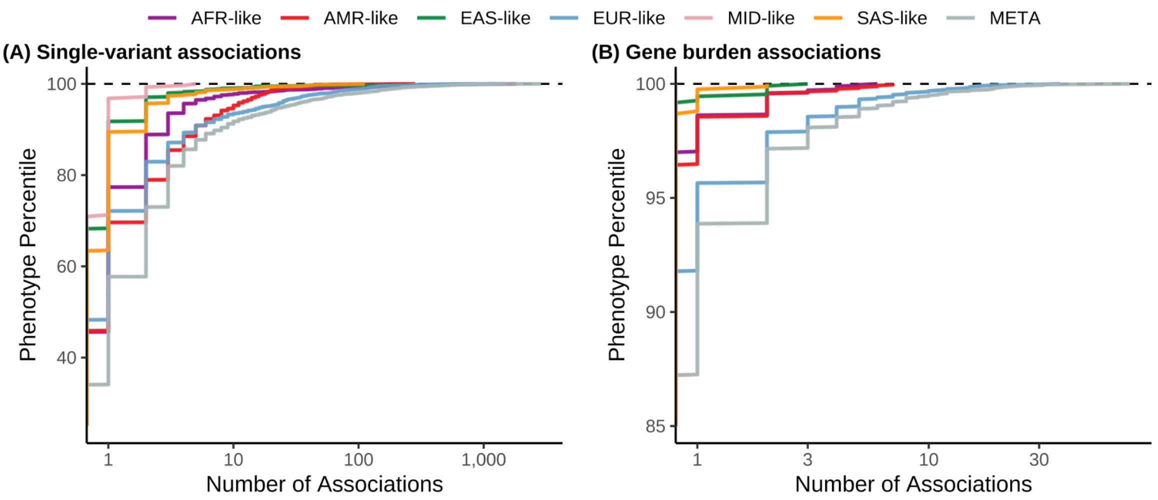


**Supplementary Figure 23** | Cumulative distribution of significant associations across phenotypes by similarity group. The y-axis shows the cumulative phenotype percentile of significant association counts (x-axis) across phenotypes within each genetic similarity group (color). **A**, Single-variant associations combining both ACAF and Exome results, after QC and LD pruning using PLINK clumping ($r^2 < 0.1$). **B**, Gene-level burden associations, after QC with maxMAF = 0.001.

| Similarity Group | Single-variant | Gene (Burden test; max MAF = 0.001) | | | | | Total |
|---|---|---|---|---|---|---|---|
| | | pLoF | missense | synonymous | pLoF + missense | Total | |
| AFR-like | **9,682** | 26 | 25 | 9 | 32 | **92** | **9,774** |
| AMR-like | **3,152** | 27 | 29 | 10 | 49 | **115** | **3,267** |
| EAS-like | **2,561** | 1 | 3 | 1 | 4 | **9** | **2,570** |
| EUR-like | **14,803** | 215 | 139 | 14 | 203 | **571** | **15,374** |
| MID-like | **49** | 0 | 0 | 0 | 0 | **0** | **49** |
| SAS-like | **431** | 0 | 2 | 1 | 6 | **9** | **440** |
| META | **26,571** | 362 | 191 | 31 | 292 | **876** | **27,447** |
| **Unique** | **48,831** | **376** | **239** | **64** | **353** | **1,032** | **49,863** |
| **Total** | **57,249** | **631** | **389** | **66** | **586** | **1,672** | **58,921** |

**Supplementary Table 9 |** Summary of significant associations after result QC. Shown are numbers of QCed significant association across similarity groups, including the meta-analysis. Results are reported for LD-pruned ($r^2 < 0.1$) single-variant associations (ACAF and Exome), and for gene-level burden tests, stratified by the four functional annotation categories. The bottom row ("Total") reports the naive sum of association counts across similarity groups after result QC, whereas the penultimate row ("Unique") reports the number of unique associations after accounting for overlap across similarity groups and restricting to phenotypes with squared correlation $r^2 < 0.5$.

**Contribution of similarity groups to associations identified from meta-analysis**

We quantified the fraction contribution of each similarity group $i$ to the meta-analysis test statistics independent of sample size, defined as $\sqrt{2CAF_i(1 - CAF_i)}(-\Phi^{-1}(p_i))\times sign(\beta_i)$, corresponding to the unweighted Z-score component used in the Stouffer method[18] for meta-analysis, where $CAF_i$, $p_i$ and $\beta_i$ are the *p* value and effect sizes for similarity group $i$. In contrast to **Figure 2b**, which presents the raw magnitude of similarity group-specific Z-scores, **Supplementary Figure 24** shows the relative proportion of similarity group-specific Z-scores contributing to each meta-analysis association. These proportions reflect both the relative representation of each similarity group and strength of their genetic signals. The average contribution of each similarity group, defined as its proportion of the total unweighted Z-score, demonstrates strong concordance with the corresponding group-specific sample size proportions (**Supplementary Figure 25**).

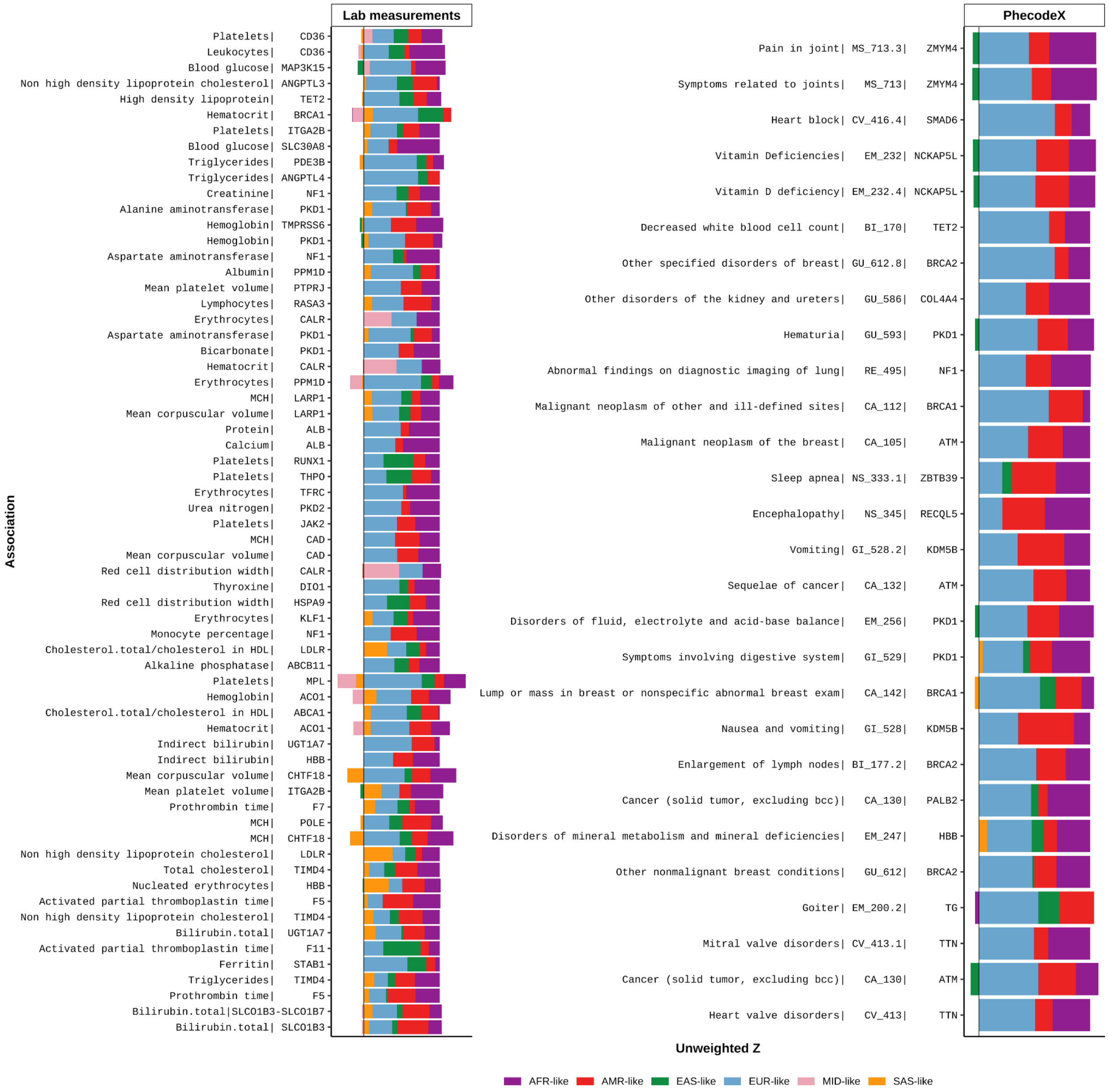


**Supplementary Figure 24** | Similarity contributions to meta-analysis gene-level burden associations. Proportion of the unweighted Z-score contributing to meta-analysis associations, defined as $\sqrt{2CAF_i(1-CAF_i)}\,(-\Phi^{-1}(p_i))\times sign(\beta_i)$, where $CAF_i$, $p_i$ and $\beta_i$ are the combined allele frequency (CAF), *p* value and beta effect size from the pLoF burden test for similarity group $i$ (color). Each bar represents a significant association from the meta-analysis ($p_{gene\text{-}burden} <$ $6.7\times10^{-7}$), for which none of the component similarity groups presented significance. Colored segments indicate the proportional contribution of each similarity group to the total unweighted Z score chunks.

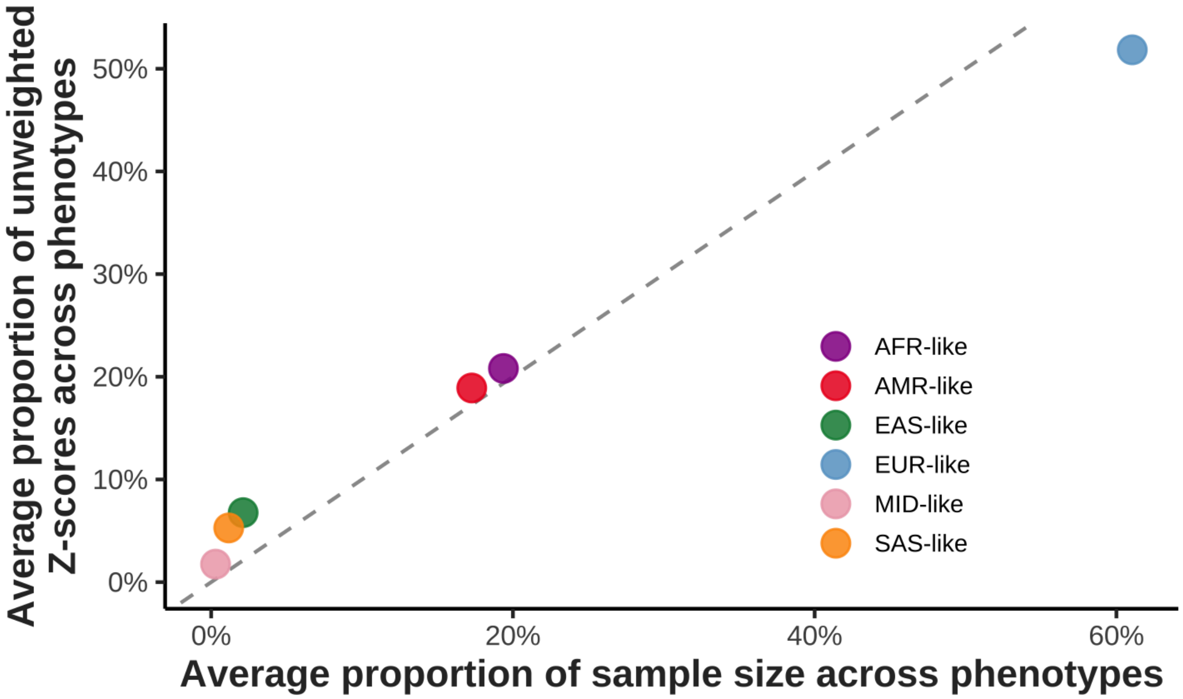


**Supplementary Figure 25** | Contribution of similarity groups to association signals relative to sample size. Relationship between the average unweighted Z-score contribution of each similarity group (x-axis), defined as the proportion of the total unweighted Z-score, and the corresponding average sample size across phenotypes (y-axis). Each point represents a similarity group. The dashed line denotes the identity line (y = x).

##### *Novelty assessment pipeline for gene-phenotype associations*

###### Overview

We developed an agentic framework to assess the novelty of discovered gene-phenotype associations. The pipeline integrates two complementary data sources: PubMed literature and the Open Targets platform. This dual approach addresses inherent limitations in each source: PubMed searches are restricted to titles and abstracts, where association study results are rarely reported, while Open Targets may miss mechanistic studies, case reports, and animal model evidence that might appear only in non-GWAS literature. For each queried association, the agentic pipeline parses the biomedical literature and integrates Open Targets association scores to assign a level of prior evidential support (not found, hypothesized, existing, or established; **Supplementary Figure 26** and **Supplementary Figure 27**). By standardizing the evaluation of prior knowledge, this tool facilitates identification of novel and biologically meaningful associations and accelerates translation from large-scale association analyses to biological insight.

The pipeline receives a gene name and phenotype name as input and returns two independent verdicts—one from the literature analysis and one from Open Targets—each classified as: Not Found, Hypothesized, Existing, or Established. All agents were implemented using Claude Haiku 4.5 via the Anthropic API in Python.

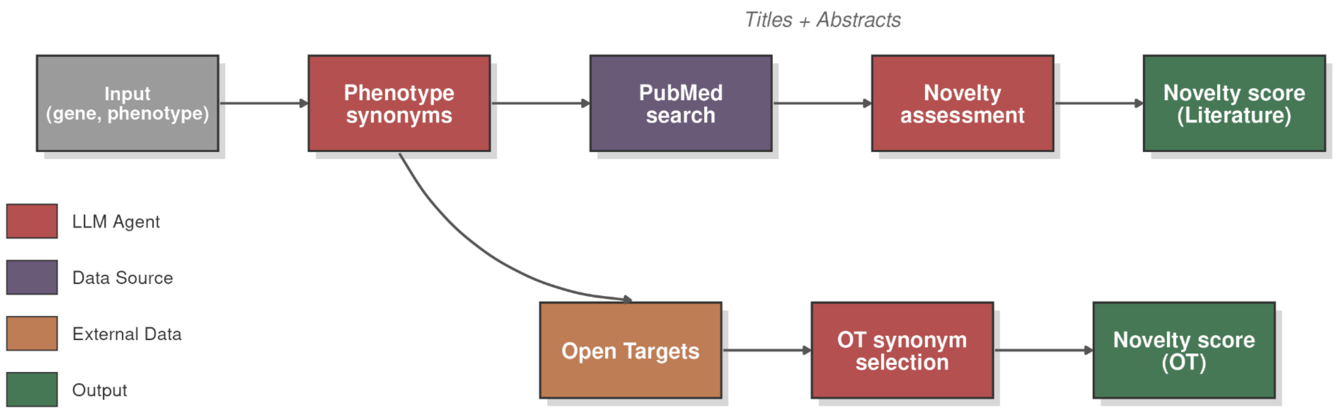


**Supplementary Figure 26** | Schematic representation of the novelty assessment agentic pipeline.

#### Literature Analysis

##### Synonym Generation

Effective PubMed searching requires accounting for nomenclature variability. Phenotypes may be referenced by different terms across publications (e.g., "height" versus "stature," or "Type 2 Diabetes" versus "Diabetes Mellitus"), and genes often have multiple aliases. Gene synonyms were retrieved directly from the HGNC database. For phenotypes, we developed a "Phenotype Synonyms" agent that generates synonyms for a given phenotype term. To maximize coverage of the synonym space, we invoked this agent four times with varying temperature parameters (0, 0.1, 0.2, 0.3) and concatenated the deduplicated results.

##### PubMed Search Strategy

Two complementary search strategies were used:

1. Association search. This targeted search identifies articles mentioning both the gene and the phenotype. The query structure combines the gene name (in title or abstract) OR phenotype terms and their synonyms (in title or abstract). Articles were retrieved in order of relevance as ranked by PubMed. We collected up to 30 articles, iterating through gene synonyms from HGNC if the initial search returned fewer results.

```
gene_name = "GENE"
phenotype_terms = ["phenotype", "pheno_syn1", "pheno_syn2"]  #
etc.

gene_clause_main = f'"{gene_name}"[Title/Abstract]'

association_query = " OR ".join([f'"{term}"[Title/Abstract]'
for term in phenotype_terms])
```

2. Agnostic search. This broader search captures general gene-related publications by combining the gene name with generic disease-related keywords ("phenotype," "disease," "syndrome," "association," "trait") without specifying the phenotype of interest. Up to 20 articles were retrieved using the same synonym iteration strategy.

```
gene_clause_main = f'"{gene_name}"[Title/Abstract]'

agnostic_keywords = (
   '"phenotype"[Title/Abstract] OR "disease"[Title/Abstract] OR
"syndrome"[Title/Abstract] '
   'OR "association"[Title/Abstract] OR
"trait"[Title/Abstract]'
)

agnostic_query = f'(({gene_clause_main})) AND
({agnostic_keywords})'
```

The final article set was generated by combining and deduplicating results from both searches.

##### Novelty Classification

Retrieved article titles and abstracts were submitted to a "Novelty Assessment" agent, which classified the association novelty based on the available literature evidence. The agent was instructed to provide a justification and cite supporting articles for its verdict.

##### Open Targets Analysis

###### Association Score Retrieval and Filtering

For each gene, we queried the Open Targets API to retrieve association scores (ranging from 0 to 1) for all linked phenotypes. Scores below 0.05 were excluded as background noise.

###### Phenotype Matching

Phenotype matching proceeded in two stages. First, we performed exact string matching between the queried phenotype (and its synonyms from the Phenotype Synonyms agent) and the phenotypes in the Open Targets results. Second, an "OT Synonym Selection" agent identified up to 10 additional phenotypes from the Open Targets list that were either synonymous with or closely related to the queried phenotype. Here, "closely related" denotes a weaker relationship than synonymy (e.g., "obesity" is closely related to "BMI," whereas "Diabetes Mellitus" is synonymous with "Type 2 Diabetes").

###### Score-Based Classification

The maximum association score among all matched phenotypes was used to assign a novelty label:

- Not Found: No phenotypes matched, or no scores exceeded 0.05
- Hypothesized: Maximum score between 0.05 and 0.2
- Existing: Maximum score between 0.2 and 0.5
- Established: Maximum score above 0.5

##### Verdict Integration

The final novelty assessment combined verdicts from both sources by taking the maximum (most established) classification, adopting a conservative approach to novelty claims. This integration strategy ensures that associations with strong evidence in either source are appropriately recognized, while acknowledging that each source captures different types of evidence. In our application, Open Targets tended to identify more associations—consistent with GWAS results being underreported in abstracts—while the literature analysis uniquely captured mechanistic studies or case studies that provide biological context for the associations.

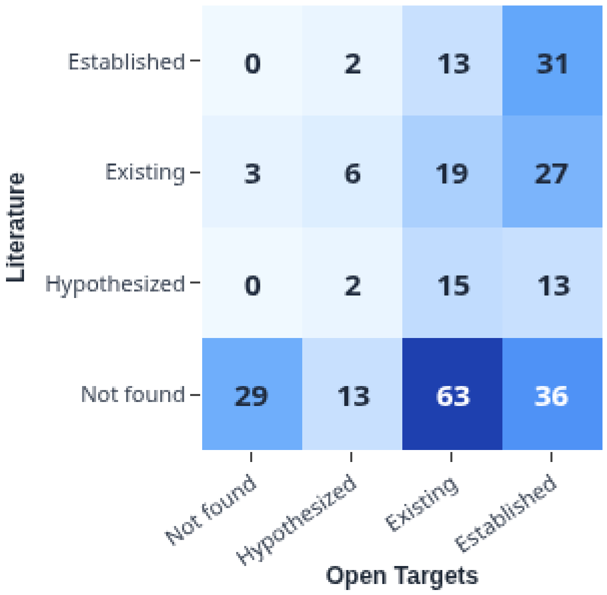


**Supplementary Figure 27** | Confusion matrix between the Literature agent scores and Open Targets agent scores across 272 associations significant in only cross-biobank meta-analysis, not in either biobank alone.

Particularly, here we applied this pipeline to the 193 tentative novel pLoF associations identified only in cross-biobank gene-level burden meta-analysis (AoU maxMAF = 0.001) but not any of the individual biobanks, where we highlighted 5 pLoF-disease associations (**Figure 4b**), 12 pLoF-lab measurement associations (**Supplementary Figure 28**) , and 5 pLoF-physical measurement associations (**Supplementary Figure 29**) that have no supporting evidence from existing literatures. For cross-biobank meta-analysis with AoU maxMAF = 0.01, the number of tentative novel pLoF associations is 190, with 32 involving disease outcomes, where we highlighted 4 pLoF-disease associations, 12 pLoF-lab measurement associations, and 5 pLoF-physical measurement associations with no previous evidence. For the potentially novel signals highlighted in the main text, no exceptionally strong common variant association peak ($p_{single\text{-}variant} < 10^{-20}$) was observed in the corresponding genomic regions, suggesting these signals are unlikely to be driven by nearby common variants with strong LD. By combining interactive visualization with transparent evidence summaries, the browser extends the novelty assessment framework from a back-end agentic pipeline into a practical discovery-support tool for large-scale GWAS and RVAS studies.

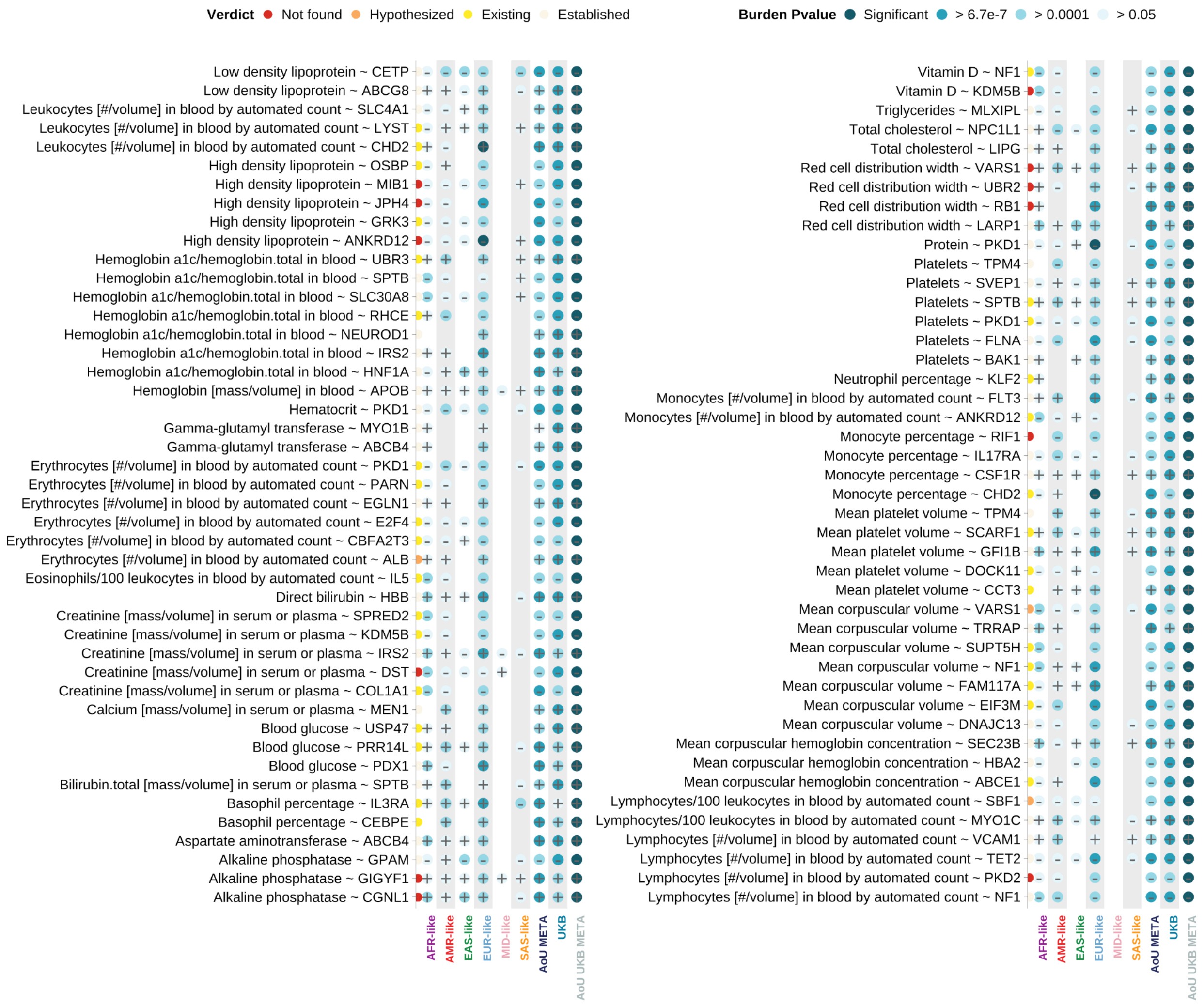

**Supplementary Figure 28** | Rare pLoF burden associations with laboratory measurements identified only through cross-biobank meta-analysis. 89 associations (y-axis) are significant in the meta-analysis of UK Biobank and *All of Us*, but not in any individual genetic similarity groups or biobanks (x-axis), with 12 classified as potentially novel (red points). Red sequentially colored dots adjacent to each association indicate AI-assisted evidence grading, reflecting the level of proper support in scientific literatures. Blue sequentially colored dots with sign labels represent cohort-specific burden test *p* value magnitude (color intensity), and effect directions (sign).

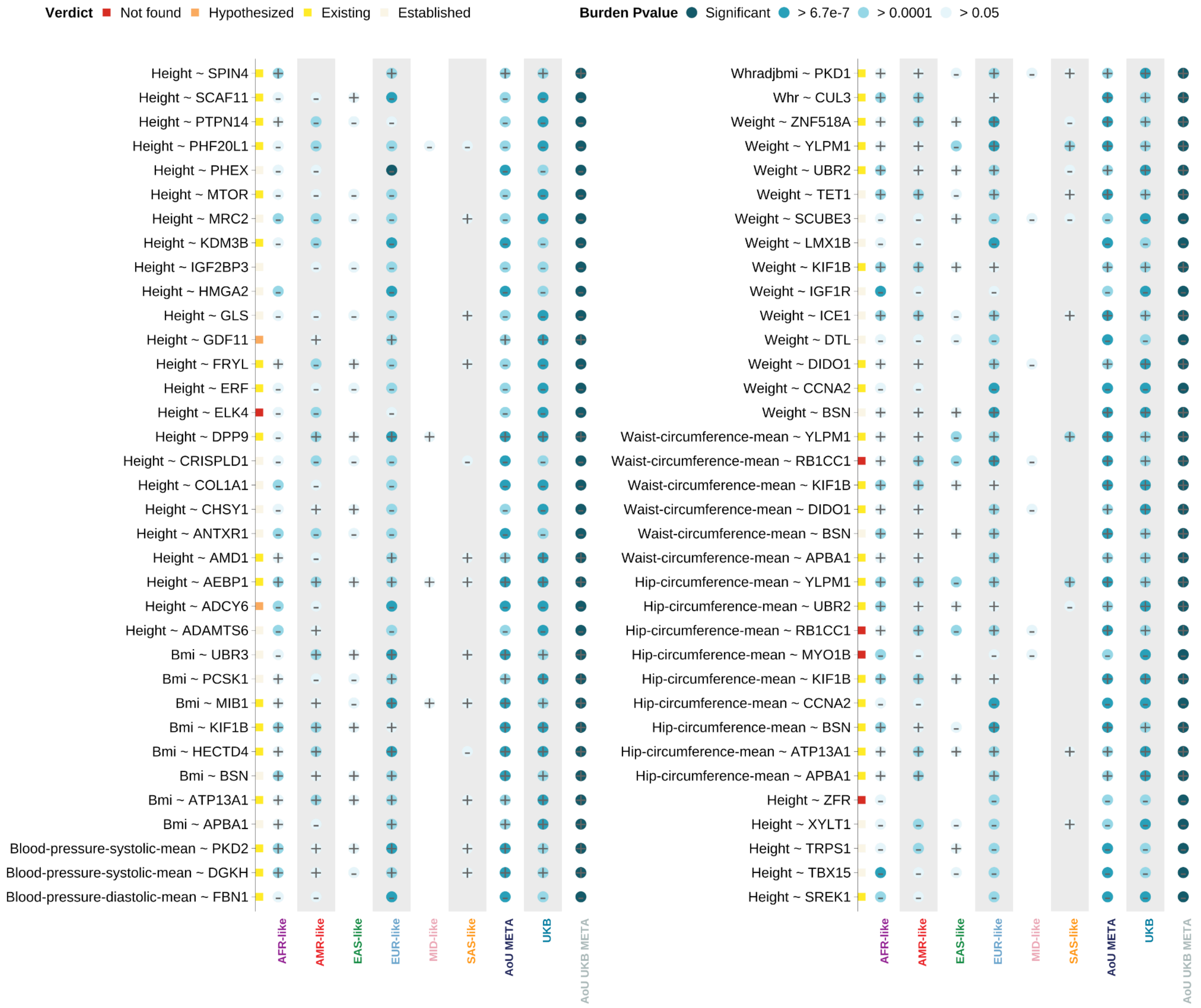


**Supplementary Figure 29** | Rare pLoF burden associations with physical measurements identified only through cross-biobank meta-analysis. 70 associations (y-axis) are significant in the meta-analysis of UK Biobank and *All of Us*, but not in any individual genetic similarity groups or biobanks (x-axis), with 5 classified as potentially novel (red points). Red sequentially colored dots adjacent to each association indicate AI-assisted evidence grading, reflecting the level of proper support in scientific literatures. Blue sequentially colored dots with sign labels represent cohort-specific burden test *p* value magnitude (color intensity), and effect directions (sign).

*References*


1. The All of Us Research Program Genomics Investigators. Genomic data in the All of Us Research Program. *Nature* **627**, 340 (2024).
2. Karczewski, K. J. *et al.* The mutational constraint spectrum quantified from variation in 141,456 humans. *Nature* **581**, 434–443 (2020).
3. Chen, S. *et al.* A genomic mutational constraint map using variation in 76,156 human genomes. *Nature* **625**, 92–100 (2024).
4. Cavalli-Sforza, L. L. The Human Genome Diversity Project: past, present and future. *Nature reviews. Genetics* **6**, (2005).
5. A global reference for human genetic variation. *Nature* **526**, 68–74 (2015).
6. National Academies of Sciences, Engineering, and Medicine. *Using Population Descriptors in Genetics and Genomics Research: A New Framework for an Evolving Field*. (National Academies Press, 2023).
7. Shuey, M. M. *et al.* Next-generation phenotyping: introducing phecodeX for enhanced discovery research in medical phenomics. *Bioinformatics* **39**, btad655 (2023).
8. Brogan, J., Waxse, B., Corty, R. W., He, M. K., Poisner, H., Sharber, B., Kouame, A., Qian, J., Denny, J. C., Bick, A., & Carroll, R. J. Unifier: multi-center unit harmonization and quality control pipeline for laboratory measurements. *[Manuscript in preparation]* **. Department of Biomedical Informatics, Vanderbilt University Medical Center**,.
9. Karczewski, K. J. *et al.* Pan-UK Biobank genome-wide association analyses enhance discovery and resolution of ancestry-enriched effects. *Nature Genetics* **57**, 2408–2417 (2025).
10. Karczewski, K. J. *et al.* Pan-UK Biobank GWAS improves discovery, analysis of genetic architecture, and resolution into ancestry-enriched effects. *medRxiv* 2024.03.13.24303864 (2024) doi:10.1101/2024.03.13.24303864.

11. Zhou, W. *et al.* Efficiently controlling for case-control imbalance and sample relatedness in large-scale genetic association studies. *Nat. Genet.* **50**, 1335–1341 (2018).
12. Zhou, W. *et al.* Scalable generalized linear mixed model for region-based association tests in large biobanks and cohorts. *Nat. Genet.* **52**, 634–639 (2020).
13. Zhou, W. *et al.* Set-based rare variant association tests for biobank scale sequencing data sets. *bioRxiv* (2021) doi:10.1101/2021.07.12.21260400.
14. Wu, M. C. *et al.* Rare-Variant Association Testing for Sequencing Data with the Sequence Kernel Association Test. *American Journal of Human Genetics* **89**, 82 (2011).
15. Lee, S. *et al.* Optimal unified approach for rare-variant association testing with application to small-sample case-control whole-exome sequencing studies. *American journal of human genetics* **91**, (2012).
16. Karczewski, K. J. *et al.* Systematic single-variant and gene-based association testing of thousands of phenotypes in 394,841 UK Biobank exomes. *Cell Genom* **2**, 100168 (2022).
17. Zhou, W. *et al.* SAIGE-GENE+ improves the efficiency and accuracy of set-based rare variant association tests. *Nat Genet* **54**, 1466–1469 (2022).
18. Stouffer, S. A. *The American Soldier ...: Adjustment during Army Life*. (1949).
19. Lin, X., Dey, R., Li, X. & Li, Z. Scalable analysis of large multi-ancestry biobanks by leveraging sparse ancestry-adjusted sample-relatedness. *Research Square* rs.3.rs–5343361 (2024).
20. Poterba, T. *et al.* The scalable variant call representation: enabling genetic analysis beyond one million genomes. *Bioinformatics* **41**, (2024).
21. Yengo, L. *et al.* A saturated map of common genetic variants associated with human height. *Nature* **610**, (2022).
